# Convergent large-scale network and local vulnerabilities underlie brain atrophy across Parkinson’s disease stages: a worldwide ENIGMA study

**DOI:** 10.1101/2025.05.25.25326586

**Authors:** Andrew Vo, Christina Tremblay, Shady Rahayel, Sarah Al-Bachari, Henk W Berendse, Joanna K Bright, Fernando Cendes, Emile d’Angremont, John C Dalrymple-Alford, Ines Debove, Michiel F Dirkx, Jason Druzgal, Gaëtan Garraux, Rick C Helmich, Michele Hu, Neda Jahanshad, Martin E Johansson, Johannes C Klein, Max A Laansma, Corey T McMillan, Tracy R Melzer, Bratislav Misic, Philip Mosley, Conor Owens-Walton, Laura M Parkes, Clelia Pellicano, Fabrizio Piras, Kathleen L Poston, Mario Rango, Christian Rummel, Petra Schwingenschuh, Melanie Suette, Paul M Thompson, Duygu Tosun, Chih-Chien Tsai, Tim D van Balkom, Odile A van den Heuvel, Ysbrand D van der Werf, Eva M van Heese, Chris Vriend, Jiun-Jie Wang, Roland Wiest, Clarissa Yasuda, Alain Dagher, ENIGMA-Parkinson’s Study

**Affiliations:** Department of Neurology and Neurosurgery, Montreal Neurological Institute, McGill University, Montreal, Canada; Department of Medicine, Centre for Advanced Research in Sleep Medicine, University of Montreal, Montreal, Canada; Faculty of Health and Medicine, Lancaster University, Lancaster, UK; Division of Neuroscience and Experimental Psychology, Faculty of Biology, Medicine and Health, The University of Manchester, Manchester, UK; Department of Neurology, Amsterdam UMC, Vrije Universiteit Amsterdam, Amsterdam, The Netherlands; Amsterdam Neuroscience, Neurodegeneration, Amsterdam, The Netherlands; Social, Genetic and Developmental Psychiatry Centre, Institute of Psychiatry, Psychology and Neuroscience, King’s College London, London, UK; Department of Neurology, University of Campinas–UNICAMP, Campinas, Brazil; Department of Anatomy and Neurosciences, Amsterdam UMC, Vrije Universiteit Amsterdam, Amsterdam, The Netherlands; Te Kura Mahi ā-Hirikapo | School of Psychology, Speech and Hearing, University of Canterbury, Christchurch, New Zealand; New Zealand Brain Research Institute, Christchurch, New Zealand; Department of Neurology, Inselspital, University of Bern, Bern, Switzerland; Department of Neurology and Center of Expertise for Parkinson & Movement Disorders, Donders Institute for Brain, Cognition and Behaviour, Radboud University Medical Center, Nijmegen, The Netherlands; Centre for Cognitive Neuroimaging, Donders Institute for Brain, Cognition and Behaviour, Radboud University, Nijmegen, The Netherlands; Department of Radiology and Medical Imaging, University of Virginia, Charlottesville, VA, USA; MoVeRe group, GIGA-CRC in vivo imaging, University of Liège, Liège, Belgium; Department of Neurology, CHU Liège, Liège, Belgium; Division of Clinical Neurology, Department of Clinical Neurosciences, Oxford Parkinson’s Disease Centre, Nuffield, University of Oxford, Oxford, UK; Imaging Genetics Center, Mark and Mary Stevens Neuroimaging and Informatics Institute, Keck School of Medicine, University of Southern California, Marina del Rey, CA, USA; Division of Clinical Neurology, Department of Clinical Neurosciences; Oxford Centre for Integrative Neuroimaging, Oxford Parkinson’s Disease Centre, Nuffield, University of Oxford, Oxford, UK; Department of Neurology, Perelman School of Medicine, University of Pennsylvania, University of Pennsylvania Perelman School of Medicine, Philadelphia, PA, USA; Department of Medicine, University of Otago, Christchurch, New Zealand; Pacific Radiology Canterbury, Christchurch, New Zealand; QIMR Berghofer Medical Research Institute, Queensland, Australia; Division of Psychology, Communication and Human Neuroscience, School of Health Sciences, Faculty of Biology, Medicine and Health, The University of Manchester, Manchester, UK; Geoffrey Jefferson Brain Research Centre, Manchester Academic Health Science Centre, Northern Care Alliance & University of Manchester, Manchester, UK; Laboratory of Neuropsychiatry, IRCCS Santa Lucia Foundation, Rome, Italy; Department of Neurology & Neurological Sciences, Movement Disorders, Stanford University, Palo Alto, CA, USA; Excellence Interdepartmental Center for Advanced MR Techniques and Department of Neurosciences, Neurology Unit, Parkinson’s Disease Center, Fondazione Cà Granda, IRCCS, Ospedale Policlinico, University of Milan, Milan, Italy; Support Center for Advanced Neuroimaging (SCAN), University Institute of Diagnostic and Interventional Neuroradiology, University Hospital Bern, Bern, Switzerland; Department of Neurology, Medical University of Graz, Graz, Austria; Department of Radiology and Biomedical Imaging, University of California San Francisco, San Francisco, CA, USA; Healthy Ageing Research Center, Chang Gung University, Taoyuan City; Department of Psychiatry, Amsterdam UMC, Vrije Universiteit Amsterdam, Amsterdam, The Netherlands; Amsterdam Neuroscience, Brain Imaging, Amsterdam, The Netherlands; Department of Medical Imaging and Radiological Sciences, Chang Gung University, Taoyuan City, Taiwan; Brazilian Institute of Neuroscience and Neurotechnology, Campinas, Brazil

**Keywords:** Parkinson’s disease, neurodegeneration, structural MRI, connectivity, imaging transcriptomics

## Abstract

Parkinson’s disease (PD) is associated with extensive structural brain changes. Recent work has proposed that the spatial pattern of disease pathology is shaped by both network spread and local vulnerability. However, only few studies assessed these biological frameworks in large patient samples across disease stages. Analyzing the largest imaging cohort in PD to date (N = 3,096 patients), we investigated the roles of network architecture and local brain features by relating regional abnormality maps to normative profiles of connectivity, intrinsic networks, cytoarchitectonics, neurotransmitter receptor densities, and gene expression. We found widespread cortical and subcortical atrophy in PD to be associated with advancing disease stage, longer time since diagnosis, and poorer global cognition. Structural brain connectivity best explained cortical atrophy patterns in PD and across disease stages. These patterns were robust among individual patients. The precuneus, lateral temporal cortex, and amygdala were identified as likely network-based epicentres, with high convergence across disease stages. Individual epicentres varied significantly among patients, yet they consistently localized to the default mode and limbic networks. Furthermore, we showed that regional overexpression of genes implicated in synaptic structure and signalling conferred increased susceptibility to brain atrophy in PD. In summary, this study demonstrates in a well-powered sample that structural brain abnormalities in PD across disease stages and within individual patients are influenced by both network spread and local vulnerability.

## INTRODUCTION

Parkinson’s disease (PD) is a progressive neurodegenerative disorder marked by extensive structural changes in the brain, affecting both cortical and subcortical regions [1–3]. However, the spatial pattern of atrophy is not uniform across the brain. Some regions show greater vulnerability to disease pathology than others. This raises questions as to what the underlying factors are that shape and drive the spread of pathology in PD.

Early post-mortem studies describe a distribution pattern of Lewy pathology in PD that appears to map onto large-scale intrinsic networks in the brain [4, 5]. This Braak staging implies that the spread of PD pathology is not random, but constrained by the organization of the underlying connectome [6, 7]. It has been hypothesized that this network spreading process involves the propagation of misfolded alpha-synuclein protein via neuronal synapses in a prion-like manner [7, 8]. The primary support for this hypothesis comes from animal studies that traced the neuronal spread of injected alpha-synuclein [9–11]. In addition, there is mounting evidence in support of this hypothesis derived from patient populations using non-invasive brain imaging and computational modelling [3, 12–17]. The progression of PD pathology does not always align neatly with the Braak staging framework however [18], suggesting that network spreading is not the only driver of pathology. Indeed, local vulnerability features, such as cellular composition [19], metabolic demands [20], or gene expression [21], may predispose certain brain regions to disease pathology and damage. Studies employing imaging transcriptomics to explore the relationship between brain morphometry in PD and transcriptional gene activity, for example, have shown that the local expression of genes related to synaptic, mitochondrial, and metabolic activity render certain brain regions particularly susceptible to atrophy [15, 16, 22, 23].

Studies of structural brain abnormalities, as well as network spread and local vulnerability in PD, have so far been limited to relatively small samples of clinically heterogeneous patients. Moreover, these studies are complicated by variability in analytic approaches between different study sites. The Enhancing Neuroimaging Genetics through Meta-analysis PD (ENIGMA-PD) working group is an international collaboration across multiple centres that has curated and harmonized the largest imaging dataset in PD to date [1, 24, 25]. Here, we analyzed this well-powered dataset to map cortical and sub-cortical atrophy in PD, across disease stages, and within single subjects. We then related these spatial atrophy patterns to normative network models, and consistently found that cortical network connectivity constrained and identified likely epicentres of pathology spread. Next, we investigated whether PD atrophy mapped onto specific intrinsic functional networks, tissue cytoarchitectonics, or neuroreceptor densities. Finally, we performed imaging transcriptomic analysis and were able to show that gene expression profiles related to the atrophy patterns were associated with synaptic structure and signalling. Taken together, our results demonstrate that structural brain abnormalities in PD across disease stages and within individual patients are influenced by both network spread and local vulnerability.

## RESULTS

### Participants

The ENIGMA-PD working group collected and processed T1-weighted brain MRI and clinical data across 23 international sites, yielding a final sample of 3,096 PD patients and 1,262 healthy controls (HC). Table 1 displays the demographic and clinical details of the participants included in the study. Supplementary Table S1 details the demographic and clinical characteristics of the participants for each contributing site and Supplementary Table S2 details the inclusion/exclusion criteria. The PD group was significantly older than the HC group (*t*(2522) = -5.24, *P* < 0.001). Although the age range of the HC group (40-85 years) did not entirely cover the span of the PD group (40-89 years), only seven PD patients (<1%) exceeded this range, which suggests an overall sufficient overlap for deriving atrophy maps using *w*-scoring (see *Methods* for details). Critically, a Levene’s test demonstrated that the two groups did not differ in the variance of ages (*W*(1,4358) = 1.00, *P* = 0.316), indicating comparable age distributions. The proportion of males to females was also significantly different between groups (*χ*^2^ = 44.95, *P* < 0.001). Despite these group differences in age and sex, a sensitivity analysis comparing atrophy maps derived from the complete sample and an age- and sex-matched subsample (see Supplementary Table S3) demonstrated comparable patterns of structural abnormalities (Supplementary Fig. S1a and S1b). For PD patients with available information on Hoehn and Yahr (HY) disease stages [26] (82.5% of total sample), the majority were classified as HY 2.

**TABLE 1:** ENIGMA-PD sample demographics and clinical details.

| Group | HY stage | N | Age<br>(mean ± SD) | Sex<br>(% female) | Time since<br>diagnosis<br>(mean ± SD) | MoCA<br>(mean ± SD) |
| --- | --- | --- | --- | --- | --- | --- |
| HC | – | 1262 | 62.09 ± 9.25 | 47.31 | – | 27.88 ± 1.70<sup>b</sup> |
| PD | – | 3096 | 63.69 ± 9.09 | 36.30 | 5.18 ± 5.11<sup>a</sup> | 25.89 ± 3.29<sup>b</sup> |
| | 1 | 466 | 60.15 ± 8.63 | 40.56 | 2.38 ± 2.52 | 27.20 ± 2.32 |
| | 2 | 1596 | 63.90 ± 8.80 | 34.21 | 4.50 ± 4.43 | 26.26 ± 2.91 |
| | 3 | 397 | 66.05 ± 9.43 | 40.81 | 7.76 ± 5.98 | 24.80 ± 3.74 |
| | 4/5 | 96 | 67.48 ± 9.38 | 38.54 | 13.05 ± 6.42 | 21.41 ± 4.88 |

HC = healthy controls; PD = Parkinson’s disease; SD = standard deviation; HY stage = Hoehn and Yahr disease stage; MoCA = Montreal Cognitive Assessment score. <sup>a</sup>Information available in 2,879 of 3,096 patients.
<sup>b</sup>Information available in 1,796 of 3,096 patients and 454 of 1,262 controls.

### Widespread structural brain abnormalities in Parkinson’s disease

We harmonized regional cortical thickness, cortical surface area, and subcortical volume estimates in PD patients and HCs across all contributing sites. All analyses were performed using the 68-region Desikan-Killiany atlas [27]. Individual age- and sex-adjusted maps of brain abnormalities in PD patients were derived using *w*-scoring (see *Methods* for details) and reflect regional deviations of a given brain measure from what would be expected in the HC reference group. To investigate PD-related deviations in these *w*-maps, one-sample *t*-tests compared regional mean *w*-scores to zero. We found statistically significant and diffuse negative deviations in cortical thickness that were most pronounced in parietal and temporal regions (Fig. 1a and Supplementary Table S4a). Cortical surface area was also generally lower in PD, with the greatest negative deviations in occipital and middle frontal cortex (Fig. 1a and Supplementary Table S4b). Finally, grey matter volumes were lower across the majority of subcortical nuclei, with peak negative deviations in the putamen and amygdala. The volume of the left thalamus and lateral ventricles were higher than expected, however (Fig. 1a and Supplementary Table S4c). When stratified by HY disease stage scores, the number of regions demonstrating significant deviations and the magnitude of these abnormalities increased with higher disease stage (see Supplementary Figure S2). Overall, these findings show a widespread pattern of reduced cortical and subcortical grey matter in PD.

**Figure 1:**
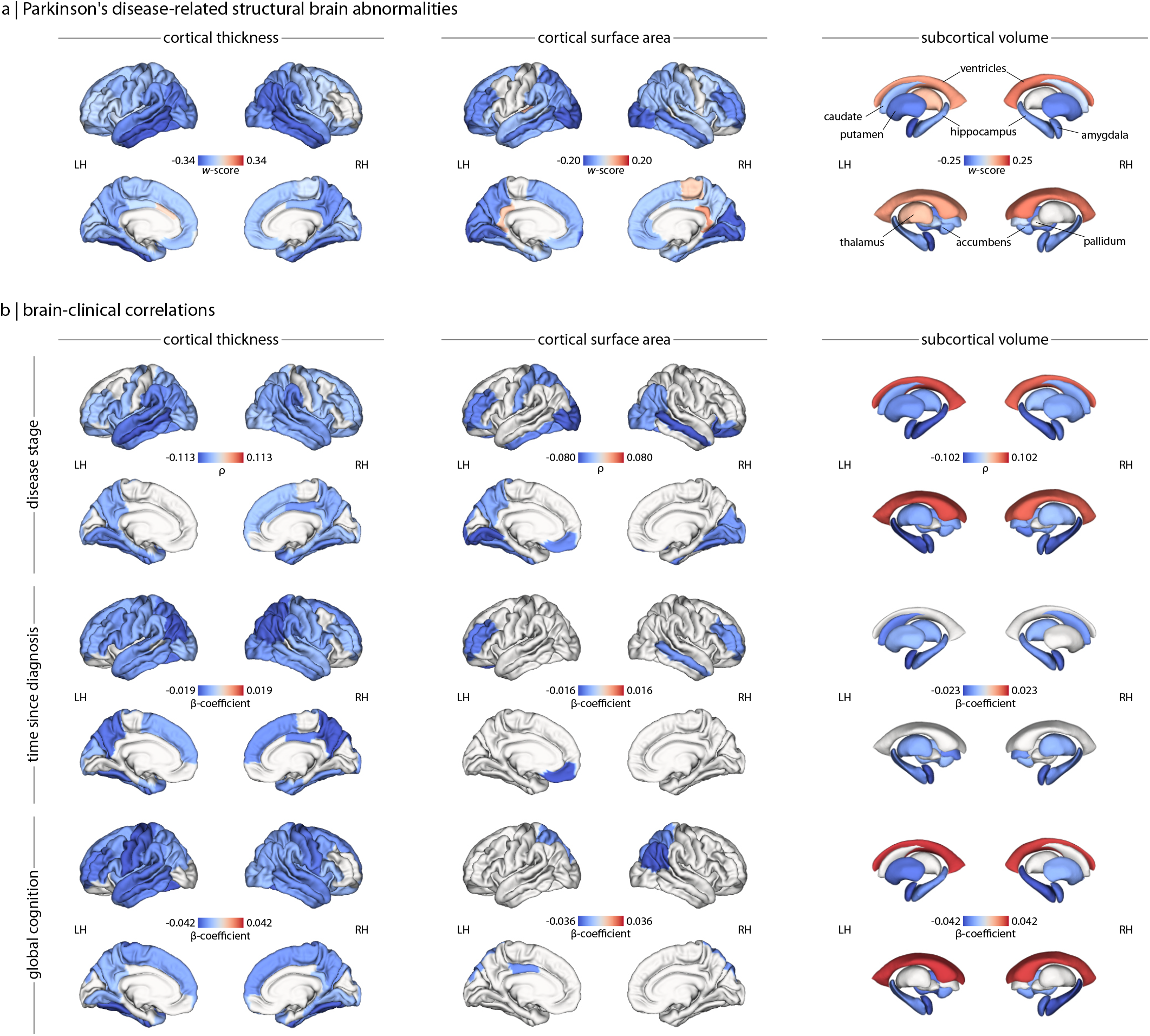
Structural brain abnormalities in PD and their clinical correlates. **(a)** *W*-score maps of cortical thickness, cortical surface area, and subcortical volume deviations reveal a widespread pattern of atrophy in PD. More negative *w*-scores (or bluer regions) represent lower estimates or greater atrophy in PD patients relative to what would be expected in the healthy reference group. **(b)** Partial rho (*ρ*) and *β*-coefficient maps of brain-clinical correlations between cortical thickness, cortical surface area, and subcortical volume deviations and Hoehn and Yahr disease stage, time since diagnosis (in years), and global cognition (Montreal Cognitive Assessment scores), controlling for age and sex effects. Overall, more negative deviations in brain measures were related to advancing disease stage, longer disease durations, and poorer cognition. For display purposes, only regions surviving FDR correction for multiple comparisons (*P*_*FDR*_ < 0.05) are shown. LH = left hemisphere; RH = right hemisphere.

Next, we explored the relationships between regional brain abnormalities and clinical scores in PD, adjusting for age and sex. Rank-based partial correlations revealed regional deviations became more negative with higher disease stages for all brain measures, except for ventricular volumes that displayed the inverse relationship. These patterns are consistent with a progressive atrophic process in PD. Similarly, linear regressions showed time since diagnosis was negatively correlated with each brain measure, such that longer durations corresponded with more negative deviations in regional cortical thickness, cortical surface area, and subcortical volume (Fig. 1b and Supplementary Tables S5a-c). Global cognition, assessed with the Montreal Cognitive Assessment (MoCA) [28], was also negatively correlated with each brain measure such that poorer cognition was related to more negative deviations in cortical thickness and subcortical grey matter volume but more positive deviations in the lateral ventricles. In summary, we found that cortical thickness and subcortical volume abnormalities appear to be most sensitive to PD-related clinical features.

### Network architecture shapes cortical atrophy patterns across disease stages and in individual patients

It has been proposed that PD pathology advances through the brain via a network spreading process [6–8]. To test the hypothesis that the atrophy pattern in PD is shaped by network architecture, we related regional abnormality (or “nodal atrophy”) to the abnormality across structurally connected regions (or “neighbourhood atrophy”). We defined nodal atrophy as the cortical thickness and subcortical volume deviations in PD, and neighbourhood atrophy of a given node as the mean abnormality of structurally connected nodes weighted by the strength of structural or functional connectivity. The connectivity profile of each node was determined by connectivity matrices derived from diffusion-weighted imaging and resting-state MRI in a separate group of healthy participants [29]. A total of four normative network models, representing the connectivity among cortical regions (“cortico-cortical”) or between cortex and subcortex (“subcortico-cortical”), were used to inform our calculation of neighbourhood atrophy: (i) cortico-cortical structural network, (ii) subcortico-cortical structural network, (iii) cortico-cortical functional network, and (iv) subcortico-cortical functional network. For each network model, we examined the node-neighbourhood relationships in the group average, disease stage, and single-subject atrophy maps.

In the group average maps (Fig. 2b, top row and Supplementary Table S6a), nodal atrophy was positively correlated with neighbourhood atrophy for both structural (*ρ* = 0.545, *P*_*spin*_ = 0.001) and functional (*ρ* = 0.366, *P*_*spin*_ = 0.071) cortico-cortical network models, though the latter was no longer statistically significant after spatial null testing. Structural and functional subcortico-cortical network models poorly explained the node-neighbourhood correlation, however (both *P*_*perm*_ > 0.05). For the disease stage maps (Fig. 2b, middle row and Supplementary Table S6b), in which patients were stratified by their HY stage, we found that the atrophy pattern was again best explained by cortico-cortical network models. Structural connectivity accounted for cortical atrophy patterns across HY stages 1 to 4/5, and this finding was robust when tested against spatial null models. Similarly, functional connectivity informed the cortical atrophy pattern across early HY stages (HY 1-3), but node-neighbourhood atrophy did not survive spatial null testing beyond HY stage 1 (Fig. 2b, middle row and Supplementary Table S6b). In subcortico-cortical network models, subcortical atrophy patterns were not well explained by either structural or functional connectivity at any disease stage (all *P*_*perm*_ > 0.05). In short, corticocortical networks better reflect node-neighbourhood atrophy than subcortico-cortical networks, especially when weighted by structural compared to functional connectivity.

**Figure 2:**
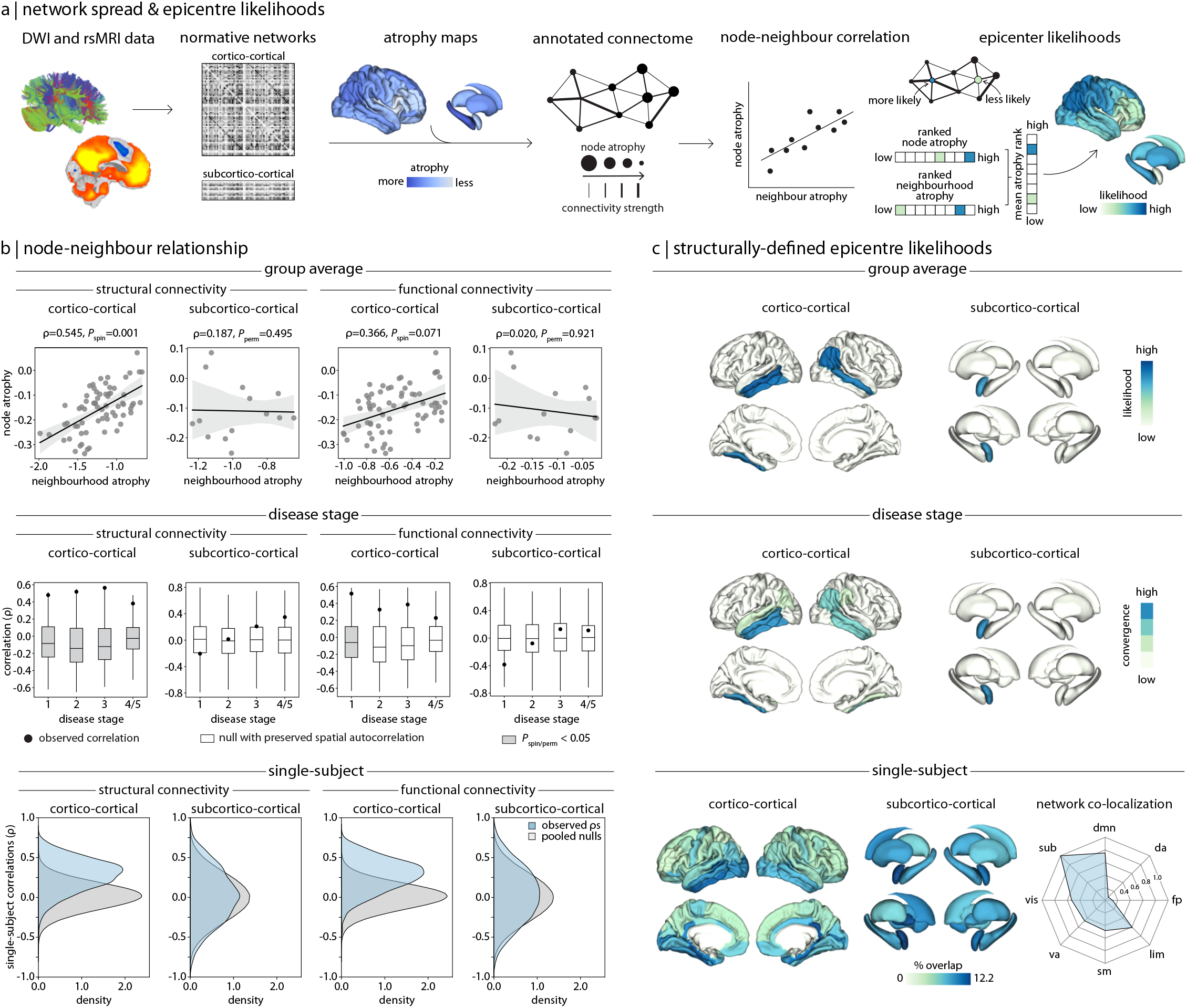
Network architecture shapes the pattern of atrophy in PD. **(a)** Schematic of network-based disease exposure and epicentre likelihood workflows. Structural and functional connectivity was defined by cortico-cortical and subcortico-cortical normative network models derived from an unrelated cohort of healthy participants. This information was used to relate regional abnormality (or “node atrophy”) to the average abnormality across connected neighbour regions (or “neighbourhood atrophy”). The mean rank of node and neighbourhood atrophy was used to identify regions as likely epicentres. **(b)** Node-neighbourhood atrophy correlations revealed that atrophy patterns were best explained by cortico-cortical structural network models (top row), across disease stages (middle row), and in a large proportion of individual patients (bottom row), followed by cortico-cortical functional network models. Neither subcortico-cortical structural or functional network models explained node-neighbourhood coupling, however. **(c)** Epicentre likelihoods identified the precuneus, lateral temporal cortex, and amygdala as network-based epicentres (top row), which were consistently identified across disease stages (middle row). Although single-subject epicentres did not frequently generalize across individual patients (bottom row), they were found to co-localize to common networks (*dmn*: default mode, *da*: dorsal attention, *fp*: frontoparietal, *lim*: limbic, *sm*: sensorimotor, *va*: ventral attention; *vis*: visual). For all brain visualizations, only regions surviving spatial null testing (*P*_*spin/perm*_ < 0.05) are displayed.

Finally, for single-subject atrophy maps (Fig. 2b, bottom row), we observed high stability in node-neighbourhood atrophy coupling for both structural (*P*_*spin*_ < 0.05 in 59.43% of patients) and functional (*P*_*spin*_ < 0.05 in 59.04% of patients) cortico-cortical network models. In contrast, there was low stability between node-neighbourhood atrophy in structural (*P*_*perm*_ < 0.05 in 5.98% of patients) and functional (*P*_*perm*_ < 0.05 in 6.40% of patients) subcortico-cortical models. Despite the large degree of heterogeneity expected across these individualized atrophy maps, a large proportion of PD patients still demonstrated connectivity-based cortical atrophy patterns observed in the group average. As before, cortico-cortical networks explained atrophy patterns better than subcortico-cortical networks.

### Network-based epicentres converge across disease stage and co-localize to common networks

As a follow-up to our previous analysis, we sought to identify network-based epicentre regions that are both greatly affected by and promote the spread of PD pathology. The epicentre likelihood of each region was determined by the mean rank of their node and neighbourhood atrophy, such that regions with high node and neighbourhood atrophy were considered more likely epicentres. We estimated network-based epicentres for each normative network model and for the group average, disease stage, and single-subject atrophy maps.

In the group average maps (Fig. 2c, top row), we identified the right precuneus and bilateral lateral temporal cortex, and left amygdala as having high epicentre likelihoods using both cortico-cortical and subcortico-cortical network models, respectively. These likely epicentres demonstrated a high degree of atrophy and were themselves connected to neighbourhoods of highly atrophied regions. Similarly, these regions were consistently identified as likely epicentres across the disease stage maps (Fig. 2c middle row), showing high convergence across the four HY stages. Network models defined by structural and functional connectivity resulted in the same epicentres (see Supplementary Fig. S3a and S3b). These findings suggest that these identified regions are effective propagators of PD pathology.

Repeating this approach in single-subject atrophy maps (Fig. 2c, bottom row), the right isthmus cingulate showed the highest percentage of convergence across individual PD patients—although the maximum overlap was only 12.2% of patients. This was unsurprising, however, given the large degree of heterogeneity expected at the single-subject level. When we mapped each PD patient’s likely epicentres to intrinsic brain networks, these disparate individualized epicentres co-localized to the default mode and limbic networks (75.48% and 60.92% of PD patients, respectively) and the subcortex (100% of PD patients). Therefore, although individual patient epicentres highly vary, they belong to shared common networks.

### Cortical abnormalities are distributed in specific brain systems

Beyond network structure, we also studied whether local brain features contributed to the atrophy pattern in PD. We began by testing whether cortical abnormalities were more or less prominent within specific macro-scale cortical systems. We calculated the mean cortical thickness and surface area deviations within seven intrinsic resting-state networks defined by Yeo et al. [30] and seven von Economo cytoarchitectonic tissue classes [31, 32]. For the functional resting state networks, mean cortical thickness deviations were more negative in the default mode network (*P*_*spin*_ = 0.049) but less so in the ventral attention network (*P*_*spin*_ = 0.015; Fig. 3a). For cytoarchitectonic classes, these deviations were also found to be more negative in association cortex (*P*_*spin*_ = 0.009; Fig. 3b). Mean cortical surface area deviations were mainly distributed in visual and sensorimotor networks, with more negative deviations in the visual (*P*_*spin*_ = 0.002) but less pronounced in the sensorimotor network (*P*_*spin*_ = 0.006; Fig. 3a). They were also relatively more negative in primary/secondary sensory cortex (*P*_*spin*_ = 0.006) but less severe in limbic (*P*_*spin*_ = 0.005) and primary motor systems (*P*_*spin*_ = 0.017; Fig. 3b). These findings suggest that cortical PD pathology primarily affects higher-order, transmodal areas compared to unimodal cortex.

**Figure 3:**
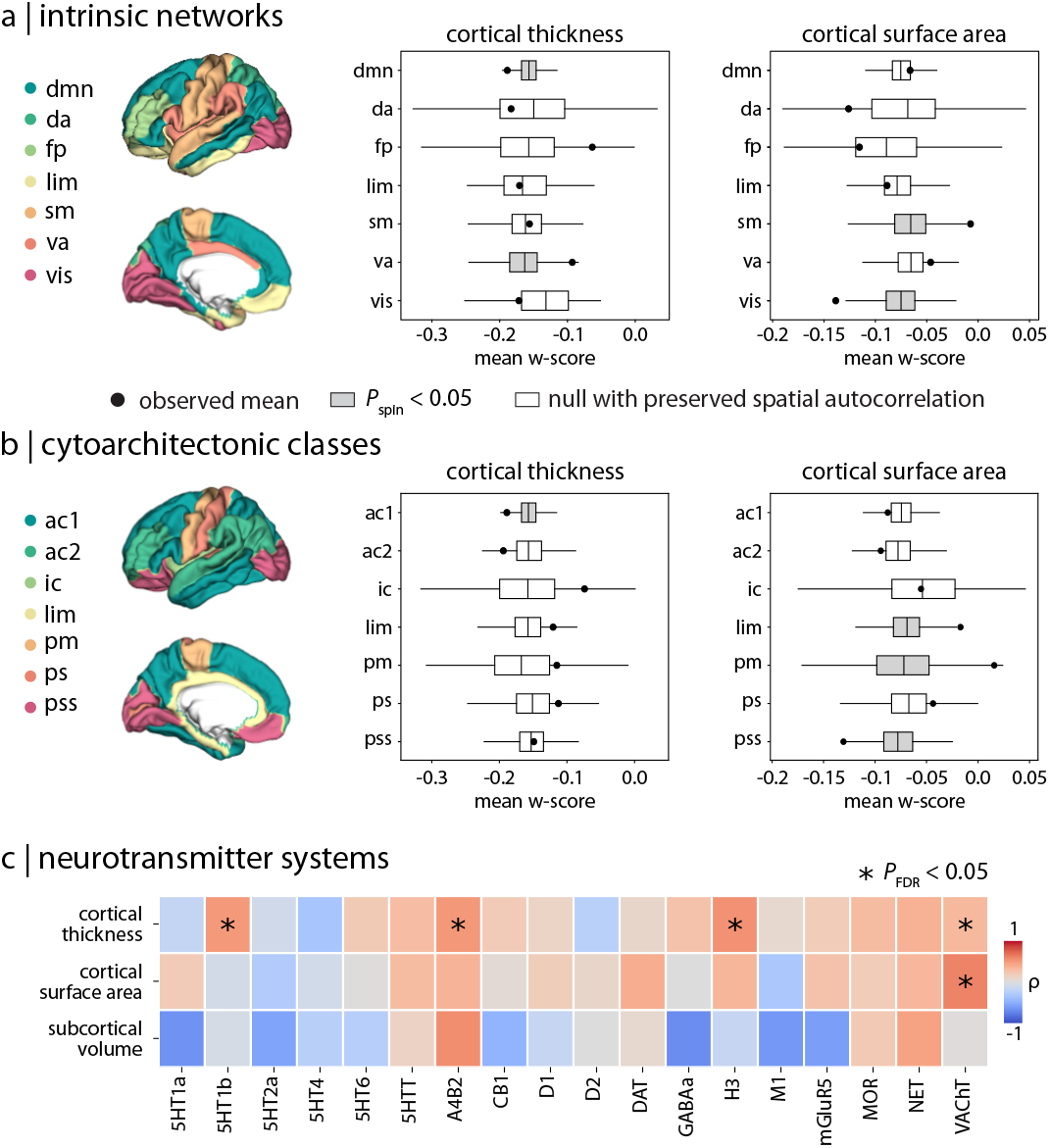
Distribution of PD-related atrophy in specific brain systems. Cortical thickness (left) and cortical surface area (right) abnormalities in PD are localized within **(a)** intrinsic resting networks [30] (*dmn*: default mode, *da*: dorsal attention, *fp*: frontoparietal, *lim*: limbic, *sm*: sensorimotor, *va*: ventral attention; *vis*: visual) and **(b)** cytoarchitectonic tissue classes [31, 32] (*ac1/2*: association 1/2, *ic*: insular; *lim*: limbic, *pm*: primary motor, *ps*: primary sensory; *pss*: primary/secondary sensory). **(c)** Correlations between regional brain atrophy and expression of 18 neurotransmitter systems [33]: acetylcholine (*α*_4_*β*_2_, M_1_, VAChT), cannabinoid (CB_1_), dopamine (D_1_, D_2_, DAT), GABA (GABA_*A/BZ*_, histamine (H_3_), glutamate (mGluR_5_, NMDA), norepinephrine (NET), opioid (MOR), and serotonin (5-HT1_*A*_, 5-HT1_*B*_, 5-HT2_*A*_, 5-HT_4_, 5-HT_6_, 5-HTT). * indicates correlations that survived both spatial null testing (*P*_*spin/perm*_ < 0.05) and FDR correction for multiple comparisons (*P*_*FDR*_ < 0.05).

We then asked if the pattern of brain abnormalities in PD was related to molecular profiles of neurotransmitter systems. Positron emission tomography tracer maps for 18 distinct neurotransmitter receptors and transporters from different cohorts of healthy participants were correlated with regional cortical thickness, surface area, and subcortical volume deviations in PD [33, 34]. Cortical thickness deviations were significantly related to several neurotransmitter distributions (Fig. 3c and Supplementary Table S7a), including 5-HT1_*B*_ receptor (*ρ* = 0.493, *P*_*FDR*_ = 0.018), nicotinic *α*_4_*β*_2_ receptor (*ρ* = 0.503, *P*_*FDR*_ = 0.024), histamine H_3_ receptor (*ρ* = 0.537, *P*_*FDR*_ = 0.018), and vesicular acetylcholine transporter (VAChT; *ρ* = 0.322, *P*_*FDR*_ = 0.031). Cortical surface area deviations were also significantly correlated with VAChT (*ρ* = 0.595, *P*_*FDR*_ = 0.018; Fig. 3c and Supplementary Table S7b). In each case, there was a positive association between the brain measure and neurotransmitter receptor density, suggesting that greater atrophy was observed in regions with lower expression of these systems (Supplementary Fig. S4). No correlations involving subcortical volumes survived FDR correction for multiple comparisons, however (see Supplementary Table S7c). These results suggest that cortical areas expressing select serotonergic, histaminergic, and cholinergic receptors and transporters might be less vulnerable to PD atrophy.

### Cortical atrophy is associated with synapse-related gene expression profiles

To understand the biological and cellular underpinnings of the atrophy pattern in PD, we integrated our imaging findings with brain-wide gene expression data and explored the biological relevance of the associated genes. We first applied partial least squares (PLS) analysis to identify the multivariate relationship between cortical thickness deviations and the expression profiles of 15,633 genes obtained from the Allen Human Brain Atlas [35]. PLS analysis identified a single significant latent variable (PLS1) that explained 48.53% (*P*_*spin*_ = 0.044) of the covariance between the cortical atrophy pattern and gene expression (Fig. 4b). Regional weights associated with PLS1 were positively correlated with the cortical atrophy pattern (*ρ* = 0.620, *P*_*spin*_ = 0.008), such that more negatively weighted genes were associated with more negative deviations in cortical thickness (Fig. 4c and 4d). Therefore, this analysis identifies genes with expression profiles associated with greater vulnerability to PD atrophy.

**Figure 4:**
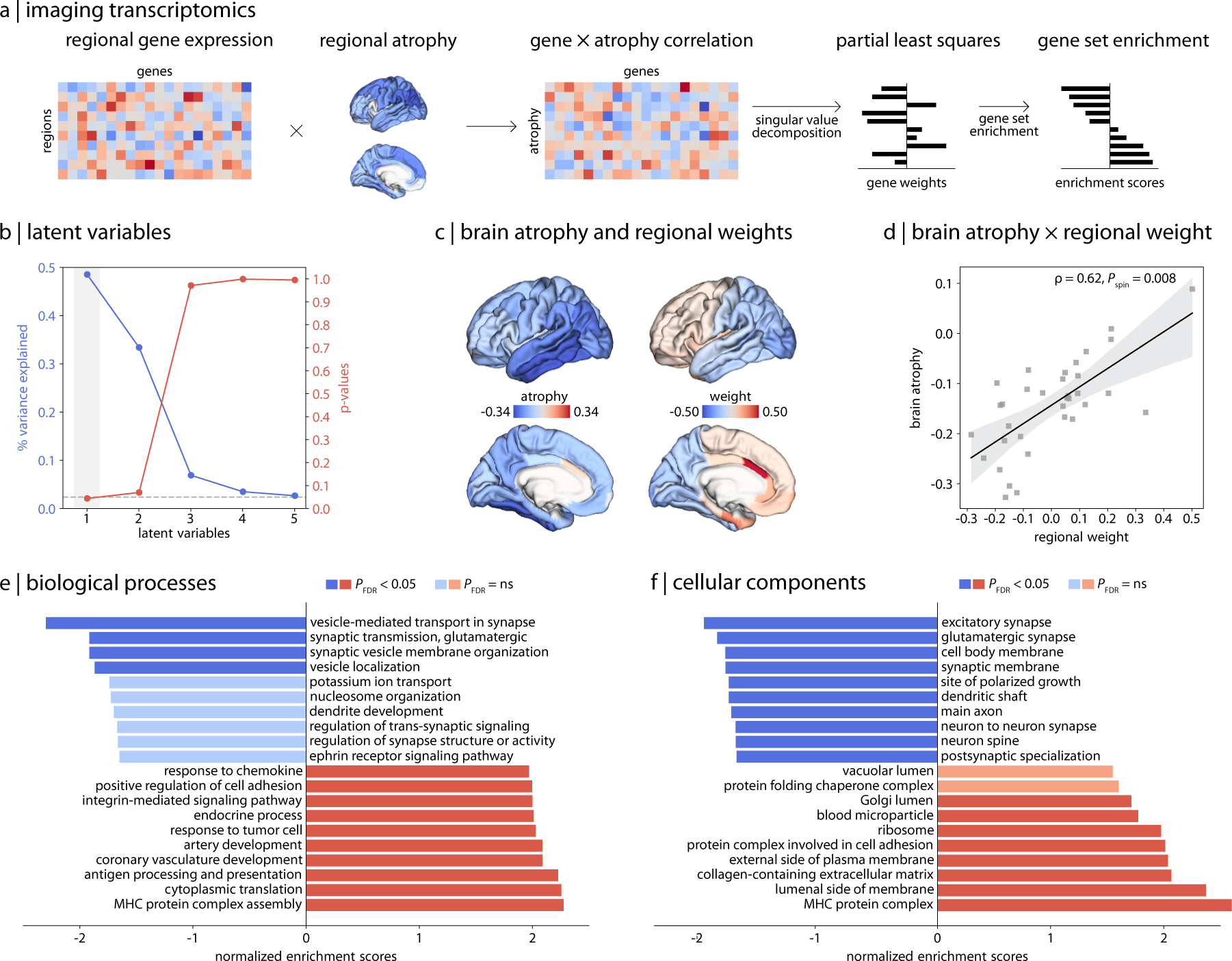
Relationship between cortical atrophy in PD and gene expression. **(a)** Schematic of the imaging transcriptomics workflow. **(b)** PLS analysis identified a single latent variable (PLS1) that significantly explained 48.53% of the covariance between cortical thickness deviations and gene expression profiles. **(c)** Regional weights were **(d)** negatively correlated with regional brain atrophy, indicating that more positive weights were associated with greater cortical atrophy. Gene set enrichment analysis revealed that genes most correlated with cortical atrophy are enriched for **(e)** synaptic regulation and signalling and **(f)** synapse and neuron components.

Next, genes were ranked from most positively to most negatively weighted on PLS1 and interpreted for their biological relevance using gene set enrichment analysis (GSEA). This analysis revealed that regions with greater cortical atrophy were found to overexpress genes related to synaptic signalling and regulation. Concerning biological processes (Fig. 4e and Supplementary Table S8a), the genes most associated with cortical atrophy were enriched for terms such as “vesicle-mediated transport in synapse” (normalized enrichment score [*NES*] = -2.305, *P*_*FDR*_ < 0.001), “synaptic transmission, glutamatergic” (*NES* = -1.921, *P*_*FDR*_ = 0.045), “synaptic vesicle membrane organization” (*NES* = -1.921, *P*_*FDR*_ = 0.031), and “vesicle localization” (*NES* = -1.874, *P*_*FDR*_ = 0.044). Positively weighted genes were significantly enriched for diverse functional biological processes. In terms of cellular components (Fig. 4f and Supplementary Table S8b), genes most strongly related to cortical atrophy were enriched for components localized to the synapse and neuron, such as “excitatory synapse” (*NES* = -2.068, *P*_*FDR*_ = 0.009), “synaptic membrane” (*NES* = -1.877, *P*_*FDR*_ = 0.014), and “main axon” (*NES* = -1.826, *P*_*FDR*_ = 0.013). For genes least associated with cortical atrophy, significantly enriched terms included structural cellular components. Collectively, these analyses show that cortical regions with higher expression of genes related to synapses demonstrate greater atrophy in PD.

## DISCUSSION

We mapped structural brain abnormalities in PD and contextualized this spatial pattern using connectomics, annotation enrichment, and imaging transcriptomic approaches. Global reductions in cortical thickness, cortical surface area, and subcortical volumes were found in PD relative to HCs. Regional cortical thickness and sub-cortical volume reductions were most sensitive to measures of advancing disease stage, longer disease duration, and cognitive decline. We then showed that the pattern of PD-related cortical atrophy to be constrained by network architecture. Network-based epicentres were identified in the precuneus, temporal lobes, and amygdala, which generalized across disease stages and co-localized to default mode and limbic networks in individual patients. We also found that cortical thickness and surface area deficits mapped onto specific brain systems related to distinct intrinsic networks, cytoarchitectonic tissue classes, and neurotransmitter receptor profiles. Finally, we demonstrated that the pattern of cortical atrophy correlated with gene expression profiles implicated in synaptic and neuronal processes.

Structural brain abnormalities in PD were characterized by widespread reductions in cortical thickness, cortical surface area, and subcortical volume. These findings reproduce—in an updated sample—the atrophy pattern first reported by Laansma et al. [1]. However, our findings of morphometric differences in PD were generally inconsistent with previous case-control studies in terms of location and size of effects [36]. These discrepancies likely stem from limited sample sizes, clinical heterogeneity, and variability in analytical methods used across studies. We address these limitations by harmonizing data across multiple sites to create a large, well-powered sample and applying standardized analysis methods. The resulting atrophy pattern provides a map against which future, smaller PD studies can be benchmarked. Among the structural brain metrics, cortical thickness and sub-cortical volume deviations were most strongly correlated with clinical features, including more advanced disease stages, longer disease durations, and poorer global cognition. This relationship between PD atrophy and measures of disease severity and cognitive impairment are well-established in the literature [1, 37, 38]. Critically, our results are based on a significantly larger sample size spanning a wider disease course than most previous work.

Nodal atrophy was correlated with neighbourhood atrophy such that regions with greater atrophy were more strongly connected to neighbours with collectively greater atrophy themselves. This relationship was best captured when normative connectivity profiles were defined by structural cortico-cortical network models and was consistent across disease stages. Node-neighbourhood correlations were notably stable among single-subject atrophy maps, suggesting network spreading as a fundamental underlying mechanism in PD. Our results extend prior findings in prodromal and *de novo* PD [3, 12–17, 39] to the entire disease course and within individual patients. Similarly, a network spreading process has also been demonstrated in other neurodegenerative diseases and psychiatric disorders, including Alzheimer’s disease, frontotemporal dementia, amyotrophic lateral sclerosis, and schizophrenia [6, 20, 40– 44]. This repeated observation is consistent with the theory that neurodegeneration in PD is in part driven by synaptic transmission of pathogenic alpha-synuclein [7, 8].

In contrast to our cortical-driven results, subcortical atrophy patterns were poorly explained by subcortico-cortical network models. This lack of an association might be explained by the subcortical atlas used in this study, which features a limited number of coarsely parcellated regions, and is the native and only available representation of ENIGMA data. Limited granularity in the subcortex may not adequately capture the spatial variance of PD pathology across substructures [45, 46]. Likewise, the subcortico-cortical connectivity profiles generated with this parcellation might fail to capture the spatial gradients of connectivity between the subcortex and cortex [47–50]. Future studies should use atlases with multiple available resolutions to test dependency of results on spatial scale information. Alternatively, distinct connectivity profiles between cortex versus subcortex might explain their differential ability to inform patterns of disease pathology. Whereas cortical connectivity is highly distributed and hierarchical with many interconnections, subcortical connectivity is instead modular and specialized with unidirectional relay connections [51]. Consequently, cortico-cortical networks may better reflect the widespread atrophy patterns in PD compared to subcortico-cortical networks.

Connectome-based atrophy modeling also allowed the identification of cortical disease epicentres, here thought to represent regions that may act as efficient propagators of pathology. At the group level, the precuneus, lateral temporal cortex, and amygdala emerged as network-based epicentres, displaying high convergence across disease stages. The precuneus and lateral temporal cortex are central hubs of the default mode network, exhibiting widespread connections with other brain areas [52–54]. Such connectivity profiles well-position these regions as propagators of disease pathology. We also identified patient-specific epicentres: although we observed considerable regional variability in individualized epicentres, they broadly mapped to the default mode and limbic networks, and to subcortical nuclei. Importantly, the epicentres thus defined are not necessarily the originators of pathology, which are likely located outside the cerebral cortex [3]. For example, according to the alpha-synuclein Origin site and Connectome model [55], the amygdala and olfactory bulb are important origin sites of pathology in a brain-first subtype of PD whereas pathology originates from the enteric nervous system and brainstem in the body-first subtype. Overall, however, our findings support the robust contribution of default mode, limbic, and subcortical regions, along with their associated network architecture, in shaping the atrophy pattern in PD.

Cortical thickness abnormalities in PD mapped predominantly onto the default mode network, which aligns with our finding of individualized epicentres co-localizing to this specific network. Previous studies have also reported cortical thinning in key regions of the default mode network, such as the precuneus and posterior cingulate cortex, suggesting a consistent pattern of vulnerability in PD [38, 56, 57]. These structural changes are complemented by findings from functional MRI studies in PD that have demonstrated altered connectivity within this network [58–60]. While the motor symptoms of PD are mainly caused by substantia nigra dopamine neuron loss, default mode network dysfunction likely accounts for the cognitive and mood symptoms that are a hallmark of later disease stages [61–64].

We also found greater cortical abnormalities in regions with lower normative expression of specific serotonergic, histaminergic, and cholinergic neurotransmitter systems. The 5-HT1_*B*_ receptor has previously been attributed a neuroprotective role, as serotonin receptor agonism reduced alpha-synuclein deposition and oxidative stress in a rodent model of PD [65]. Similarly, the *α*_4_*β*_2_ nicotinic acetylcholine receptor mediates the neuroprotective effects of nicotine against PD pathology [66]. Thus, regions with lower expression of these neuroreceptor systems could exhibit greater susceptibility to neurodegeneration, but further investigation is needed.

Cortical atrophy in PD was associated with gene expression profiles enriched for synaptic and neuronal terms, suggesting a link between disease pathology and underlying biological mechanisms. Our finding replicates previous imaging transcriptomic studies in PD demonstrating a link between grey matter atrophy or iron accumulation and genes related to synaptic transmission and signalling [15, 23]. Alpha-synuclein protein normally plays an important role in the regulation of synaptic function. In PD, misfolded, pathogenic alpha-synuclein has been shown to aggregate in presynaptic terminals, disrupting neurotransmission, and eventually leading to neuronal death [67–69]. This loss of neuropil—the synapses, axons, and dendrites within cortical columns—would be reflected as cortical thinning [70]. In sum, the overexpression of genes related to synapses and neuropil components may identify cortical regions with heightened vulnerability to PD pathology.

We examined the contributions of network structure and local vulnerability to the pattern of atrophy in PD independently; however, it is important to consider that these biological principles are not mutually exclusive but rather interactive. In other words, the spread of pathology might be constrained to the connectivity between regions but its impact on regional morphometry or function may be modulated by local features. This notion is clearly demonstrated by agent-based simulations of pathology spread in PD [22, 71, 72]. Zheng et al. [72] used a susceptible-infected-removed model to simulate the propagation of alpha-synuclein along a structural connectome anchored by regional expression of genes that modify pathogenic protein levels. They showed that this approach could successfully recreate *in silico* the pattern of brain atrophy observed in PD *in vivo*. In short, the current study is consistent with PD pathology being the result of an interaction between connectomics and local vulnerability.

The current report has some limitations. First, the cross-sectional nature of the dataset does not consider individual variability in disease progression over time. In addition, this study relied on retrospective data collection that resulted in inconsistent availability of clinical information across the contributing sites, limiting our ability to deeply phenotype our participants. For example, the Unified Parkinson’s Disease Rating Scale is the gold-standard assessment of motor symptom severity in PD [73]. Depending on the protocol at a given study site, this scale was administered when the PD patient was either on or off their dopaminergic medications, or both on and off in separate sessions, making it challenging to harmonize and interpret the resulting scores. Our analyses relied on normative models and estimates of network connectivity, gene expression, and other brain features derived from unrelated cohorts of young healthy adults to contextualize the atrophy pattern measured in PD. Relating individual patient multimodal data instead could better account for between-subject variability. Finally, structural brain estimates were parcellated using the Desikan-Killiany atlas [27] in accordance with standardized ENIGMA protocols. Future work should aim to replicate the findings reported here at multiple spatial scales.

In summary, we found widespread patterns of brain atrophy in PD that were shaped by network architecture, across disease stages, and in individual patients. Cortical abnormalities overlapped with maps of local brain features, including intrinsic networks, cytoarchitectonics, and neurotransmitter systems. The observed atrophy pattern also correlated with gene expression profiles related to synaptic structure and function. Our results demonstrate how we can contextualize the spatial pattern of atrophy using multimodal data to better understand the contribution of network spreading and local vulnerability in PD.

## METHODS

### Participants

The ENIGMA-PD working group aggregated 3D volumetric T1-weighted brain MRI and clinical data from PD patients and HCs across 23 international contributing sites (Supplementary Table S1). Brain imaging was available from 3,216 PD patients and 1,480 HCs. Individual site MRI scanning protocols and participant inclusion/exclusion criteria are detailed in Supplementary Table S2. Clinical information from PD patients included Hoehn and Yahr (HY) stage score [26], time since diagnosis (in years), and Montreal Cognitive Assessment (MoCA) score [28]. HY scores ranged from 1 (i.e., unilateral motor impairment) to 5 (i.e., confinement to bed or wheelchair). We used a modified HY classification such that intermediate scores HY 1.5 and 2.5 were regrouped into HY 2, and HY 4 and 5 were combined due to smaller group sizes. All participants provided written informed consent to their local site before participating in site-specific studies, which were approved by the respective local ethics committee and institutional review board. Anonymized imaging and clinical data were shared with the ENIGMA-PD working group.

### Structural brain morphometry in PD

Each contributing site collected and processed MRI data using a standardized FreeSurfer 5.3 pipeline [74], extracting 68 regional cortical thickness, 68 cortical surface area, 16 subcortical volume, and total intracranial volume estimates according to the Desikan-Killiany atlas [27]. These regional brain estimates were visually inspected for quality following standard ENIGMA protocols (http://enigma.usc.edu/protocols/imaging-protocols) and shared with the ENIGMA-PD investigators. Participants younger than 40 years of age or those with more than 50% missing data after quality control were excluded from the analysis. This resulted in a final sample of 3,096 PD patients and 1,262 HCs. Details regarding the participant samples and scanner protocols used at each contributing site are provided in Supplementary Table S1 and S2.

The 23 sites contributed a combined 53 cohorts, each with different cohort-specific scanning and clinical testing environments. Accordingly, we first harmonized brain estimates using ComBat, a bayesian statistical harmonization method designed to account for batch effects in multi-site MRI studies [75]. A small subset of cohorts (9 of 53 cohorts) had data available only from PD patients but no matched HCs. To rule out that our results may have been influenced by inclusion of these unmatched cohorts during data harmonization, we conducted a robustness test by comparing group average brain maps derived from data harmonized before versus after excluding PD-only cohorts from the sample. We found comparable abnormality patterns between these two samples for all measures (see Supplementary Fig. S1a and S1c). Any remaining missing values were imputed based on the mean of the group (i.e., PD or HC) and disease stage (i.e., HY score) to which a given participant belonged.

For each brain measure estimate, we generated *w*-score maps for each PD patient [76]. This procedure is analogous to *z*-scoring with the additional adjustment for age and sex covariates. Since cortical surface area and subcortical volumes scale with head size [77], total intracranial volume was also included as a covariate in their *w*-score models. Given the PD and HC groups were significantly different in terms of age and sex and the potential for this to influence *w*-score estimates, we ran a sensitivity analysis comparing group average brain maps derived from data with versus without HY stage stratified, propensity score matching with replacement using the MatchIt tool (see Supplementary Fig. S1a and S1c) [78]. We found comparable abnormality patterns between these two samples for all measures.

For the *w*-scoring procedure, linear regressions between regional brain estimates and age and sex were first performed in the HC reference cohort. Then regional *w*-scores were calculated for each PD patient using the following formula:

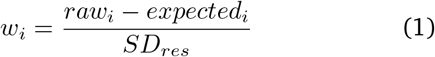

where *w*_*i*_ is the *w*-score of region *i, raw*_*i*_ is the estimate value at region *i* observed in the patient, *expected*_*i*_ is the estimate value at region *i* expected in the HC group given the patient’s age and sex (estimate ∼ age + sex), and *SD*_*residual*_ is the standard deviation of the residuals in HCs. Here, negative *w*-scores reflected lower estimates (i.e., atrophy) whereas positive *w*-scores indicated higher values in PD than expected in HCs. Group average maps of PD-related deviations for each estimate were generated from the mean regional *w*-scores across all PD patients. One-sample *t*-tests examined if group average *w*-scores in PD significantly deviated from HCs (i.e., test if average PD had a *w*-score that differed from 0). Results were corrected for multiple comparisons using the false discovery rate (FDR) method [79].

### Relationship between structural brain abnormalities and clinical measures

We tested whether maps of PD-related brain abnormalities were associated with clinical measures, controlling for age and sex. Clinical measures included HY stage, time since diagnosis (in years), and global cognition as estimated by MoCA scores (i.e., lower scores reflect poorer cognition) [28]. Given that HY scores are ordinal in nature, we used non-parametric, rank-based partial correlations that model the monotonic relationship between variables of interest. For the other continuous variables, we used linear regression models. To ensure consistent interpretation of correlations, we inverted MoCA scores to align with the directionality of the other clinical measures so that higher values reflected greater disease severity. Results were FDR-corrected for multiple comparisons separately for each brain and clinical measure [79].

### Spatial null models

The inherent autocorrelation among brain regions can artificially inflate correlations when testing the spatial overlap between two brain maps [80]. To mitigate this issue, we evaluated the statistical significance of such correlations using spatial null models, or “spin tests” [81, 82], whenever appropriate. Null models were generated using the netneurotools toolbox (https://netneurotools.readthedocs.io/en/latest/). Coordinates of cortical surface parcels were first projected onto the surface of spheres, then randomly rotated for one hemisphere and mirrored to the other. Subsequently, cortical surface data were reassigned with the values of the closest rotated parcel. This procedure was repeated 1,000 times, to construct a null distribution that preserved the spatial autocorrelation of the original surface map and provided a benchmark against which we can compare the original observation. For subcortical data, instead of spherical projection, parcels were randomly shuffled to generate permuted null models [83].

### Structural and functional MRI data

Structural and functional connectivity networks were obtained from the `enigmatoolbox` [83], derived from diffusion-weighted imaging and resting-state functional MRI data in a cohort of unrelated healthy adults from the Human Connectome Project (n = 207, 83 males, mean age ± SD = 28.74 ± 3.73 years, range = 22-36 years) [29]. Diffusion-weighted imaging data (spin-echo EPI sequence, TR = 5520 ms, TE = 89.5 ms, FOV = 210 /texttimes/ 180, voxel size = 1.25 mm^3^, b-value = 1000/2000/3000 s/mm^2^, 270 diffusion directions, 18 b0 images) underwent b0 intensity normalization and were corrected for susceptibility distortion, eddy currents, and head motion. Resting-state functional MRI data (gradient-echo EPI sequence, TR = 720 ms, TE = 33.1 ms, FOV = 208 × 180 mm^2^, voxel size = 2 mm^3^, 72 slices) were corrected for distortion, head motion, and magnetic field bias, and underwent skull removal, intensity normalization, and registration to MNI152 space [84]. Automatic removal of noise components (e.g., head motion, white matter, cardiac pulsation, arterial, and large vein-related effects) was performed using FSL FIX [85]. The resulting preprocessed time series were transformed into grey matter ordinate space using cortical ribbon-constrained volume-to-surface mapping and combined into a single time series.

### Normative connectivity network models

Structural connectivity networks from the enigmatoolbox [83] were built from preprocessed diffusion-weighted imaging data using MRtrix3 [86]. This involved performing tractography constrained to anatomically derived tissue types (i.e., cortical and subcortical grey matter, white matter, and cerebrospinal fluid) [87], estimation of multi-shell and multi-tissue response functions [88], and constrained spherical deconvolution and intensity normalization [89]. The initial tractogram was generated with 40 million stream-lines (max length = 250, fractional anisotropy cutoff = 0.06). Spherical deconvolution-informed filtering of tractograms with SIFT2 [90] was used to reconstruct whole brain streamlines weighted by cross-sectional multipliers. Individual structural connectomes were generated by mapping the reconstructed streamlines onto 68 cortical and 14 subcortical regions (the lateral ventricles were excluded). A distance-dependent thresholding procedure [91], which preserves the edge length distribution of individual connectomes, and log transformation was used to define a group-average structural connectivity network in which each connection represents the number of streamlines or fibre density between two brain regions. Functional connectivity networks were constructed by performing pairwise correlations between the time series of 68 cortical and 14 subcortical regions. Negative connections were set to zero. Individual functional connectomes were *z*-transformed and averaged across subjects to generate a group-average functional connectivity network.

Four normative network models were used in the present study: (i) cortico-cortical structural network, (ii) subcortico-cortical structural network, (iii) corticocortical functional network, and (iv) subcortico-cortical functional network. Cortico-cortical networks were based on connections among 68 cortical regions (68 × 68 matrix). Subcortico-cortical networks featured connections between 14 subcortical and 68 cortical regions (14 × 68 matrix).

### Network-based disease exposure

We examined how the patterns of PD-related atrophy are shaped by structural and functional connectivity across PD disease stages (Fig. 2a). Specifically, we tested if the atrophy measured at a given region (or node) was related to atrophy across its structurally and functionally connected neighbours [43, 44]. Nodal atrophy refers to the regional *w*-scores from the cortical thickness and subcortical volume maps, which showed the most sensitivity to PD-related atrophy and clinical features (Fig. 1a-b). Neighbourhood atrophy of a given node was defined as the average *w*-score across its structurally connected neighbour regions weighted by the strength of connectivity, described in the following formula:

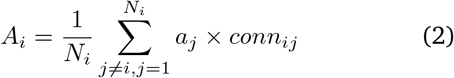

where *A*_*i*_ is the neighbourhood atrophy of node *i, a*_*j*_ is the atrophy of the *j*-th neighbour of node *i, conn*_*ij*_ is the strength of the connection between nodes *i* and *j*, and *N*_*i*_ is the total number of structurally connected neighbours to node *i* (i.e., node degree). Note that neighbourhood atrophy is normalized by the node degree (*N*_*i*_) and is thus made independent of nodal atrophy. Self-connections between a node and itself are also excluded (*j* ≠ *i*).

For each network model, we tested the relationship between node and neighbourhood atrophy using Spearman rank correlations. Each test was compared against spatial null models to determine statistical significance. Finally, we repeated this analysis separately on the group average, disease stage, and single-subject atrophy maps.

### Network-based epicentre likelihoods

For each network model, we ranked brain regions based on their nodal atrophy and neighbourhood atrophy (Fig. 2a). The epicentre likelihood of a given region was defined as the average of node and neighbourhood atrophy ranks, so that those regions with greater atrophy that are also connected to neighbourhoods with greater atrophy are considered more likely epicentres [43, 44]. These likelihoods were tested against spatial null models for statistical significance. It is important to note that a network-based epicentre does not necessarily implicate a region as the initiation site of disease. Instead, the term describes a region with an atrophy and connectivity profile ideal for a propagator of disease pathology.

Epicentre likelihoods were examined separately for the group average, disease stage, and single-subject atrophy maps. For disease stage atrophy maps, after identifying regions with significant epicentre likelihoods for each HY stage, we explored the convergence of these epicentre regions across the four disease stages. For the single-subject atrophy maps, we similarly looked at the convergence of significant epicentre regions across individual PD patients. Anticipating a large degree of heterogeneity between individualized epicentre maps, epicentre regions were also mapped to intrinsic brain networks [30] to test if they might co-localize to common circuitry.

### Spatial overlap between atrophy and brain annotations

We investigated the spatial correspondence between PD-related atrophy and annotations of discrete brain systems, including intrinsic networks [30] and cytoarchitectonic tissue classes [31, 32]. The Yeo intrinsic networks [30] classify cortical regions into seven distinct resting-state networks: default mode, frontoparietal, limbic, visual attention, dorsal attention, sensorimotor, and visual networks. Similarly, the von Economo classification [31, 32] assigns cortical regions into seven cytoarchitectonic types: insular, limbic, primary sensory, primary/secondary sensory, association 1 and 2, and primary motor classes. For cortical thickness and surface area maps, we computed the mean w-score within each network or tissue class to localize the macroscale distribution of atrophy in PD. Observed mean atrophy within each system was compared against null mean distributions derived from spatial null models.

We also explored the spatial overlap between PD-related atrophy and 18 receptor and transporter densities that span nine neurotransmitter systems [33, 34]. Volumetric radiotracer maps from a total cohort of over 1,200 healthy individuals were obtained from https://github.com/netneurolab/hansen_receptors. This included data for acetylcholine (*α*_4_*β*_2_, M_1_, VAChT), cannabinoid (CB_1_), dopamine (D_1_, D_2_, DAT), GABA (GABA_*A/BZ*_, histamine (H_3_), glutamate (mGluR_5_, NMDA), norepinephrine (NET), opioid (MOR), and serotonin (5-HT1_*A*_, 5-HT1_*B*_, 5-HT2_*A*_, 5-HT_4_, 5-HT_6_, 5-HTT). These tracer maps were parcellated according to the Desikan-Killiany atlas and individually *z*-scored using the neuromaps toolbox [34]. For tracers with multiple available maps, we combined them into a single map using weighted averaging. Spearman rank correlations assessed the relationship between each cortical thickness, cortical surface area, and subcortical volume map and each neurotransmitter map. Correlations were tested against spatial null models and FDR-corrected for multiple comparisons.

### Gene expression profiles

Gene expression data were obtained from the Allen Human Brain Atlas [35] and processed with the abagen toolbox [92]. The dataset is composed of microarray data derived from six post-mortem brains (5 males, 1 female; mean age ± SD = 42.5 ± 13.4 years). Probes were reannotated following previous recommendations [93]. Those with a signal-to-noise ratio greater than 50% were retained. If there were multiple probes of the same gene, the one with the most consistent pattern of regional expression across donors was selected. This procedure resulted in a total of 15,633 genes retained. Samples were assigned to parcels of the Desikan-Killiany atlas [27]. This sample-to-parcel matching was restricted to each hemisphere and within gross structural divisions to minimize assignment errors. When a probe was not found directly within a parcel, the nearest sample up to 2 mm away was selected. If no probes were found within 2 mm of a parcel, then the sample closest to the centroid of the parcel across all donors was chosen. Samples unable to be assigned to a parcel were discarded. Expression values were normalized across genes using a scaled robust sigmoid function and rescaled to the unit interval, then normalized across tissue samples using the same procedure. Regional gene expression profiles were obtained by first averaging across probes belonging to the same parcels separately for each donor, then averaging across donors. Given that data from the right hemisphere were available from only two of the six donors, the current analysis was restricted to the left hemisphere. In addition, due to the large transcriptional difference between the cortex and subcortex [94] and relatively few subcortical regions to assess (i.e., seven parcels), we restricted our analysis to the cortex. Regional gene expression profiles were averaged across donors to construct a 34 region × 15,633 gene expression matrix that was used for partial least squares analysis.

### Partial least squares analysis

We used partial least squares (PLS) analysis [95, 96] to identify the patterns of gene expression associated with PD-related atrophy (Fig. 4a). This approach identifies latent variables that explain the maximum covariance between matrices *X* (gene expression: 34 regions × 15,633 genes) and *Y* (atrophy: 34 regions). The statistical significance of latent variables was assessed against the variance observed in 1,000 spatial null models. For each significant latent variable identified, the contribution of individual genes was determined through bootstrap resampling. This procedure involves shuffling matrices *X* and *Y*, then repeating the PLS analysis 1,000 times to generate a null distribution, and using the standard errors to estimate the weight (or contribution) of each gene. Bootstrap ratios, which are interpreted in the same way as *z*-scores, were calculated as the ratio of each gene weight to its bootstrap-estimated standard error. Genes with larger bootstrap ratios contribute more significantly and reliably to a given latent variable. Ranked gene lists based on these bootstrap ratios were submitted to gene set enrichment analysis [97].

### Gene set enrichment analysis

To investigate the biological relevance of gene expression correlates of PD-related atrophy, we performed gene set enrichment analysis (GSEA) using the WebGestalt platform (https://www.webgestalt.org) and the Gene Ontology knowledge base (https://geneontology.org). GSEA tests whether the most positively and negatively weighted genes in a ranked gene list, derived here from bootstrap resampling, occur more frequently than expected by random chance and identifies the biological process and cellular component terms associated with these significant genes. The minimum and maximum number of genes for enrichment was set to 3 and 2,000, respectively. Results were adjusted by running 1,000 random permutations, followed by FDR correction for multiple comparisons. We report and interpret the 10 most positively and negatively weighted terms.

## Data availability

Publicly available datasets used in this report include the Parkinson Progression Marker Initiative (PPMI; https://ppmi-info.org), OpenNeuro Japan including Udall cohort (https://openneuro.org/datasets/ds000245), and Neurocon and Tao Wu’s datasets (https://fcon_1000.projects.nitrc.org/indi/retro/parkinsons.html). The PPMI – a public-private partnership – is funded by the Michael J. Fox Foundation for Parkinson’s Research and funding partners, including 4D Pharma, Abbvie, AcureX, Allergan, Amathus Therapeutics, Aligning Science Across Parkinson’s, AskBio, Avid Radiopharmaceuticals, BIAL, BioArctic, Biogen, Biohaven, BioLegend, BlueRock Therapeutics, Bristol-Myers Squibb, Calico Labs, Capsida Biotherapeutics, Celgene, Cerevel Therapeutics, Coave Therapeutics, DaCapo Brainscience, Denali, Edmond J. Safra Foundation, Eli Lilly, Gain Therapeutics, GE HealthCare, Genentech, GSK, Golub Capital, Handl Therapeutics, Insitro, Jazz Pharmaceuticals, Johnson Johnson Innovative Medicine, Lundbeck, Merck, Meso Scale Discovery, Mission Therapeutics, Neurocrine Biosciences, Neuron23, Neuropore, Pfizer, Piramal, Prevail Therapeutics, Roche, Sanofi, Servier, Sun Pharma Advanced Research Company, Takeda, Teva, UCB, Vanqua Bio, Verily, Voyager. Individual ENIGMA-PD sites retain ownership of their MRI data and only share anonymized derived data for analysis. Data are therefore not openly available but researchers are invited to join the ENIGMA-PD working group where they can formally request derived data via secondary proposals. Data requests are then considered by the individual site’s principal investigator(s). If you are interested in joining the ENIGMA-PD, please contact. For more information, please see the working group website: https://enigma.ini.usc.edu/ongoing/enigma-parkinsons/.

Tools for mapping cortical parcellations to network and cytoarchitectonic partitions and for generating spatial null models are available as part of netneurotools available at https://netneurotools.readthedocs.io/en/latest/. Structural and functional cortico-cortical and subcortico-cortical connectivity matrices are available as part of the enigmatoolbox available at https://enigma-toolbox.readthedocs.io/en/latest/. Volumetric radiotracer maps of neurotransmitter receptors and transporters are available at https://github.com/netneurolab/hansen_receptors. Post-mortem gene expression data from the Allen Human Brain Atlas are available as part of the abagen toolbox available at https://abagen.readthedocs.io/en/stable/.

## Code availability

All code used to perform the analyses are available from the corresponding author upon reasonable request.

## Acknowledgments

A.V. was supported by a postdoctoral fellowship from Fonds de recherche du Quebec–Santé. F.C. is funded by a São Paulo Research Foundation (FAPESP) Grant 2013/07559-3. J.D. is funded by an ENIGMA-PD R01 grant. J.C.D-A. is funded by the Health Research Council of New Zealand (20/538), Marsden Fund New Zealand (UOC2105), Neurological Foundation of New Zealand (2232 PRG), and the Research and Education Trust Pacific Radiology (MRIJDA). M.H. received funding/grant support from Parkinson’s UK, Oxford NIHR BRC, University of Oxford, CPT, Lab10X, NIHR, Michael J Fox Foundation, European Platform for Neurodegenerative Disorders (EPND; H2020), GE Healthcare, and the PSP Association. N.J. is funded by research grant RF1NS136995. J.C.K. acknowledges support from the National Institute for Health and Care Research (NIHR) Oxford Health Clinical Research Facility, and the NIHR Oxford Biomedical Research Centre (BRC). The Centre for Integrative Neuroimaging was supported by core funding from the Wellcome Trust (203139/Z/16/Z and 203139/A/16/Z). T.R.M. is funded by a Health Research Council grant (20/538). K.L.P. is funded by R01 NS115114, K23 NS075097, P50 AG047366, and The Michael J Fox Foundation for Parkinson’s Research.

## Author contributions

A.V., C.T., S.R., and A.D. conceived the study and wrote the manuscript, with valuable revision from all authors. A.V. performed the formal analysis, with contribution from C.T., S.R., B.M., and A.D. A.V. interpreted the results with contribution from C.T., S.R., E.d’A., N.J., M.A.L., C.O-W., P.M.T., Y.D.v.d.W., E.M.v.H., and A.D. S.A-B., H.W.B., J.K.B., F.C., J.C.D-A., I.D., M.F.D., J.D., G.G., R.C.H., M.H., M.E.J., J.C.K., C.T.M., T.R.M., P.M., L.M.P., C.P., F.P., K.L.P., M.R., C.R., P.S., M.S., D.T., C-C.T., T.D.v.B., O.A.v.d.H., C.V., J-J.W., R.W., C.Y., and A.D. provided the data. A.D. was the project administrator.

## Author information

^1^Department of Neurology and Neurosurgery, Montreal Neurological Institute, McGill University, Montreal, Canada; ^2^Department of Medicine, Centre for Advanced Research in Sleep Medicine, University of Montreal, Montreal, Canada; ^3^Faculty of Health and Medicine, Lancaster University, Lancaster, UK; ^4^Division of Neuroscience and Experimental Psychology, Faculty of Biology, Medicine and Health, The University of Manchester, Manchester, UK; ^5^Department of Neurology, Amsterdam UMC, Vrije Universiteit Amsterdam, Amsterdam, The Netherlands; ^6^Amsterdam Neuroscience, Neurodegeneration, Amsterdam, The Netherlands; ^7^Social, Genetic and Developmental Psychiatry Centre, Institute of Psychiatry, Psychology and Neuroscience, King’s College London, London, UK; ^8^Department of Neurology, University of Campinas–UNICAMP, Campinas, Brazil; ^9^Department of Anatomy and Neurosciences, Amsterdam UMC, Vrije Universiteit Amsterdam, Amsterdam, The Netherlands; ^10^Te Kura Mahi ā-Hirikapo | School of Psychology, Speech and Hearing, University of Canterbury, Christchurch, New Zealand; ^11^New Zealand Brain Research Institute, Christchurch, New Zealand; ^12^Department of Neurology, Inselspital, University of Bern, Bern, Switzerland; ^13^Department of Neurology and Center of Expertise for Parkinson & Movement Disorders, Donders Institute for Brain, Cognition and Behaviour, Radboud University Medical Center, Nijmegen, The Netherlands; ^14^Centre for Cognitive Neuroimaging, Donders Institute for Brain, Cognition and Behaviour, Radboud University, Nijmegen, The Netherlands; ^15^Department of Radiology and Medical Imaging, University of Virginia, Charlottesville, VA, USA; ^16^MoVeRe group, GIGA-CRC in vivo imaging, University of Liège, Liège, Belgium; ^17^Department of Neurology, CHU Liège, Liège, Belgium; ^18^Division of Clinical Neurology, Department of Clinical Neurosciences, Oxford Parkinson’s Disease Centre, Nuffield, University of Oxford, Oxford, UK; ^19^Imaging Genetics Center, Mark and Mary Stevens Neuroimaging and Informatics Institute, Keck School of Medicine, University of Southern California, Marina del Rey, CA, USA; ^20^Division of Clinical Neurology, Department of Clinical Neurosciences; Oxford Centre for Integrative Neuroimaging, Oxford Parkinson’s Disease Centre, Nuffield, University of Oxford, Oxford, UK; ^21^Department of Neurology, Perelman School of Medicine, University of Pennsylvania, University of Pennsylvania Perelman School of Medicine, Philadelphia, PA, USA; ^22^Department of Medicine, University of Otago, Christchurch, New Zealand; ^23^Pacific Radiology Canterbury, Christchurch, New Zealand; ^24^QIMR Berghofer Medical Research Institute, Queensland, Australia; ^25^Division of Psychology, Communication and Human Neuroscience, School of Health Sciences, Faculty of Biology, Medicine and Health, The University of Manchester, Manchester, UK; ^26^Geoffrey Jefferson Brain Research Centre, Manchester Academic Health Science Centre, Northern Care Alliance & University of Manchester, Manchester, UK; ^27^Laboratory of Neuropsychiatry, IRCCS Santa Lucia Foundation, Rome, Italy; ^28^Department of Neurology & Neurological Sciences, Movement Disorders, Stanford University, Palo Alto, CA, USA; ^29^Excellence Interdepartmental Center for Advanced MR Techniques and Department of Neurosciences, Neurology Unit, Parkinson’s Disease Center, Fondazione Cà Granda, IRCCS, Ospedale Policlinico, University of Milan, Milan, Italy; ^30^Support Center for Advanced Neuroimaging (SCAN), University Institute of Diagnostic and Interventional Neuroradiology, University Hospital Bern, Bern, Switzerland; ^31^Department of Neurology, Medical University of Graz, Graz, Austria; ^32^Department of Radiology and Biomedical Imaging, University of California San Francisco, San Francisco, CA, USA; ^33^Healthy Ageing Research Center, Chang Gung University, Taoyuan City; ^34^Department of Psychiatry, Amsterdam UMC, Vrije Universiteit Amsterdam, Amsterdam, The Netherlands; ^35^Amsterdam Neuroscience, Brain Imaging, Amsterdam, The Netherlands; ^36^Department of Medical Imaging and Radiological Sciences, Chang Gung University, Taoyuan City, Taiwan; ^37^Brazilian Institute of Neuroscience and Neurotechnology, Campinas, Brazil

---

**Figure S1:**
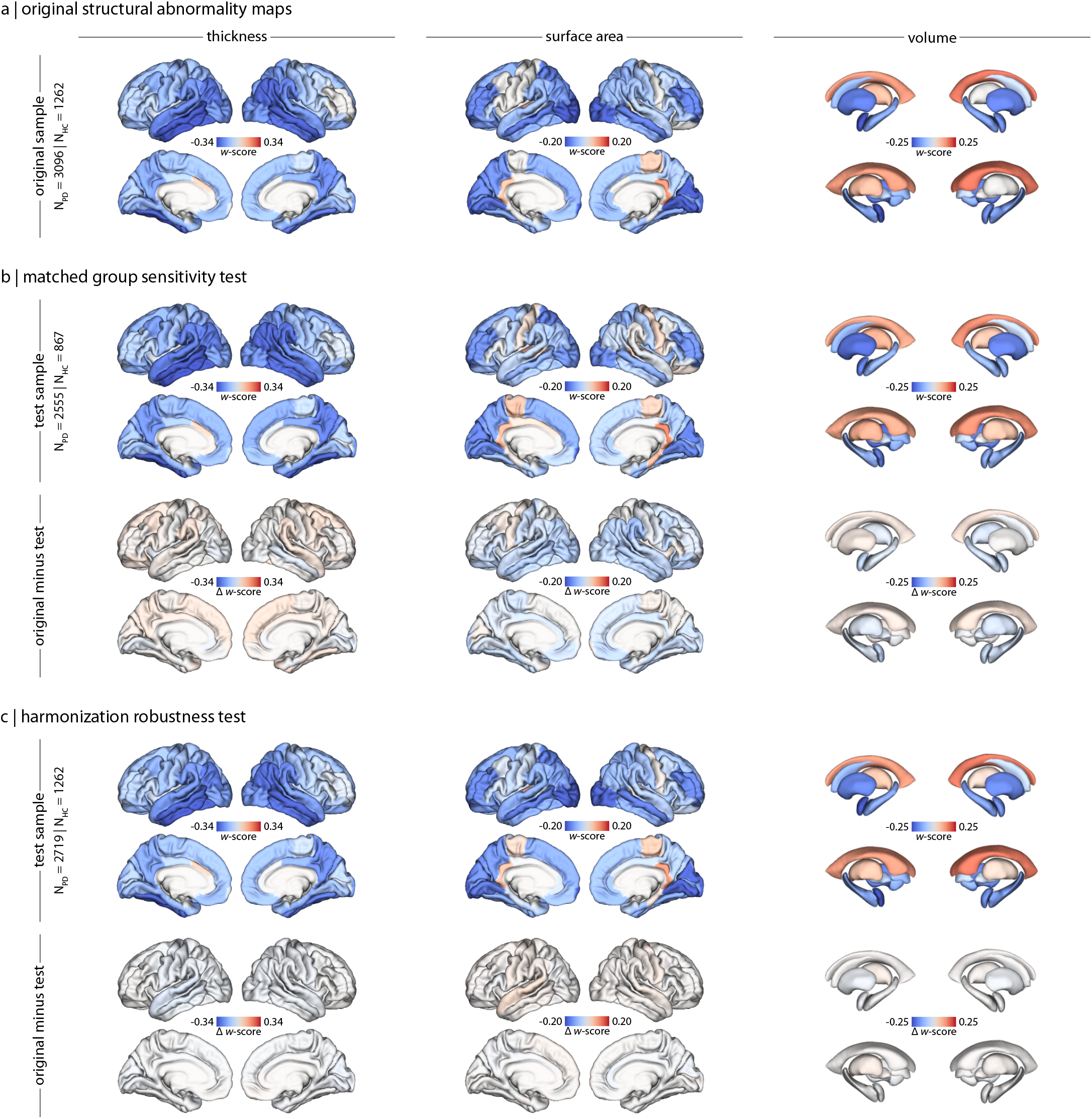
*W*-score maps of cortical thickness, cortical surface area, and subcortical volume from **(a)** the original study sample, **(b)** a test subsample after HY stage stratified, propensity score matching with replacement before computing *w*-scores, and **(c)** a test subsample after cohorts comprised of only PD patients but no HCs were removed before harmonization. *W*-score difference maps between the original study sample and **(b)** the group-match test sample or **(c)** the harmonization test subsample.

**Figure S2:**
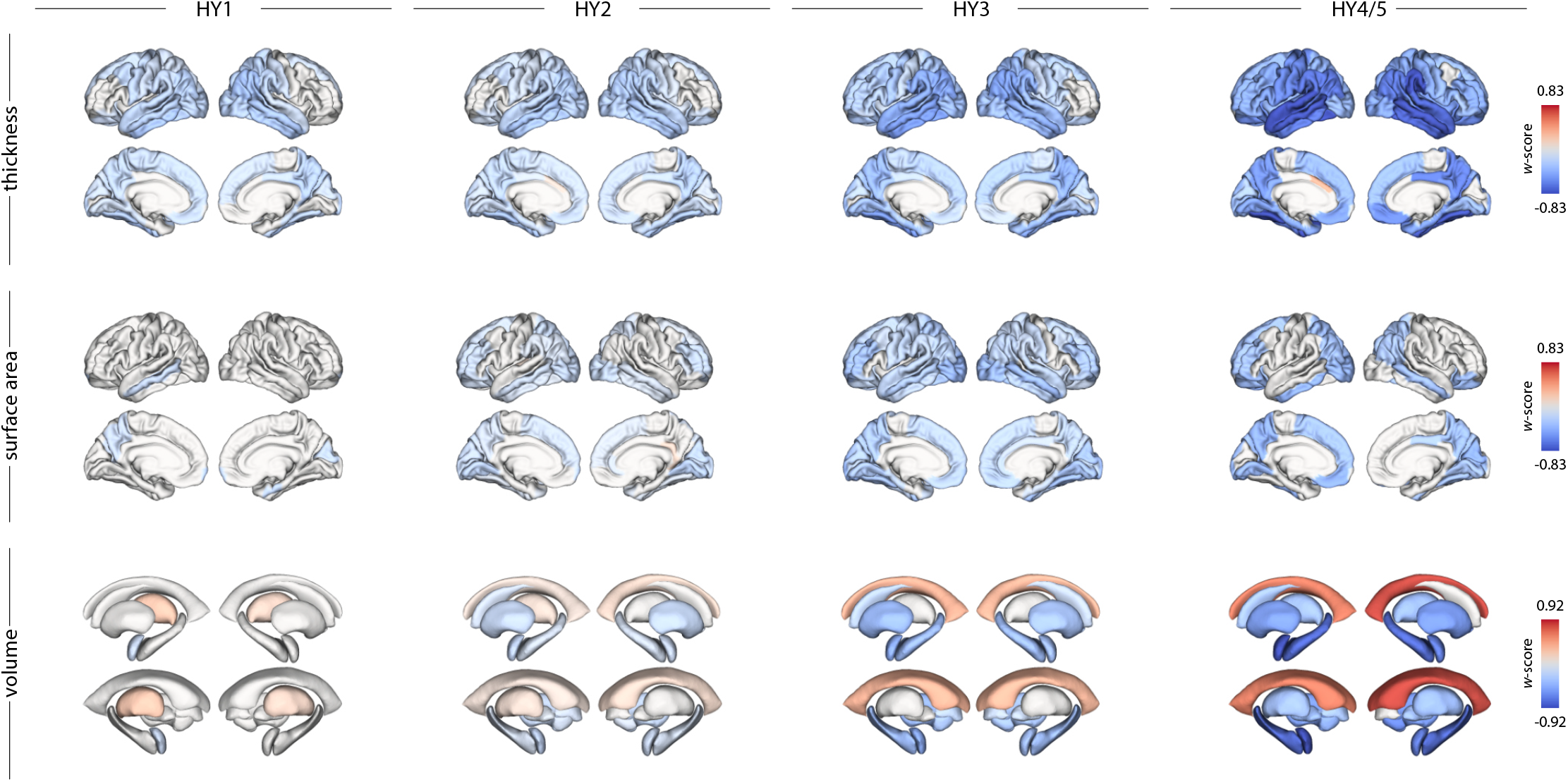
Brain maps of *w*-scores for cortical thickness (*top row*), surface area (*middle row*), and subcortical volume (*bottom row*) across Hoehn and Yahr disease stages (*left to right*). *W*-scores are plotted using a shared colour scale for each brain measure to allow visual comparisons across disease stages. Only regions surviving false discovery rate correction for multiple comparisons (*P*_*FDR*_ < 0.05) are shown.

**Figure S3:**
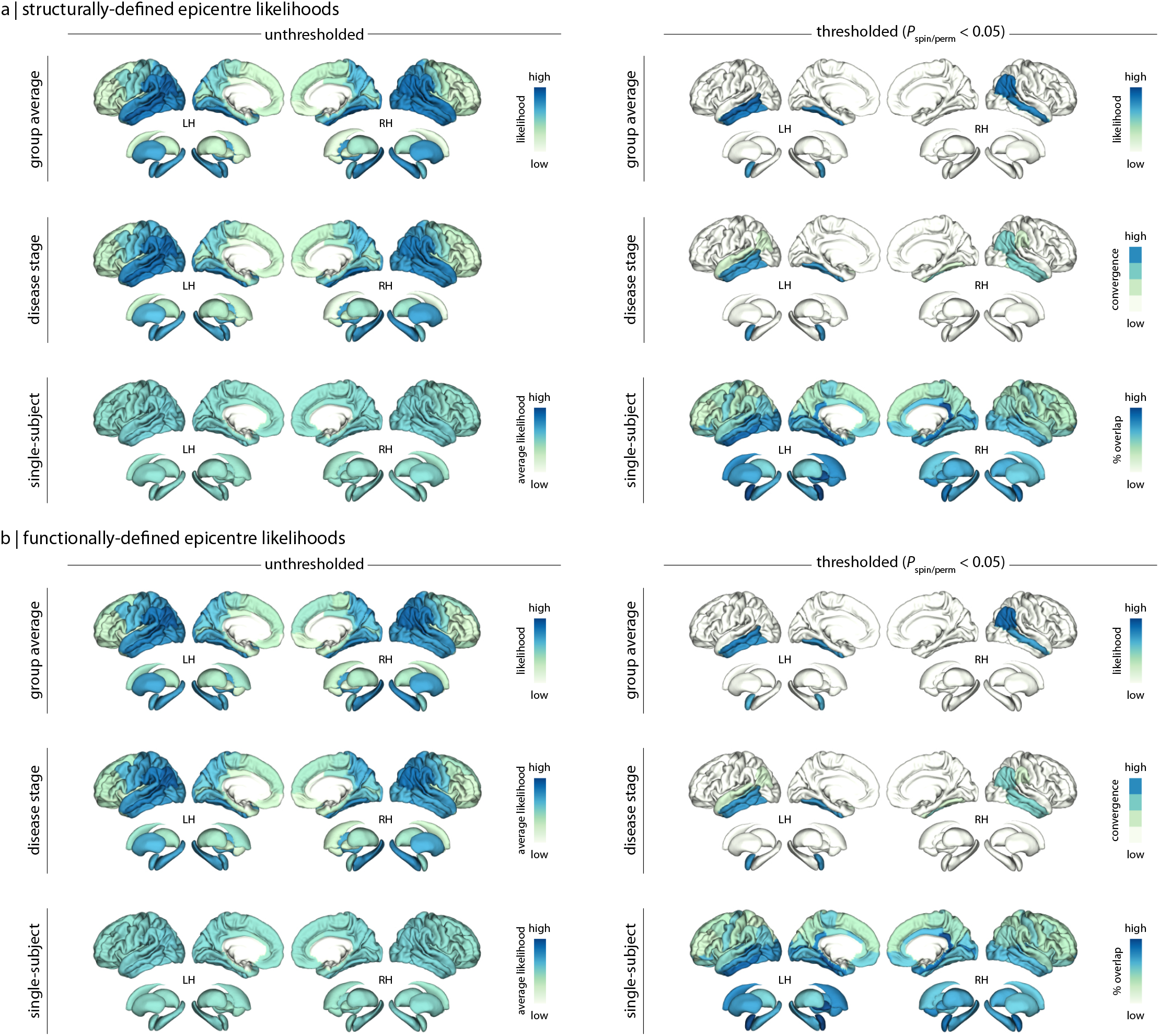
Brain maps of **(a)** structurally- and **(b)** functionally-defined epicentre likelihoods. Group average level (*top rows*) displays epicentre likelihood ranks (*left*) and significant epicentres (*right*; *P*_*spin/perm*_ < 0.05) for the group average atrophy map. Disease stage level (*middle rows*) shows average epicentre likelihood ranks (*left*) and convergence of significant epicentres (*P*_*spin/perm*_ < 0.05) across the four Hoehn and Yahr disease stages. Single-subject level (*bottom rows*) shows average epicentre likelihood ranks (*left*) and convergence of significant epicentres (*P*_*spin/perm*_ < 0.05) across individual atrophy maps.

**Figure S4:**
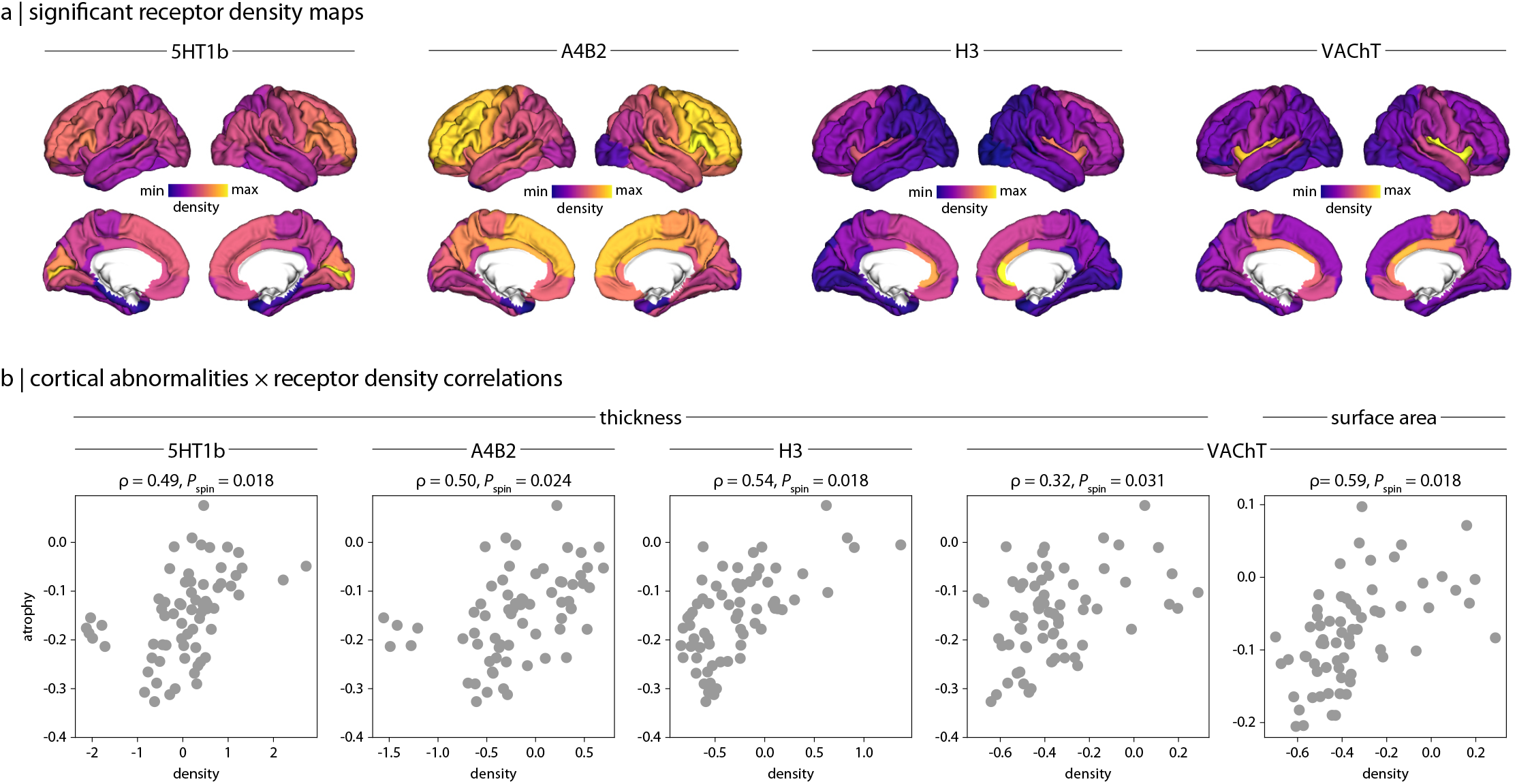
**(a)** Surface maps of neurotransmitter receptor densities significantly correlated with the pattern of cortical atrophy in PD. **(b)** Scatter plots of significant correlations between neurotransmitter receptor densities and cortical atrophy. 5-HT1_*B*_ = serotonin; *α*_4_*β*_2_ = nicotinic acetylcholine receptor; H_3_ = histamine; VAChT = vesicular acetylcholine transporter

**TABLE S1:**
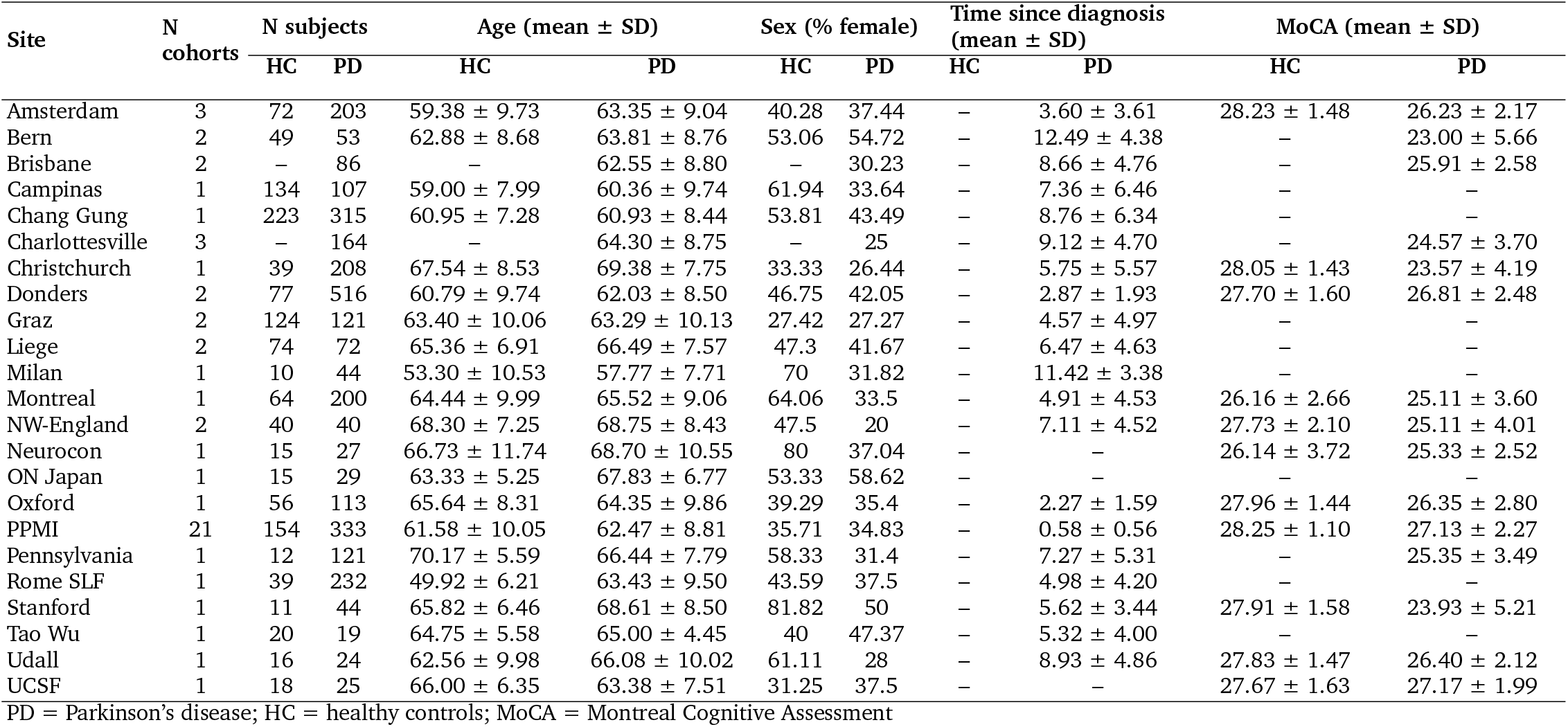
Demographic and clinical details of each ENIGMA-PD contributing site.

**TABLE S2:**
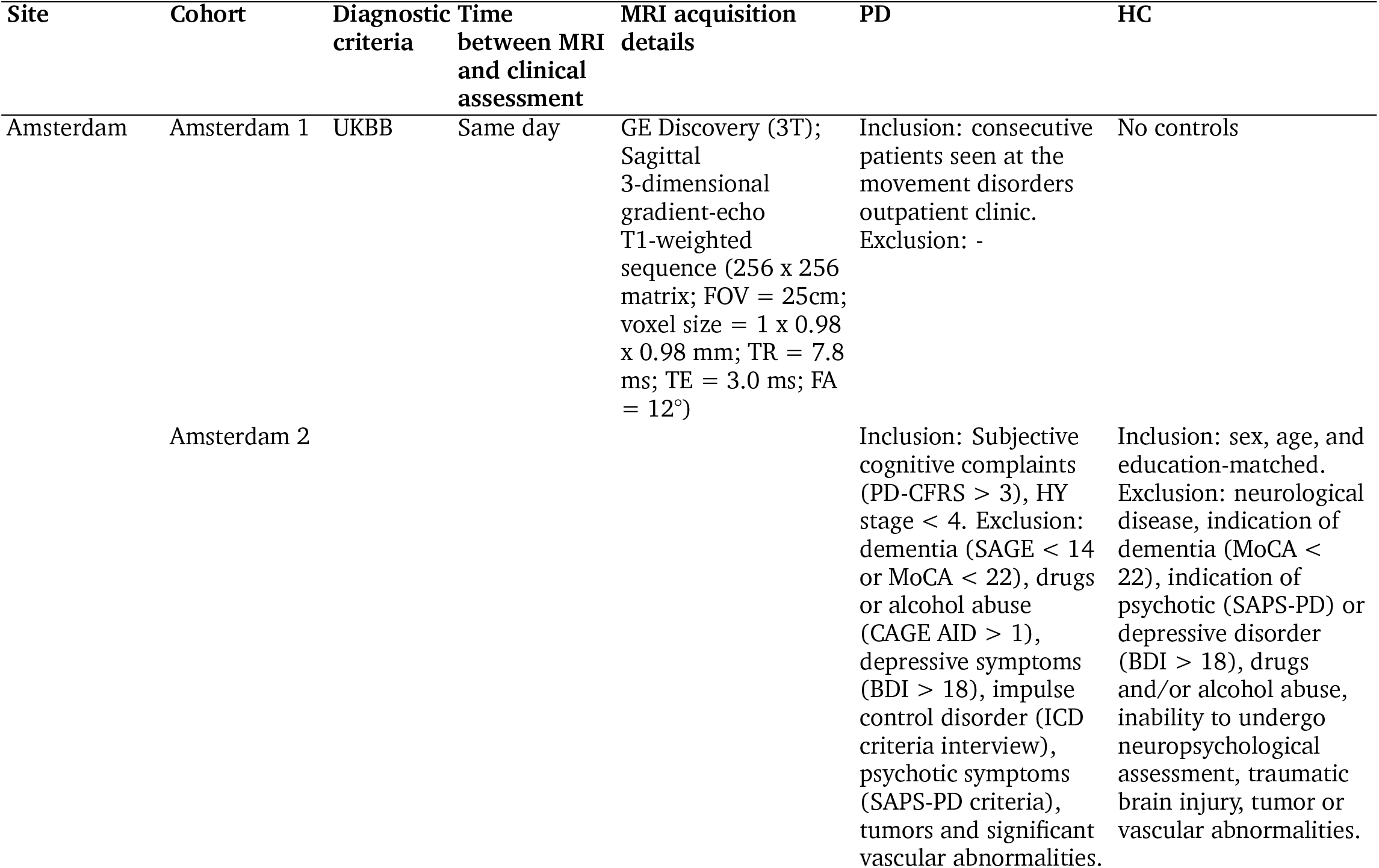
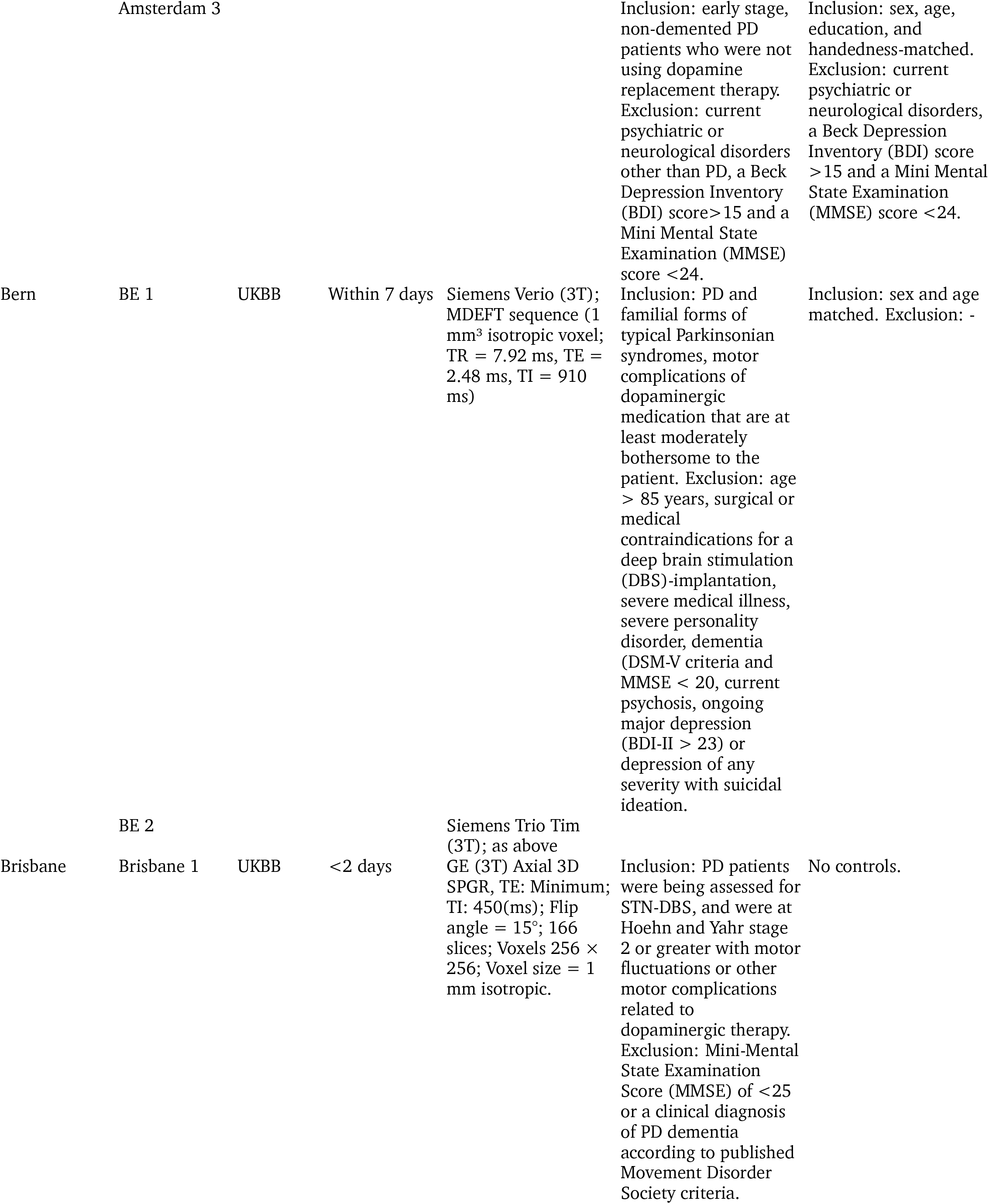
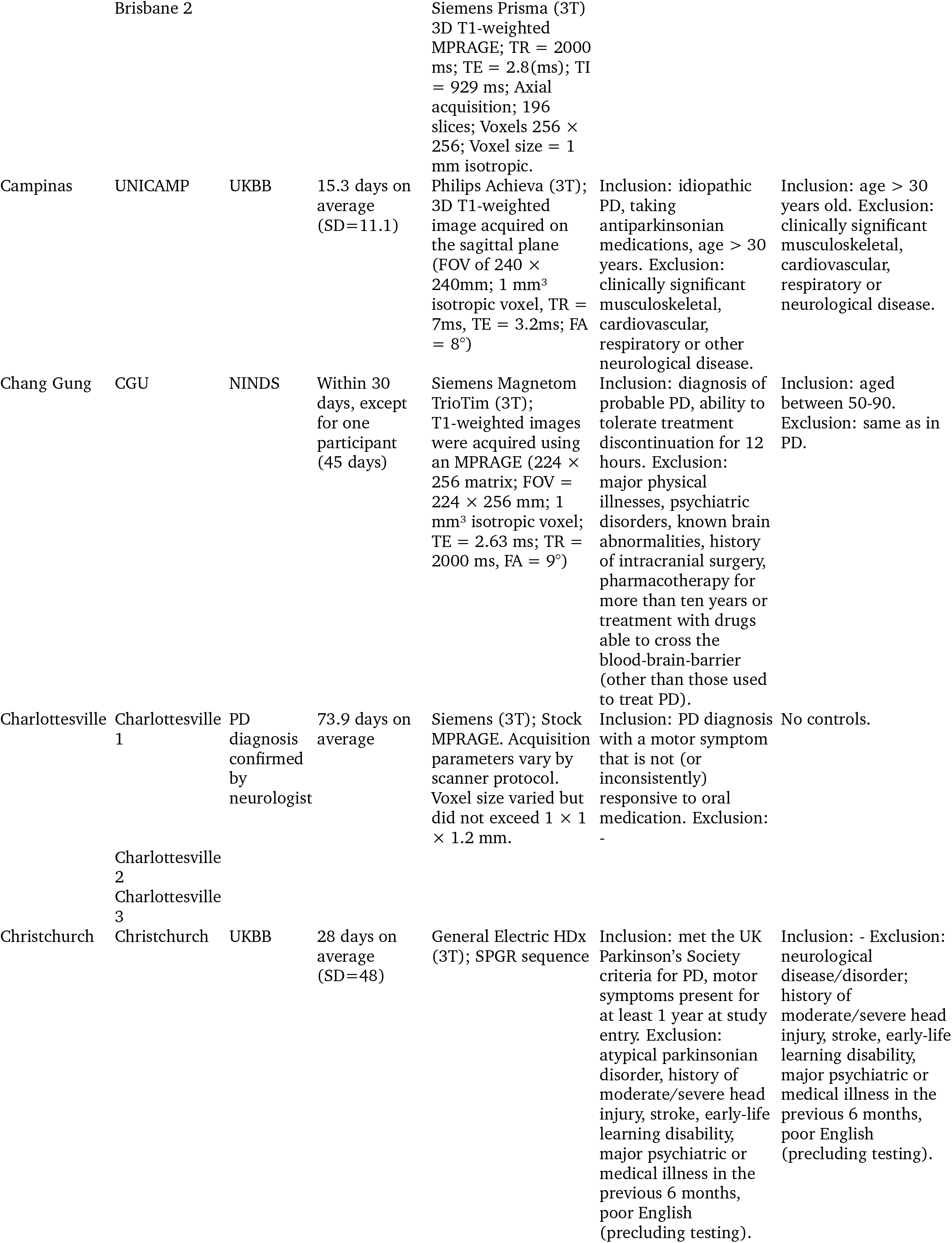
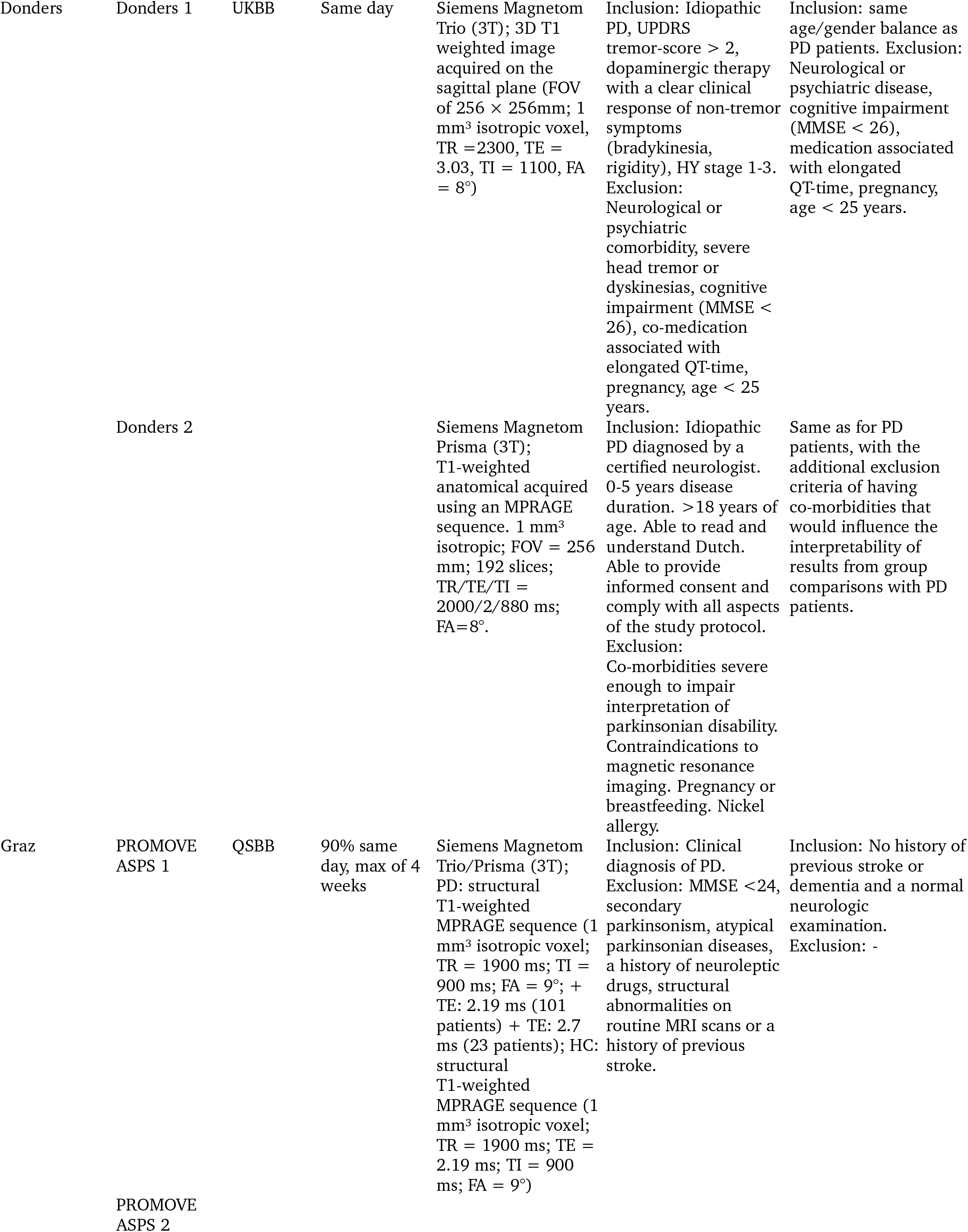
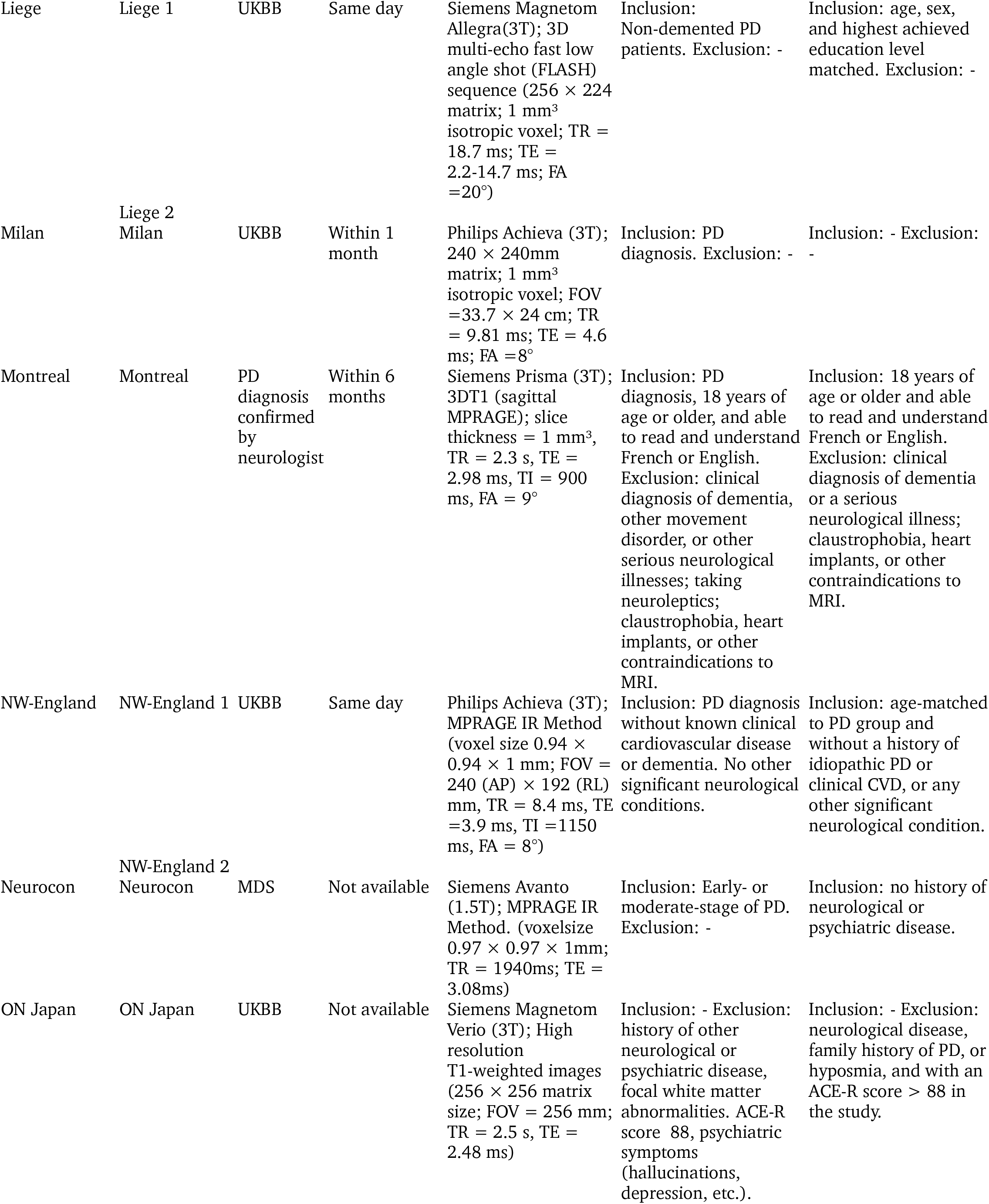
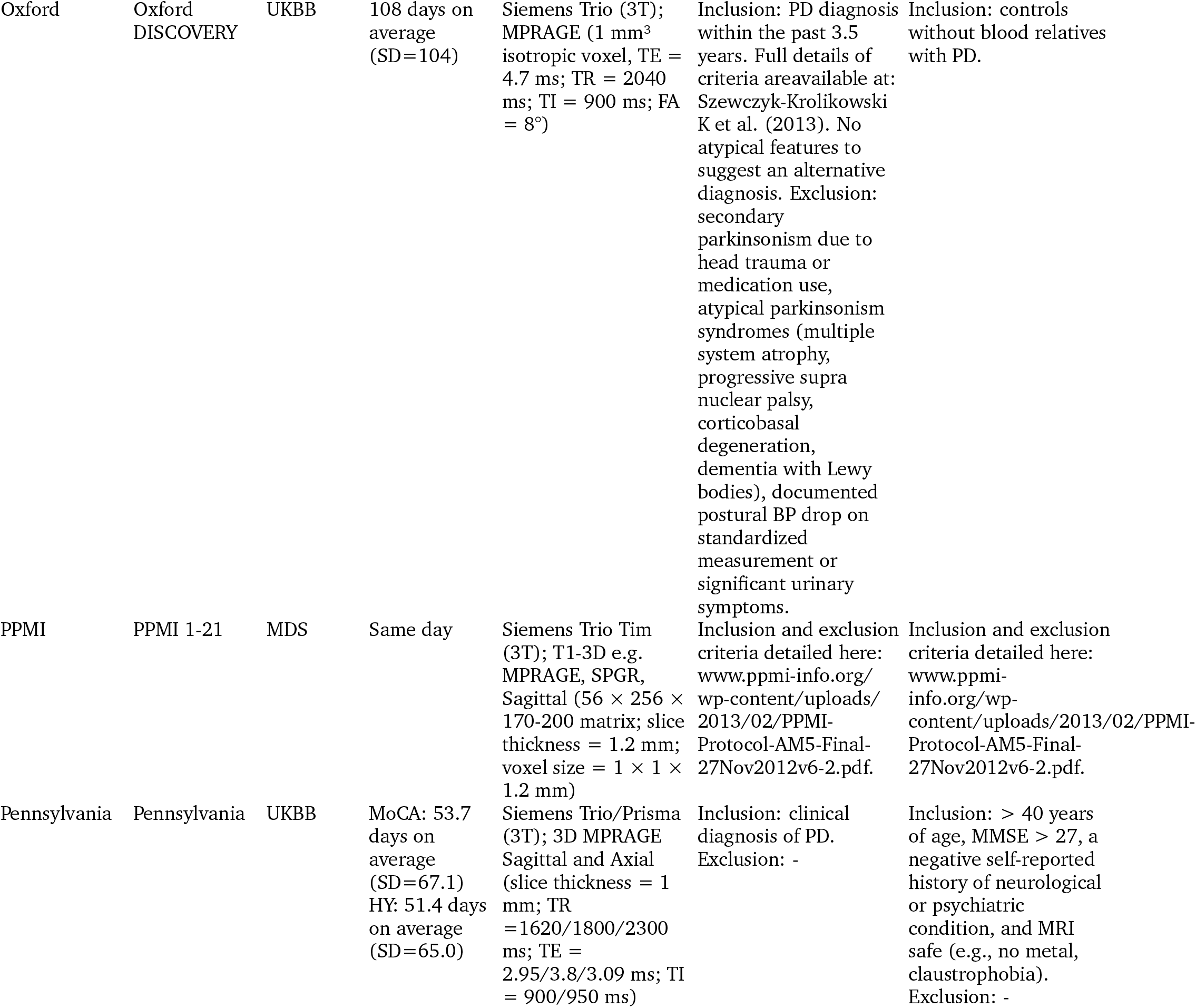
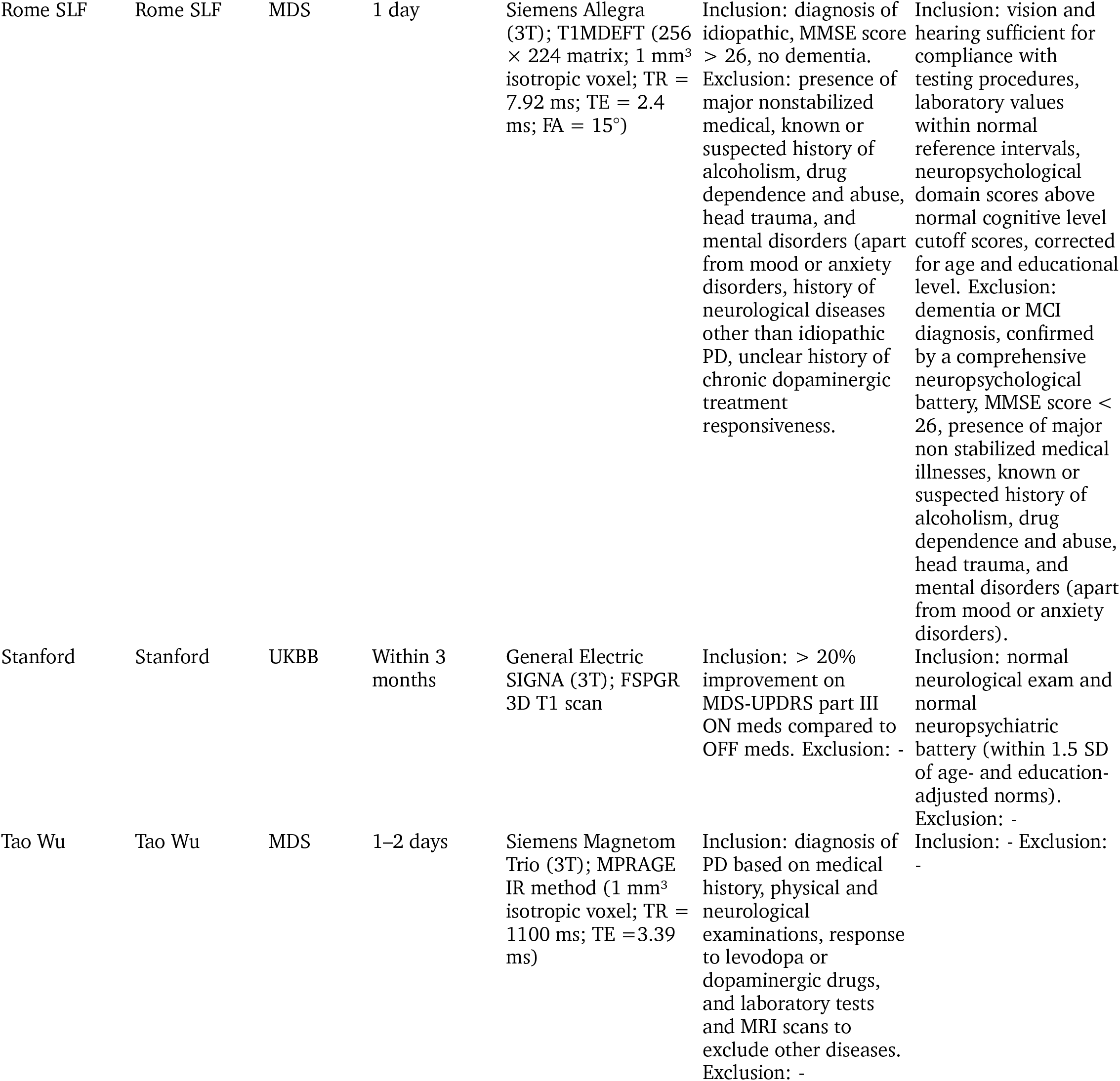
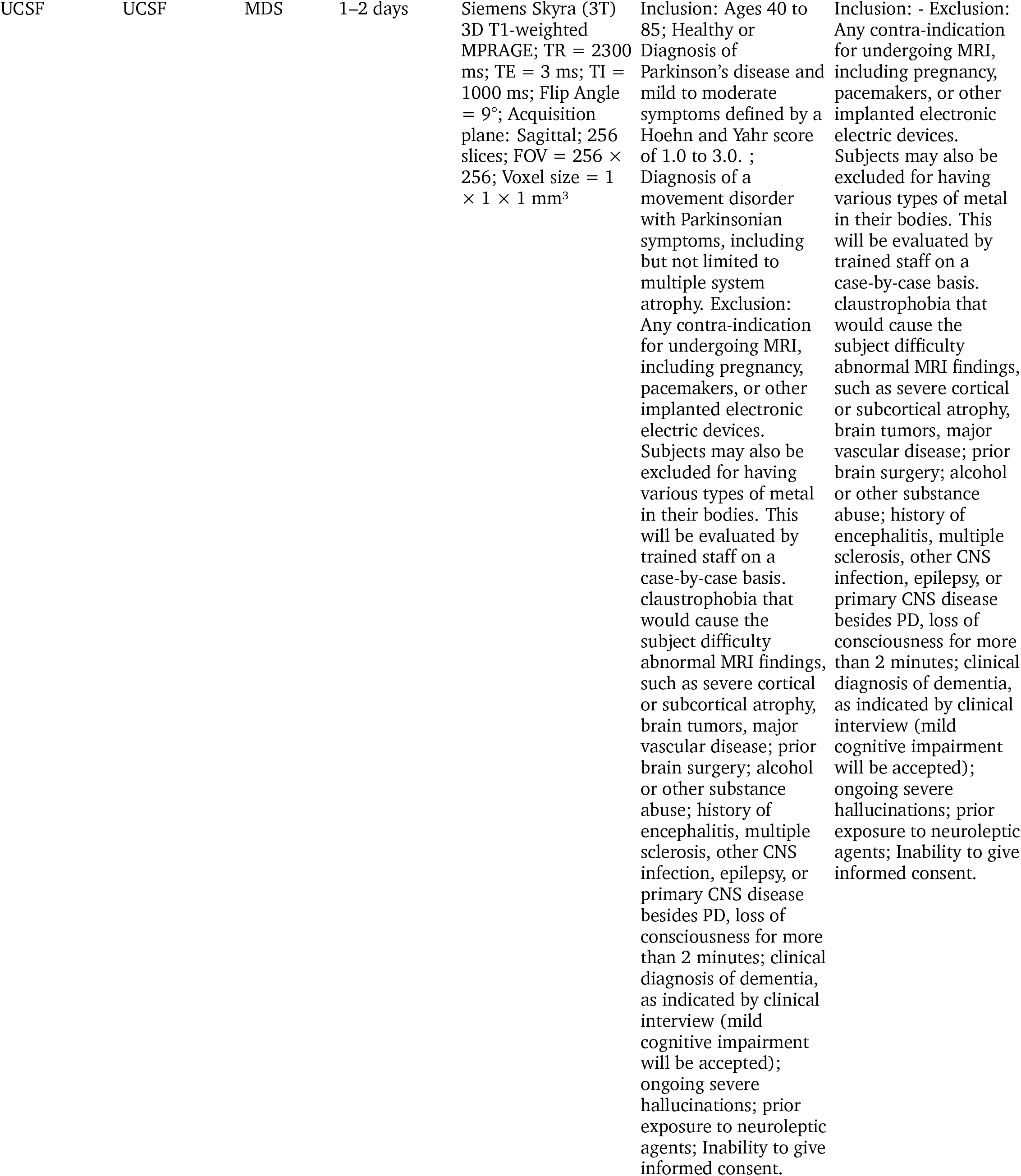
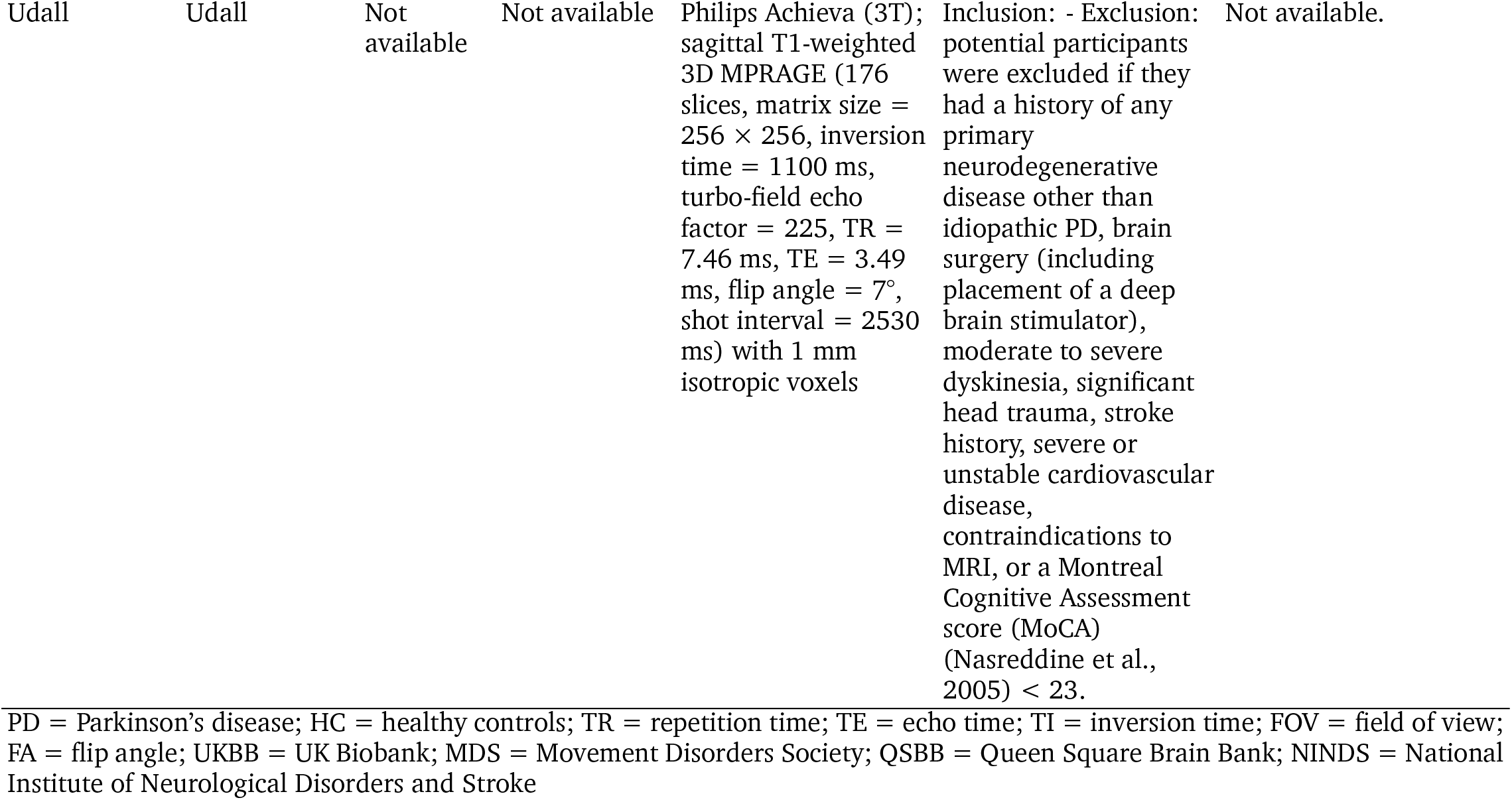
MRI scanning protocols and participant inclusion/exclusion criteria of each contributing site.

**TABLE S3:** Demographics of the age- and sex-matched subsample.

|  | PD | HC | SMD |
| --- | --- | --- | --- |
| N | 2555 | 867 | - |
| Age (mean ± SD) | 63.69 (9.09) | 62.94 (9.33) | 0.08 |
| Sex (% female) | 0.37 | 0.43 | 0.12 |

PD = Parkinson's disease; HC = healthy controls; SD = standard deviation; SMD = standard mean difference

**TABLE S4a:** One-sample t-test of cortical thickness w-score estimates.

| Region | Mean | SD | T-stat | P Value | $P_{FDR}$ |
| --- | --- | --- | --- | --- | --- |
| left bankssts | -0.259 | 1.042 | -13.858 | <0.001 | <0.001 |
| left caudalanteriorcingulate | 0.071 | 1.030 | 3.827 | <0.001 | <0.001 |
| left caudalmiddlefrontal | -0.179 | 1.074 | -9.291 | <0.001 | <0.001 |
| left cuneus | -0.113 | 1.012 | -6.203 | <0.001 | <0.001 |
| left entorhinal | -0.170 | 1.052 | -9.006 | <0.001 | <0.001 |
| left fusiform | -0.314 | 1.089 | -16.039 | <0.001 | <0.001 |
| left inferiorparietal | -0.282 | 1.090 | -14.410 | <0.001 | <0.001 |
| left inferiortemporal | -0.336 | 1.086 | -17.227 | <0.001 | <0.001 |
| left isthmuscingulate | -0.217 | 1.010 | -11.978 | <0.001 | <0.001 |
| left lateraloccipital | -0.214 | 1.033 | -11.514 | <0.001 | <0.001 |
| left lateralorbitofrontal | -0.142 | 1.088 | -7.248 | <0.001 | <0.001 |
| left lingual | -0.156 | 1.018 | -8.550 | <0.001 | <0.001 |
| left medialorbitofrontal | -0.155 | 1.042 | -8.275 | <0.001 | <0.001 |
| left middletemporal | -0.327 | 1.034 | -17.605 | <0.001 | <0.001 |
| left parahippocampal | -0.183 | 1.020 | -9.999 | <0.001 | <0.001 |
| left paracentral | -0.143 | 1.149 | -6.941 | <0.001 | <0.001 |
| left parsopercularis | -0.073 | 1.077 | -3.756 | <0.001 | <0.001 |
| left parsorbitalis | -0.133 | 1.022 | -7.231 | <0.001 | <0.001 |
| left parstriangularis | -0.105 | 1.039 | -5.623 | <0.001 | <0.001 |
| left pericalcarine | -0.087 | 1.082 | -4.498 | <0.001 | <0.001 |
| left postcentral | -0.125 | 1.055 | -6.594 | <0.001 | <0.001 |
| left posteriorcingulate | -0.099 | 1.046 | -5.247 | <0.001 | <0.001 |
| left precentral | -0.158 | 1.168 | -7.521 | <0.001 | <0.001 |
| left precuneus | -0.197 | 1.098 | -9.961 | <0.001 | <0.001 |
| left rostralanteriorcingulate | -0.007 | 1.014 | -0.378 | 0.706 | 0.716 |
| left rostralmiddlefrontal | -0.092 | 1.070 | -4.801 | <0.001 | <0.001 |
| left superiorfrontal | -0.138 | 1.081 | -7.083 | <0.001 | <0.001 |
| left superiorparietal | -0.154 | 1.093 | -7.851 | <0.001 | <0.001 |
| left superiortemporal | -0.252 | 1.058 | -13.228 | <0.001 | <0.001 |
| left supramarginal | -0.226 | 1.108 | -11.321 | <0.001 | <0.001 |
| left frontalpole | -0.027 | 1.009 | -1.511 | 0.131 | 0.137 |
| left temporalpole | -0.133 | 1.076 | -6.873 | <0.001 | <0.001 |
| left transversetemporal | -0.132 | 1.032 | -7.121 | <0.001 | <0.001 |
| left insula | -0.051 | 1.036 | -2.759 | 0.006 | 0.006 |
| right bankssts | -0.254 | 1.094 | -12.938 | <0.001 | <0.001 |
| right caudalanteriorcingulate | -0.006 | 1.067 | -0.332 | 0.740 | 0.740 |
| right caudalmiddlefrontal | -0.069 | 1.109 | -3.464 | 0.001 | 0.001 |
| right cuneus | -0.102 | 1.010 | -5.635 | <0.001 | <0.001 |
| right entorhinal | -0.204 | 1.047 | -10.833 | <0.001 | <0.001 |
| right fusiform | -0.277 | 1.057 | -14.569 | <0.001 | <0.001 |
| right inferiorparietal | -0.305 | 1.080 | -15.707 | <0.001 | <0.001 |
| right inferiortemporal | -0.299 | 1.056 | -15.752 | <0.001 | <0.001 |
| right isthmuscingulate | -0.244 | 1.062 | -12.798 | <0.001 | <0.001 |
| right lateraloccipital | -0.225 | 1.004 | -12.496 | <0.001 | <0.001 |
| right lateralorbitofrontal | -0.171 | 1.037 | -9.193 | <0.001 | <0.001 |
| right lingual | -0.148 | 1.006 | -8.172 | <0.001 | <0.001 |
| right medialorbitofrontal | -0.134 | 1.039 | -7.174 | <0.001 | <0.001 |
| right middletemporal | -0.319 | 1.067 | -16.652 | <0.001 | <0.001 |
| right parahippocampal | -0.194 | 1.066 | -10.142 | <0.001 | <0.001 |
| right paracentral | -0.063 | 1.115 | -3.121 | 0.002 | 0.002 |
| right parsopercularis | -0.072 | 1.042 | -3.871 | <0.001 | <0.001 |
| right parsorbitalis | -0.095 | 0.984 | -5.399 | <0.001 | <0.001 |
| right parstriangularis | -0.033 | 1.048 | -1.764 | 0.078 | 0.084 |
| right pericalcarine | -0.057 | 1.065 | -2.998 | 0.003 | 0.003 |
| right postcentral | -0.133 | 1.075 | -6.863 | <0.001 | <0.001 |
| right posteriorcingulate | -0.181 | 1.055 | -9.562 | <0.001 | <0.001 |
| right precentral | -0.096 | 1.138 | -4.708 | <0.001 | <0.001 |
| right precuneus | -0.249 | 1.098 | -12.634 | <0.001 | <0.001 |
| right rostralanteriorcingulate | -0.010 | 1.088 | -0.536 | 0.592 | 0.610 |
| right rostralmiddlefrontal | -0.029 | 1.056 | -1.514 | 0.130 | 0.137 |
| right superiorfrontal | -0.145 | 1.098 | -7.362 | <0.001 | <0.001 |
| right superiorparietal | -0.212 | 1.073 | -10.996 | <0.001 | <0.001 |
| right superiortemporal | -0.218 | 1.044 | -11.643 | <0.001 | <0.001 |
| right supramarginal | -0.255 | 1.102 | -12.861 | <0.001 | <0.001 |
| right frontalpole | -0.130 | 1.031 | -7.026 | <0.001 | <0.001 |
| right temporalpole | -0.148 | 1.089 | -7.539 | <0.001 | <0.001 |
| right transversetemporal | -0.142 | 1.062 | -7.418 | <0.001 | <0.001 |
| right insula | -0.097 | 1.046 | -5.137 | <0.001 | <0.001 |

**TABLE S4b:** One-sample t-test of cortical surface area w-score estimates.

| Region | Mean | SD | T-stat | P Value | P<sub>FDR</sub> |
| --- | --- | --- | --- | --- | --- |
| left bankssts | -0.044 | 1.048 | -2.356 | 0.019 | 0.025 |
| left caudalanteriorcingulate | -0.005 | 1.008 | -0.263 | 0.793 | 0.793 |
| left caudalmiddlefrontal | -0.017 | 1.065 | -0.873 | 0.383 | 0.407 |
| left cuneus | -0.144 | 0.989 | -8.087 | <0.001 | <0.001 |
| left entorhinal | -0.051 | 1.034 | -2.738 | 0.006 | 0.009 |
| left fusiform | -0.113 | 1.085 | -5.808 | <0.001 | <0.001 |
| left inferiorparietal | -0.122 | 1.066 | -6.387 | <0.001 | <0.001 |
| left inferiortemporal | -0.104 | 1.071 | -5.396 | <0.001 | <0.001 |
| left isthmuscingulate | 0.061 | 1.087 | 3.108 | 0.002 | 0.003 |
| left lateraloccipital | -0.190 | 1.028 | -10.289 | <0.001 | <0.001 |
| left lateralorbitofrontal | -0.125 | 1.041 | -6.698 | <0.001 | <0.001 |
| left lingual | -0.164 | 1.033 | -8.850 | <0.001 | <0.001 |
| left medialorbitofrontal | -0.083 | 1.060 | -4.337 | <0.001 | <0.001 |
| left middletemporal | -0.156 | 1.017 | -8.550 | <0.001 | <0.001 |
| left parahippocampal | -0.033 | 1.067 | -1.745 | 0.081 | 0.098 |
| left paracentral | 0.030 | 1.009 | 1.641 | 0.101 | 0.118 |
| left parsopercularis | -0.026 | 1.023 | -1.413 | 0.158 | 0.179 |
| left parsorbitalis | -0.115 | 1.059 | -6.032 | <0.001 | <0.001 |
| left parstriangularis | -0.052 | 1.018 | -2.866 | 0.004 | 0.006 |
| left pericalcarine | -0.134 | 1.017 | -7.350 | <0.001 | <0.001 |
| left postcentral | -0.017 | 1.040 | -0.921 | 0.357 | 0.391 |
| left posteriorcingulate | -0.008 | 1.080 | -0.388 | 0.698 | 0.708 |
| left precentral | -0.037 | 1.037 | -1.988 | 0.047 | 0.060 |
| left precuneus | -0.138 | 1.021 | -7.546 | <0.001 | <0.001 |
| left rostralanteriorcingulate | -0.032 | 1.065 | -1.667 | 0.096 | 0.114 |
| left rostralmiddlefrontal | -0.151 | 1.067 | -7.861 | <0.001 | <0.001 |
| left superiorfrontal | -0.107 | 1.042 | -5.708 | <0.001 | <0.001 |
| left superiorparietal | -0.154 | 0.984 | -8.737 | <0.001 | <0.001 |
| left superiortemporal | -0.082 | 1.038 | -4.372 | <0.001 | <0.001 |
| left supramarginal | -0.089 | 1.051 | -4.689 | <0.001 | <0.001 |
| left frontalpole | -0.195 | 1.007 | -10.761 | <0.001 | <0.001 |
| left temporalpole | -0.066 | 1.017 | -3.589 | <0.001 | 0.001 |
| left transversetemporal | 0.073 | 1.053 | 3.851 | <0.001 | <0.001 |
| left insula | -0.033 | 1.045 | -1.768 | 0.077 | 0.095 |
| right bankssts | -0.011 | 1.057 | -0.598 | 0.550 | 0.576 |
| right caudalanteriorcingulate | -0.028 | 1.020 | -1.507 | 0.132 | 0.152 |
| right caudalmiddlefrontal | -0.044 | 1.006 | -2.420 | 0.016 | 0.021 |
| right cuneus | -0.190 | 1.022 | -10.348 | <0.001 | <0.001 |
| right entorhinal | -0.086 | 1.005 | -4.743 | <0.001 | <0.001 |
| right fusiform | -0.116 | 1.106 | -5.858 | <0.001 | <0.001 |
| right inferiorparietal | -0.085 | 1.049 | -4.497 | <0.001 | <0.001 |
| right inferiortemporal | -0.100 | 1.037 | -5.387 | <0.001 | <0.001 |
| right isthmuscingulate | 0.104 | 1.070 | 5.399 | <0.001 | <0.001 |
| right lateraloccipital | -0.181 | 1.030 | -9.797 | <0.001 | <0.001 |
| right lateralorbitofrontal | -0.036 | 1.017 | -1.961 | 0.050 | 0.063 |
| right lingual | -0.186 | 1.001 | -10.313 | <0.001 | <0.001 |
| right medialorbitofrontal | -0.096 | 1.052 | -5.098 | <0.001 | <0.001 |
| right middletemporal | -0.154 | 1.055 | -8.142 | <0.001 | <0.001 |
| right parahippocampal | 0.019 | 1.150 | 0.911 | 0.363 | 0.391 |
| right paracentral | 0.051 | 1.069 | 2.637 | 0.008 | 0.012 |
| right parsopercularis | -0.047 | 1.056 | -2.476 | 0.013 | 0.019 |
| right parsorbitalis | -0.058 | 1.038 | -3.087 | 0.002 | 0.003 |
| right parstriangularis | -0.075 | 1.081 | -3.848 | <0.001 | <0.001 |
| right pericalcarine | -0.124 | 1.014 | -6.794 | <0.001 | <0.001 |
| right postcentral | -0.067 | 1.050 | -3.531 | <0.001 | 0.001 |
| right posteriorcingulate | -0.046 | 1.028 | -2.507 | 0.012 | 0.017 |
| right precentral | 0.019 | 1.045 | 1.035 | 0.301 | 0.335 |
| right precuneus | -0.063 | 1.082 | -3.215 | 0.001 | 0.002 |
| right rostralanteriorcingulate | -0.100 | 1.047 | -5.315 | <0.001 | <0.001 |
| right rostralmiddlefrontal | -0.152 | 1.048 | -8.074 | <0.001 | <0.001 |
| right superiorfrontal | -0.080 | 1.062 | -4.202 | <0.001 | <0.001 |
| right superiorparietal | -0.098 | 1.016 | -5.345 | <0.001 | <0.001 |
| right superiortemporal | -0.048 | 1.054 | -2.523 | 0.012 | 0.017 |
| right supramarginal | -0.075 | 1.036 | -4.026 | <0.001 | <0.001 |
| right frontalpole | -0.080 | 1.013 | -4.375 | <0.001 | <0.001 |
| right temporalpole | -0.042 | 1.034 | -2.277 | 0.023 | 0.030 |
| right transversetemporal | 0.007 | 1.028 | 0.391 | 0.696 | 0.708 |
| right insula | -0.078 | 1.040 | -4.146 | <0.001 | <0.001 |

**TABLE S4c:** One-sample t-test of subcortical volume w-score estimates.

| Region | Mean | SD | T-stat | P Value | P<sub>FDR</sub> |
| --- | --- | --- | --- | --- | --- |
| left lateral ventricle | 0.132 | 1.167 | 6.292 | <0.001 | <0.001 |
| left thalamus | 0.087 | 1.085 | 4.438 | <0.001 | <0.001 |
| left caudate | -0.120 | 1.113 | -5.983 | <0.001 | <0.001 |
| left putamen | -0.204 | 1.100 | -10.298 | <0.001 | <0.001 |
| left pallidum | -0.068 | 1.122 | -3.353 | 0.001 | 0.001 |
| left hippocampus | -0.143 | 1.078 | -7.379 | <0.001 | <0.001 |
| left amygdala | -0.252 | 1.057 | -13.263 | <0.001 | <0.001 |
| left accumbens | -0.133 | 1.041 | -7.131 | <0.001 | <0.001 |
| right lateral ventricle | 0.173 | 1.201 | 8.033 | <0.001 | <0.001 |
| right thalamus | 0.038 | 1.092 | 1.957 | 0.050 | 0.054 |
| right caudate | -0.051 | 1.090 | -2.599 | 0.009 | 0.011 |
| right putamen | -0.197 | 1.072 | -10.229 | <0.001 | <0.001 |
| right pallidum | -0.030 | 1.097 | -1.542 | 0.123 | 0.123 |
| right hippocampus | -0.153 | 1.086 | -7.826 | <0.001 | <0.001 |
| right amygdala | -0.176 | 1.073 | -9.131 | <0.001 | <0.001 |
| right accumbens | -0.133 | 1.078 | -6.844 | <0.001 | <0.001 |

**TABLE S5a:** Relationships between cortical thickness with clinical variables in PD.

| Region | HY disease stage |  |  | Time since diagnosis |  |  | Global cognition |  |  |
| --- | --- | --- | --- | --- | --- | --- | --- | --- | --- |
| | $\rho$ | P value | P<sub>FDR</sub> | $\beta$ | P value | P<sub>FDR</sub> | $\beta$ | P value | P<sub>FDR</sub> |
| left bankssts | -0.059 | 0.003 | 0.008 | -0.007 | 0.060 | 0.091 | -0.027 | 0.001 | 0.003 |
| left caudalanteriorcingulate | 0.029 | 0.146 | 0.168 | 0.002 | 0.596 | 0.623 | 0.005 | 0.532 | 0.557 |
| left caudalmiddlefrontal | -0.040 | 0.045 | 0.063 | -0.013 | 0.001 | 0.006 | -0.027 | 0.001 | 0.003 |
| left cuneus | -0.031 | 0.118 | 0.144 | -0.009 | 0.012 | 0.026 | -0.004 | 0.608 | 0.626 |
| left entorhinal | -0.075 | 0.000 | 0.001 | -0.009 | 0.019 | 0.037 | -0.013 | 0.101 | 0.120 |
| left fusiform | -0.085 | 0.000 | 0.000 | -0.016 | 0.000 | 0.000 | -0.042 | 0.000 | 0.000 |
| left inferiorparietal | -0.079 | 0.000 | 0.000 | -0.019 | 0.000 | 0.000 | -0.023 | 0.005 | 0.010 |
| left inferior temporal | -0.080 | 0.000 | 0.000 | -0.012 | 0.002 | 0.006 | -0.032 | 0.000 | 0.000 |
| left isthmuscingulate | -0.066 | 0.001 | 0.003 | -0.008 | 0.032 | 0.055 | -0.018 | 0.018 | 0.028 |
| left lateraloccipital | -0.049 | 0.014 | 0.025 | -0.008 | 0.029 | 0.050 | -0.006 | 0.425 | 0.452 |
| left lateralorbitofrontal | -0.052 | 0.009 | 0.019 | -0.007 | 0.099 | 0.137 | -0.015 | 0.077 | 0.099 |
| left lingual | -0.082 | 0.000 | 0.000 | -0.006 | 0.126 | 0.168 | -0.021 | 0.006 | 0.011 |
| left medialorbitofrontal | -0.021 | 0.290 | 0.299 | -0.004 | 0.275 | 0.323 | -0.008 | 0.338 | 0.365 |
| left middletemporal | -0.113 | 0.000 | 0.000 | -0.012 | 0.001 | 0.006 | -0.037 | 0.000 | 0.000 |
| left parahippocampal | -0.056 | 0.004 | 0.011 | -0.007 | 0.062 | 0.091 | -0.013 | 0.085 | 0.105 |
| left paracentral | -0.024 | 0.231 | 0.254 | -0.004 | 0.318 | 0.355 | -0.028 | 0.001 | 0.004 |
| left parsopercularis | -0.071 | 0.000 | 0.002 | -0.013 | 0.001 | 0.004 | -0.025 | 0.002 | 0.005 |
| left parsorbitalis | -0.034 | 0.088 | 0.109 | -0.005 | 0.195 | 0.239 | -0.012 | 0.112 | 0.132 |
| left parstriangularis | -0.046 | 0.020 | 0.031 | -0.014 | 0.000 | 0.002 | -0.033 | 0.000 | 0.000 |
| left pericalcarine | -0.035 | 0.080 | 0.100 | -0.008 | 0.043 | 0.071 | -0.017 | 0.043 | 0.058 |
| left postcentral | -0.038 | 0.053 | 0.069 | -0.010 | 0.009 | 0.020 | -0.039 | 0.000 | 0.000 |
| left posteriorcingulate | -0.031 | 0.120 | 0.144 | -0.003 | 0.370 | 0.405 | -0.011 | 0.165 | 0.184 |
| left precentral | -0.051 | 0.011 | 0.022 | -0.012 | 0.005 | 0.013 | -0.042 | 0.000 | 0.000 |
| left precuneus | -0.066 | 0.001 | 0.003 | -0.017 | 0.000 | 0.000 | -0.022 | 0.009 | 0.015 |
| left rostralanteriorcingulate | -0.021 | 0.297 | 0.301 | -0.004 | 0.314 | 0.355 | -0.011 | 0.149 | 0.169 |
| left rostralmiddlefrontal | -0.047 | 0.018 | 0.029 | -0.011 | 0.007 | 0.016 | -0.036 | 0.000 | 0.000 |
| left superiorfrontal | -0.039 | 0.049 | 0.067 | -0.011 | 0.006 | 0.015 | -0.026 | 0.002 | 0.004 |
| left superiorparietal | -0.053 | 0.007 | 0.018 | -0.015 | 0.000 | 0.001 | -0.027 | 0.001 | 0.004 |
| left superior temporal | -0.097 | 0.000 | 0.000 | -0.012 | 0.002 | 0.006 | -0.037 | 0.000 | 0.000 |
| left supramarginal | -0.089 | 0.000 | 0.000 | -0.010 | 0.014 | 0.029 | -0.030 | 0.000 | 0.002 |
| left frontalpole | -0.039 | 0.050 | 0.067 | -0.004 | 0.275 | 0.323 | -0.023 | 0.003 | 0.007 |
| left temporalpole | -0.050 | 0.012 | 0.023 | -0.013 | 0.001 | 0.004 | -0.018 | 0.029 | 0.041 |
| left transversetemporal | -0.066 | 0.001 | 0.003 | -0.014 | 0.000 | 0.002 | -0.029 | 0.000 | 0.001 |
| left insula | -0.049 | 0.012 | 0.023 | -0.007 | 0.048 | 0.076 | -0.023 | 0.004 | 0.008 |
| right bankssts | -0.056 | 0.004 | 0.011 | -0.005 | 0.197 | 0.239 | -0.025 | 0.003 | 0.006 |
| right caudalanteriorcingulate | -0.047 | 0.017 | 0.028 | -0.003 | 0.506 | 0.538 | -0.010 | 0.210 | 0.231 |
| right caudalmiddlefrontal | -0.037 | 0.063 | 0.081 | -0.008 | 0.046 | 0.074 | -0.027 | 0.002 | 0.004 |
| right cuneus | -0.022 | 0.268 | 0.285 | -0.007 | 0.054 | 0.083 | -0.004 | 0.623 | 0.632 |
| right entorhinal | -0.067 | 0.001 | 0.003 | -0.012 | 0.002 | 0.006 | -0.015 | 0.069 | 0.091 |
| right fusiform | -0.084 | 0.000 | 0.000 | -0.013 | 0.001 | 0.005 | -0.041 | 0.000 | 0.000 |
| right inferiorparietal | -0.067 | 0.001 | 0.003 | -0.018 | 0.000 | 0.000 | -0.024 | 0.003 | 0.007 |
| right inferior temporal | -0.059 | 0.003 | 0.008 | -0.012 | 0.002 | 0.006 | -0.028 | 0.001 | 0.002 |
| right isthmuscingulate | -0.051 | 0.010 | 0.022 | -0.003 | 0.376 | 0.406 | -0.019 | 0.019 | 0.029 |
| right lateraloccipital | -0.042 | 0.034 | 0.050 | -0.013 | 0.001 | 0.003 | -0.021 | 0.007 | 0.013 |
| right lateralorbitofrontal | -0.053 | 0.008 | 0.018 | -0.006 | 0.113 | 0.154 | -0.019 | 0.015 | 0.024 |
| right lingual | -0.049 | 0.014 | 0.025 | -0.006 | 0.096 | 0.135 | -0.019 | 0.017 | 0.027 |
| right medialorbitofrontal | -0.041 | 0.038 | 0.054 | -0.005 | 0.191 | 0.239 | -0.012 | 0.123 | 0.142 |
| right middletemporal | -0.069 | 0.000 | 0.002 | -0.010 | 0.008 | 0.019 | -0.030 | 0.000 | 0.001 |
| right parahippocampal | -0.051 | 0.010 | 0.022 | -0.002 | 0.699 | 0.721 | -0.025 | 0.002 | 0.004 |
| right paracentral | -0.023 | 0.256 | 0.276 | -0.006 | 0.145 | 0.185 | -0.028 | 0.001 | 0.003 |
| right parsopercularis | -0.044 | 0.025 | 0.036 | -0.012 | 0.002 | 0.006 | -0.018 | 0.026 | 0.038 |
| right parsorbitalis | -0.030 | 0.125 | 0.146 | -0.005 | 0.143 | 0.185 | -0.013 | 0.094 | 0.114 |
| right parstriangularis | -0.022 | 0.275 | 0.288 | -0.007 | 0.081 | 0.117 | -0.019 | 0.018 | 0.028 |
| right pericalcarine | -0.025 | 0.198 | 0.224 | -0.004 | 0.308 | 0.355 | -0.017 | 0.042 | 0.058 |
| right postcentral | -0.024 | 0.218 | 0.243 | -0.010 | 0.013 | 0.026 | -0.033 | 0.000 | 0.000 |
| right posteriorcingulate | -0.081 | 0.000 | 0.000 | -0.015 | 0.000 | 0.001 | -0.014 | 0.078 | 0.099 |
| right precentral | -0.045 | 0.024 | 0.036 | -0.010 | 0.020 | 0.037 | -0.037 | 0.000 | 0.000 |
| right precuneus | -0.068 | 0.001 | 0.002 | -0.018 | 0.000 | 0.000 | -0.030 | 0.000 | 0.002 |
| right rostralanteriorcingulate | 0.007 | 0.707 | 0.707 | 0.000 | 0.928 | 0.928 | -0.003 | 0.726 | 0.726 |
| right rostralmiddlefrontal | -0.047 | 0.017 | 0.028 | -0.009 | 0.023 | 0.042 | -0.015 | 0.054 | 0.073 |
| right superiorfrontal | -0.050 | 0.011 | 0.022 | -0.013 | 0.001 | 0.004 | -0.030 | 0.000 | 0.002 |
| right superiorparietal | -0.050 | 0.012 | 0.023 | -0.018 | 0.000 | 0.000 | -0.024 | 0.004 | 0.007 |
| right superior temporal | -0.070 | 0.000 | 0.002 | -0.008 | 0.026 | 0.046 | -0.032 | 0.000 | 0.000 |
| right supramarginal | -0.091 | 0.000 | 0.000 | -0.013 | 0.001 | 0.004 | -0.022 | 0.007 | 0.013 |
| right frontalpole | -0.047 | 0.017 | 0.028 | -0.001 | 0.780 | 0.791 | -0.019 | 0.015 | 0.024 |
| right temporalpole | -0.089 | 0.000 | 0.000 | -0.012 | 0.002 | 0.006 | -0.023 | 0.004 | 0.009 |
| right transversetemporal | -0.063 | 0.001 | 0.004 | -0.012 | 0.002 | 0.006 | -0.029 | 0.000 | 0.002 |
| right insula | -0.061 | 0.002 | 0.006 | -0.010 | 0.011 | 0.025 | -0.026 | 0.001 | 0.003 |

**TABLE S5b:**
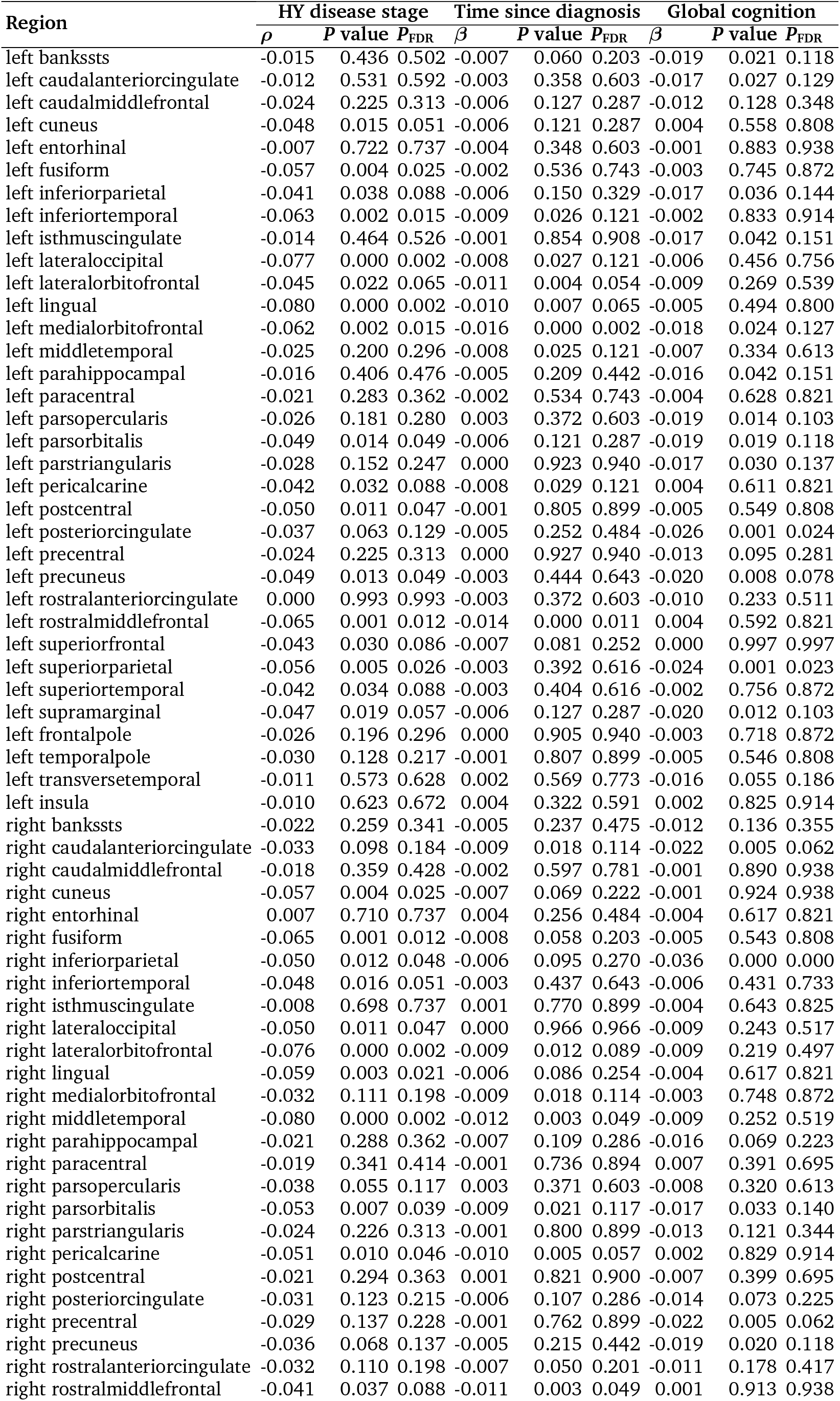
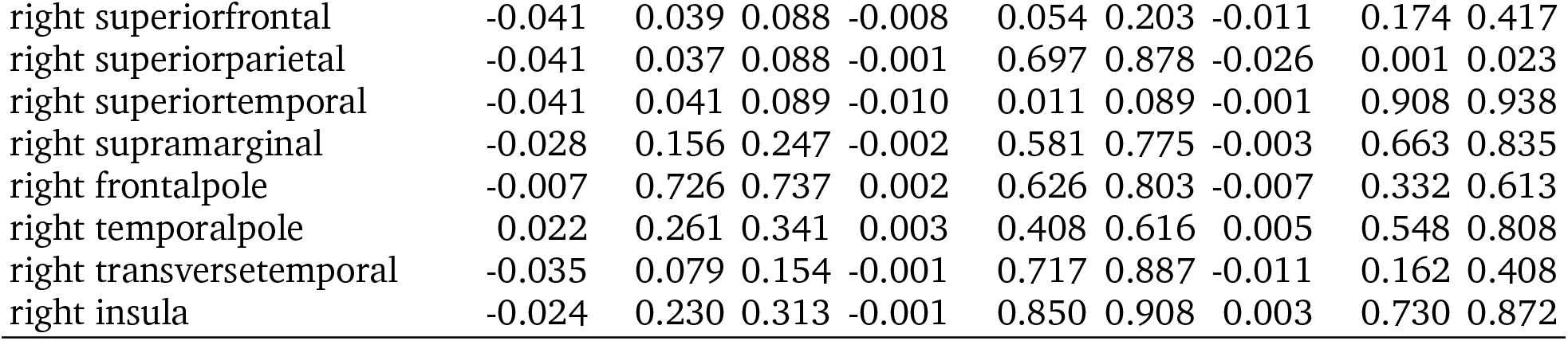
Relationships between cortical surface area with clinical variables in PD.

| Region | HY disease stage |  |  | Time since diagnosis |  |  | Global cognition |  |  |
| --- | --- | --- | --- | --- | --- | --- | --- | --- | --- |
| | $\rho$ | P value | P<sub>FDR</sub> | $\beta$ | P value | P<sub>FDR</sub> | $\beta$ | P value | P<sub>FDR</sub> |
| left bankssts | -0.015 | 0.436 | 0.502 | -0.007 | 0.060 | 0.203 | -0.019 | 0.021 | 0.118 |
| left caudalanteriorcingulate | -0.012 | 0.531 | 0.592 | -0.003 | 0.358 | 0.603 | -0.017 | 0.027 | 0.129 |
| left caudalmiddlefrontal | -0.024 | 0.225 | 0.313 | -0.006 | 0.127 | 0.287 | -0.012 | 0.128 | 0.348 |
| left cuneus | -0.048 | 0.015 | 0.051 | -0.006 | 0.121 | 0.287 | 0.004 | 0.558 | 0.808 |
| left entorhinal | -0.007 | 0.722 | 0.737 | -0.004 | 0.348 | 0.603 | -0.001 | 0.883 | 0.938 |
| left fusiform | -0.057 | 0.004 | 0.025 | -0.002 | 0.536 | 0.743 | -0.003 | 0.745 | 0.872 |
| left inferiorparietal | -0.041 | 0.038 | 0.088 | -0.006 | 0.150 | 0.329 | -0.017 | 0.036 | 0.144 |
| left inferiortemporal | -0.063 | 0.002 | 0.015 | -0.009 | 0.026 | 0.121 | -0.002 | 0.833 | 0.914 |
| left isthmuscingulate | -0.014 | 0.464 | 0.526 | -0.001 | 0.854 | 0.908 | -0.017 | 0.042 | 0.151 |
| left lateraloccipital | -0.077 | 0.000 | 0.002 | -0.008 | 0.027 | 0.121 | -0.006 | 0.456 | 0.756 |
| left lateralorbitofrontal | -0.045 | 0.022 | 0.065 | -0.011 | 0.004 | 0.054 | -0.009 | 0.269 | 0.539 |
| left lingual | -0.080 | 0.000 | 0.002 | -0.010 | 0.007 | 0.065 | -0.005 | 0.494 | 0.800 |
| left medialorbitofrontal | -0.062 | 0.002 | 0.015 | -0.016 | 0.000 | 0.002 | -0.018 | 0.024 | 0.127 |
| left middletemporal | -0.025 | 0.200 | 0.296 | -0.008 | 0.025 | 0.121 | -0.007 | 0.334 | 0.613 |
| left parahippocampal | -0.016 | 0.406 | 0.476 | -0.005 | 0.209 | 0.442 | -0.016 | 0.042 | 0.151 |
| left paracentral | -0.021 | 0.283 | 0.362 | -0.002 | 0.534 | 0.743 | -0.004 | 0.628 | 0.821 |
| left parsopercularis | -0.026 | 0.181 | 0.280 | 0.003 | 0.372 | 0.603 | -0.019 | 0.014 | 0.103 |
| left parsorbitalis | -0.049 | 0.014 | 0.049 | -0.006 | 0.121 | 0.287 | -0.019 | 0.019 | 0.118 |
| left parstriangularis | -0.028 | 0.152 | 0.247 | 0.000 | 0.923 | 0.940 | -0.017 | 0.030 | 0.137 |
| left pericalcarine | -0.042 | 0.032 | 0.088 | -0.008 | 0.029 | 0.121 | 0.004 | 0.611 | 0.821 |
| left postcentral | -0.050 | 0.011 | 0.047 | -0.001 | 0.805 | 0.899 | -0.005 | 0.549 | 0.808 |
| left posteriorcingulate | -0.037 | 0.063 | 0.129 | -0.005 | 0.252 | 0.484 | -0.026 | 0.001 | 0.024 |
| left precentral | -0.024 | 0.225 | 0.313 | 0.000 | 0.927 | 0.940 | -0.013 | 0.095 | 0.281 |
| left precuneus | -0.049 | 0.013 | 0.049 | -0.003 | 0.444 | 0.643 | -0.020 | 0.008 | 0.078 |
| left rostralanteriorcingulate | 0.000 | 0.993 | 0.993 | -0.003 | 0.372 | 0.603 | -0.010 | 0.233 | 0.511 |
| left rostralmiddlefrontal | -0.065 | 0.001 | 0.012 | -0.014 | 0.000 | 0.011 | 0.004 | 0.592 | 0.821 |
| left superiorfrontal | -0.043 | 0.030 | 0.086 | -0.007 | 0.081 | 0.252 | 0.000 | 0.997 | 0.997 |
| left superiorparietal | -0.056 | 0.005 | 0.026 | -0.003 | 0.392 | 0.616 | -0.024 | 0.001 | 0.023 |
| left superiortemporal | -0.042 | 0.034 | 0.088 | -0.003 | 0.404 | 0.616 | -0.002 | 0.756 | 0.872 |
| left supramarginal | -0.047 | 0.019 | 0.057 | -0.006 | 0.127 | 0.287 | -0.020 | 0.012 | 0.103 |
| left frontalpole | -0.026 | 0.196 | 0.296 | 0.000 | 0.905 | 0.940 | -0.003 | 0.718 | 0.872 |
| left temporalpole | -0.030 | 0.128 | 0.217 | -0.001 | 0.807 | 0.899 | -0.005 | 0.546 | 0.808 |
| left transversetemporal | -0.011 | 0.573 | 0.628 | 0.002 | 0.569 | 0.773 | -0.016 | 0.055 | 0.186 |
| left insula | -0.010 | 0.623 | 0.672 | 0.004 | 0.322 | 0.591 | 0.002 | 0.825 | 0.914 |
| right bankssts | -0.022 | 0.259 | 0.341 | -0.005 | 0.237 | 0.475 | -0.012 | 0.136 | 0.355 |
| right caudalanteriorcingulate | -0.033 | 0.098 | 0.184 | -0.009 | 0.018 | 0.114 | -0.022 | 0.005 | 0.062 |
| right caudalmiddlefrontal | -0.018 | 0.359 | 0.428 | -0.002 | 0.597 | 0.781 | -0.001 | 0.890 | 0.938 |
| right cuneus | -0.057 | 0.004 | 0.025 | -0.007 | 0.069 | 0.222 | -0.001 | 0.924 | 0.938 |
| right entorhinal | 0.007 | 0.710 | 0.737 | 0.004 | 0.256 | 0.484 | -0.004 | 0.617 | 0.821 |
| right fusiform | -0.065 | 0.001 | 0.012 | -0.008 | 0.058 | 0.203 | -0.005 | 0.543 | 0.808 |
| right inferiorparietal | -0.050 | 0.012 | 0.048 | -0.006 | 0.095 | 0.270 | -0.036 | 0.000 | 0.000 |
| right inferiortemporal | -0.048 | 0.016 | 0.051 | -0.003 | 0.437 | 0.643 | -0.006 | 0.431 | 0.733 |
| right isthmuscingulate | -0.008 | 0.698 | 0.737 | 0.001 | 0.770 | 0.899 | -0.004 | 0.643 | 0.825 |
| right lateraloccipital | -0.050 | 0.011 | 0.047 | 0.000 | 0.966 | 0.966 | -0.009 | 0.243 | 0.517 |
| right lateralorbitofrontal | -0.076 | 0.000 | 0.002 | -0.009 | 0.012 | 0.089 | -0.009 | 0.219 | 0.497 |
| right lingual | -0.059 | 0.003 | 0.021 | -0.006 | 0.086 | 0.254 | -0.004 | 0.617 | 0.821 |
| right medialorbitofrontal | -0.032 | 0.111 | 0.198 | -0.009 | 0.018 | 0.114 | -0.003 | 0.748 | 0.872 |
| right middletemporal | -0.080 | 0.000 | 0.002 | -0.012 | 0.003 | 0.049 | -0.009 | 0.252 | 0.519 |
| right parahippocampal | -0.021 | 0.288 | 0.362 | -0.007 | 0.109 | 0.286 | -0.016 | 0.069 | 0.223 |
| right paracentral | -0.019 | 0.341 | 0.414 | -0.001 | 0.736 | 0.894 | 0.007 | 0.391 | 0.695 |
| right parsopercularis | -0.038 | 0.055 | 0.117 | 0.003 | 0.371 | 0.603 | -0.008 | 0.320 | 0.613 |
| right parsorbitalis | -0.053 | 0.007 | 0.039 | -0.009 | 0.021 | 0.117 | -0.017 | 0.033 | 0.140 |
| right parstriangularis | -0.024 | 0.226 | 0.313 | -0.001 | 0.800 | 0.899 | -0.013 | 0.121 | 0.344 |
| right pericalcarine | -0.051 | 0.010 | 0.046 | -0.010 | 0.005 | 0.057 | 0.002 | 0.829 | 0.914 |
| right postcentral | -0.021 | 0.294 | 0.363 | 0.001 | 0.821 | 0.900 | -0.007 | 0.399 | 0.695 |
| right posteriorcingulate | -0.031 | 0.123 | 0.215 | -0.006 | 0.107 | 0.286 | -0.014 | 0.073 | 0.225 |
| right precentral | -0.029 | 0.137 | 0.228 | -0.001 | 0.762 | 0.899 | -0.022 | 0.005 | 0.062 |
| right precuneus | -0.036 | 0.068 | 0.137 | -0.005 | 0.215 | 0.442 | -0.019 | 0.020 | 0.118 |
| right rostralanteriorcingulate | -0.032 | 0.110 | 0.198 | -0.007 | 0.050 | 0.201 | -0.011 | 0.178 | 0.417 |
| right rostralmiddlefrontal | -0.041 | 0.037 | 0.088 | -0.011 | 0.003 | 0.049 | 0.001 | 0.913 | 0.938 |
| right superiorfrontal | -0.041 | 0.039 | 0.088 | -0.008 | 0.054 | 0.203 | -0.011 | 0.174 | 0.417 |
| right superiorparietal | -0.041 | 0.037 | 0.088 | -0.001 | 0.697 | 0.878 | -0.026 | 0.001 | 0.023 |
| right superiortemporal | -0.041 | 0.041 | 0.089 | -0.010 | 0.011 | 0.089 | -0.001 | 0.908 | 0.938 |
| right supramarginal | -0.028 | 0.156 | 0.247 | -0.002 | 0.581 | 0.775 | -0.003 | 0.663 | 0.835 |
| right frontalpole | -0.007 | 0.726 | 0.737 | 0.002 | 0.626 | 0.803 | -0.007 | 0.332 | 0.613 |
| right temporalpole | 0.022 | 0.261 | 0.341 | 0.003 | 0.408 | 0.616 | 0.005 | 0.548 | 0.808 |
| right transversetemporal | -0.035 | 0.079 | 0.154 | -0.001 | 0.717 | 0.887 | -0.011 | 0.162 | 0.408 |
| right insula | -0.024 | 0.230 | 0.313 | -0.001 | 0.850 | 0.908 | 0.003 | 0.730 | 0.872 |

**TABLE S5c:** Relationships between subcortical volume with clinical variables in PD.

| Region | HY disease stage |  |  | Time since diagnosis |  |  | Global cognition |  |  |
| --- | --- | --- | --- | --- | --- | --- | --- | --- | --- |
| | $\rho$ | *P* value | *P*<sub>FDR</sub> | $\beta$ | *P* value | *P*<sub>FDR</sub> | $\beta$ | *P* value | *P*<sub>FDR</sub> |
| left lateral ventricle | 0.088 | 0.000 | 0.000 | 0.004 | 0.361 | 0.413 | 0.039 | 0.000 | 0.000 |
| left thalamus | -0.063 | 0.002 | 0.003 | -0.012 | 0.002 | 0.005 | -0.007 | 0.352 | 0.375 |
| left caudate | -0.061 | 0.002 | 0.003 | -0.019 | 0.000 | 0.000 | -0.013 | 0.108 | 0.133 |
| left putamen | -0.071 | 0.000 | 0.001 | -0.011 | 0.005 | 0.009 | -0.036 | 0.000 | 0.000 |
| left pallidum | -0.028 | 0.152 | 0.152 | 0.000 | 0.982 | 0.982 | -0.018 | 0.039 | 0.057 |
| left hippocampus | -0.102 | 0.000 | 0.000 | -0.015 | 0.000 | 0.000 | -0.029 | 0.000 | 0.001 |
| left amygdala | -0.092 | 0.000 | 0.000 | -0.023 | 0.000 | 0.000 | -0.041 | 0.000 | 0.000 |
| left accumbens | -0.061 | 0.002 | 0.003 | -0.010 | 0.010 | 0.014 | -0.028 | 0.001 | 0.001 |
| right lateral ventricle | 0.080 | 0.000 | 0.000 | 0.005 | 0.257 | 0.317 | 0.038 | 0.000 | 0.000 |
| right thalamus | -0.047 | 0.018 | 0.021 | -0.012 | 0.003 | 0.005 | -0.007 | 0.412 | 0.412 |
| right caudate | -0.059 | 0.003 | 0.003 | -0.016 | 0.000 | 0.000 | -0.015 | 0.067 | 0.089 |
| right putamen | -0.061 | 0.002 | 0.003 | -0.008 | 0.052 | 0.070 | -0.023 | 0.005 | 0.007 |
| right pallidum | -0.032 | 0.111 | 0.119 | -0.002 | 0.580 | 0.619 | -0.010 | 0.220 | 0.251 |
| right hippocampus | -0.095 | 0.000 | 0.000 | -0.016 | 0.000 | 0.000 | -0.042 | 0.000 | 0.000 |
| right amygdala | -0.082 | 0.000 | 0.000 | -0.021 | 0.000 | 0.000 | -0.036 | 0.000 | 0.000 |
| right accumbens | -0.083 | 0.000 | 0.000 | -0.011 | 0.005 | 0.009 | -0.026 | 0.001 | 0.003 |

**TABLE S6a:** Group-average node-neighbourhood correlations.

| Weighting | Network model | Correlation ($\rho$) | *P* value | *P*<sub>spin/perm</sub> |
| --- | --- | --- | --- | --- |
| structural connectivity | cortico-cortical | 0.545 | 0.000 | 0.001 |
|  | subcortico-cortical | 0.187 | 0.523 | 0.495 |
| functional connectivity | cortico-cortical | 0.366 | 0.002 | 0.071 |
|  | subcortico-cortical | 0.020 | 0.946 | 0.921 |

**TABLE S6b:** Disease stage node-neighbourhood correlations.

| Weighting | Network model | HY stage | Correlation ($\rho$) | $P$ value | $P_{\text{spin/perm}}$ |
| --- | --- | --- | --- | --- | --- |
| structural connectivity | cortico-cortical | 1 | 0.479 | <0.001 | 0.004 |
|  |  | 2 | 0.518 | <0.001 | 0.003 |
|  |  | 3 | 0.564 | <0.001 | 0.001 |
|  |  | 4/5 | 0.381 | 0.001 | 0.025 |
|  | subcortico-cortical | 1 | -0.204 | 0.483 | 0.478 |
|  |  | 2 | 0.015 | 0.958 | 0.918 |
|  |  | 3 | 0.209 | 0.474 | 0.449 |
|  |  | 4/5 | 0.349 | 0.221 | 0.238 |
| functional connectivity | cortico-cortical | 1 | 0.516 | <0.001 | 0.006 |
|  |  | 2 | 0.327 | 0.007 | 0.104 |
|  |  | 3 | 0.387 | 0.001 | 0.056 |
|  |  | 4/5 | 0.228 | 0.061 | 0.258 |
|  | subcortico-cortical | 1 | -0.385 | 0.175 | 0.165 |
|  |  | 2 | -0.077 | 0.794 | 0.806 |
|  |  | 3 | 0.130 | 0.659 | 0.679 |
|  |  | 4/5 | 0.112 | 0.703 | 0.671 |

**TABLE S7a:**
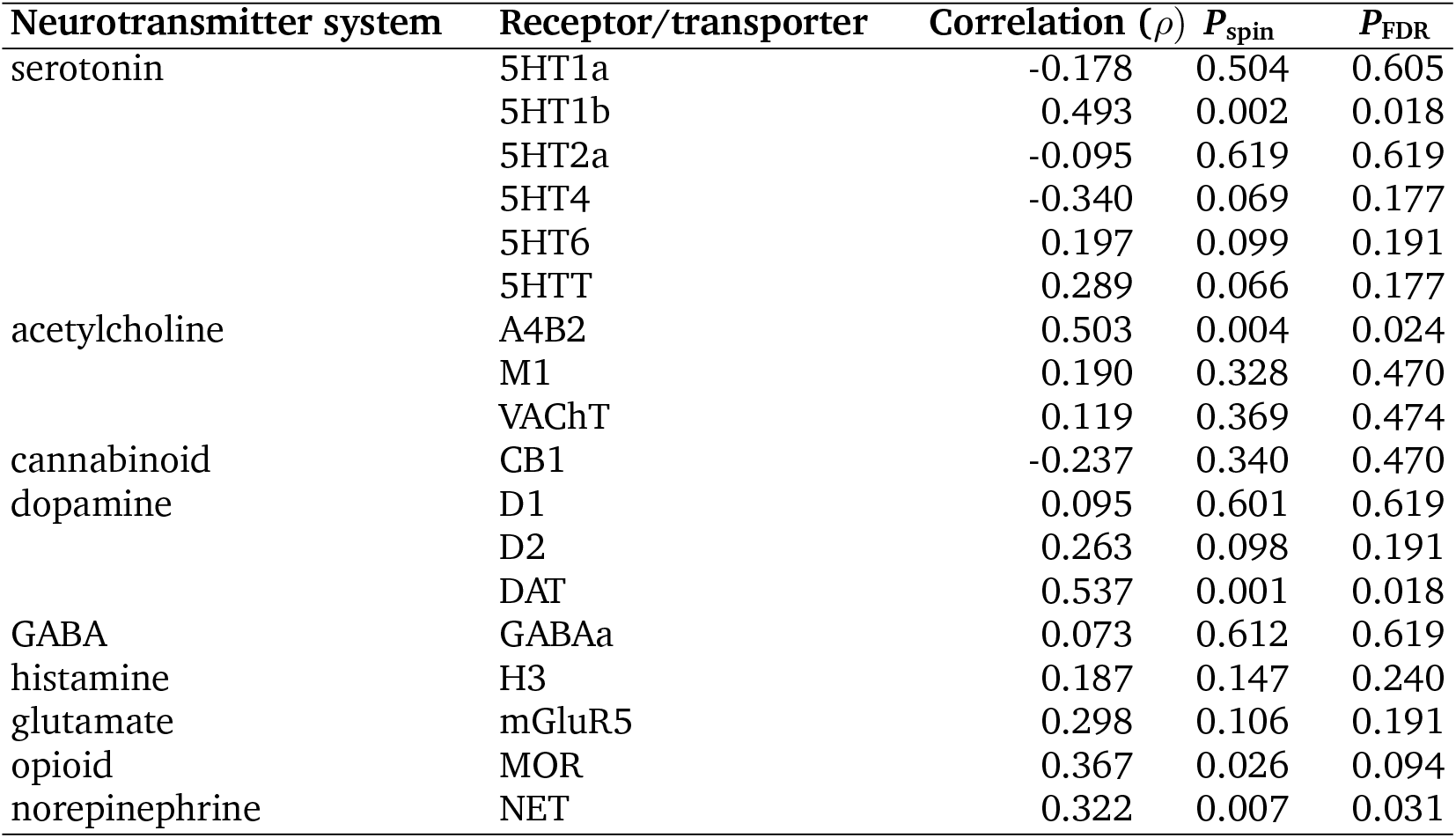
Neurotransmitter system annotation enrichment of cortical thickness.

| Neurotransmitter system | Receptor/transporter | Correlation ($\rho$) | $P_{\text{spin}}$ | $P_{\text{FDR}}$ |
| --- | --- | --- | --- | --- |
| serotonin | 5HT1a | -0.178 | 0.504 | 0.605 |
|  | 5HT1b | 0.493 | 0.002 | 0.018 |
|  | 5HT2a | -0.095 | 0.619 | 0.619 |
|  | 5HT4 | -0.340 | 0.069 | 0.177 |
|  | 5HT6 | 0.197 | 0.099 | 0.191 |
|  | 5HTT | 0.289 | 0.066 | 0.177 |
| acetylcholine | A4B2 | 0.503 | 0.004 | 0.024 |
|  | M1 | 0.190 | 0.328 | 0.470 |
|  | VACHT | 0.119 | 0.369 | 0.474 |
| cannabinoid | CB1 | -0.237 | 0.340 | 0.470 |
| dopamine | D1 | 0.095 | 0.601 | 0.619 |
|  | D2 | 0.263 | 0.098 | 0.191 |
|  | DAT | 0.537 | 0.001 | 0.018 |
| GABA | GABAA | 0.073 | 0.612 | 0.619 |
| histamine | H3 | 0.187 | 0.147 | 0.240 |
| glutamate | mGluR5 | 0.298 | 0.106 | 0.191 |
| opioid | MOR | 0.367 | 0.026 | 0.094 |
| norepinephrine | NET | 0.322 | 0.007 | 0.031 |

**TABLE S7b:** Neurotransmitter system annotation enrichment of cortical surface area.

| Neurotransmitter system | Receptor/transporter | Correlation ($\rho$) | $P_{\text{spin}}$ | $P_{\text{FDR}}$ |
| --- | --- | --- | --- | --- |
| serotonin | 5HT1a | 0.195 | 0.316 | 0.517 |
|  | 5HT1b | -0.074 | 0.750 | 0.831 |
|  | 5HT2a | -0.287 | 0.082 | 0.184 |
|  | 5HT4 | -0.082 | 0.501 | 0.694 |
|  | 5HT6 | 0.006 | 0.831 | 0.831 |
|  | 5HTT | 0.304 | 0.078 | 0.184 |
| acetylcholine | A4B2 | 0.358 | 0.012 | 0.054 |
|  | M1 | 0.044 | 0.750 | 0.831 |
|  | VACHT | 0.174 | 0.437 | 0.655 |
| cannabinoid | CB1 | 0.099 | 0.823 | 0.831 |
| dopamine | D1 | 0.397 | 0.028 | 0.101 |
|  | D2 | -0.005 | 0.801 | 0.831 |
|  | DAT | 0.343 | 0.012 | 0.054 |
| GABA | GABAA | -0.325 | 0.010 | 0.054 |
| histamine | H3 | 0.258 | 0.126 | 0.252 |
| glutamate | mGluR5 | 0.181 | 0.205 | 0.369 |
| opioid | MOR | 0.329 | 0.036 | 0.108 |
| norepinephrine | NET | 0.595 | 0.001 | 0.018 |

**TABLE S7c:** Neurotransmitter system annotation enrichment of subcortical volume.

| Neurotransmitter system | Receptor/transporter | Correlation ($\rho$) | $P_{\text{perm}}$ | $P_{\text{FDR}}$ |
| --- | --- | --- | --- | --- |
| serotonin | 5HT1a | -0.662 | 0.009 | 0.081 |
|  | 5HT1b | -0.055 | 0.847 | 0.953 |
|  | 5HT2a | -0.569 | 0.040 | 0.126 |
|  | 5HT4 | -0.244 | 0.442 | 0.759 |
|  | 5HT6 | -0.262 | 0.361 | 0.721 |
|  | 5HTT | 0.130 | 0.656 | 0.844 |
| acetylcholine | A4B2 | 0.552 | 0.042 | 0.126 |
|  | M1 | -0.464 | 0.099 | 0.241 |
|  | VACHT | -0.169 | 0.552 | 0.770 |
| cannabinoid | CB1 | -0.002 | 0.991 | 0.991 |
| dopamine | D1 | 0.073 | 0.798 | 0.953 |
|  | D2 | -0.688 | 0.009 | 0.081 |
|  | DAT | -0.165 | 0.556 | 0.770 |
| GABA | GABAA | -0.627 | 0.016 | 0.096 |
| histamine | H3 | -0.596 | 0.023 | 0.103 |
| glutamate | mGluR5 | 0.209 | 0.464 | 0.759 |
| opioid | MOR | 0.451 | 0.107 | 0.241 |
| norepinephrine | NET | 0.011 | 0.946 | 0.991 |

**TABLE S8a:** Summary of gene set enrichment analysis of biological processes for PLS1.

| Gene Set | Description | Size | Leading Edge Number | ES | NES | P value | P<sub>FDR</sub> |
| --- | --- | --- | --- | --- | --- | --- | --- |
| GO:0099003 | vesicle-mediated transport in synapse | 205 | 96 | -0.456 | -2.305 | <0.001 | <0.001 |
| GO:0035249 | synaptic transmission, glutamatergic | 97 | 36 | -0.431 | -1.921 | <0.001 | 0.045 |
| GO:0048499 | synaptic vesicle membrane organization | 27 | 10 | -0.548 | -1.921 | <0.001 | 0.031 |
| GO:0051648 | vesicle localization | 203 | 73 | -0.374 | -1.874 | <0.001 | 0.044 |
| GO:0006813 | potassium ion transport | 191 | 69 | -0.352 | -1.744 | <0.001 | 0.142 |
| GO:0034728 | nucleosome organization | 61 | 22 | -0.421 | -1.730 | 0.004 | 0.137 |
| GO:0016358 | dendrite development | 217 | 64 | -0.337 | -1.703 | <0.001 | 0.147 |
| GO:0099177 | regulation of trans-synaptic signaling | 425 | 125 | -0.305 | -1.673 | <0.001 | 0.135 |
| GO:0050803 | regulation of synapse structure or activity | 225 | 66 | -0.330 | -1.668 | <0.001 | 0.128 |
| GO:0048013 | ephrin receptor signaling pathway | 46 | 17 | -0.428 | -1.654 | 0.010 | 0.134 |
| GO:1990868 | response to chemokine | 56 | 27 | 0.481 | 1.975 | <0.001 | 0.007 |
| GO:0045785 | positive regulation of cell adhesion | 361 | 131 | 0.371 | 2.003 | <0.001 | 0.006 |
| GO:0007229 | integrin-mediated signaling pathway | 91 | 39 | 0.445 | 2.005 | <0.001 | 0.007 |
| GO:0050886 | endocrine process | 59 | 33 | 0.480 | 2.016 | <0.001 | 0.006 |
| GO:0002347 | response to tumor cell | 34 | 19 | 0.549 | 2.034 | <0.001 | 0.004 |
| GO:0060840 | artery development | 85 | 31 | 0.466 | 2.092 | <0.001 | 0.002 |
| GO:0060976 | coronary vasculature development | 40 | 16 | 0.550 | 2.095 | <0.001 | 0.003 |
| GO:0019882 | antigen processing and presentation | 92 | 45 | 0.493 | 2.232 | <0.001 | <0.001 |
| GO:0002181 | cytoplasmic translation | 148 | 90 | 0.469 | 2.262 | <0.001 | <0.001 |
| GO:0002396 | MHC protein complex assembly | 18 | 13 | 0.733 | 2.280 | <0.001 | <0.001 |

ES = Enrichment score; NES = Normalized enrichment score.

**TABLE S8b:** Summary of gene set enrichment analysis of cellular components for PLS1.

| Gene Set | Description | Size | Leading Edge Number | ES | NES | P value | P<sub>FDR</sub> |
| --- | --- | --- | --- | --- | --- | --- | --- |
| GO:0060076 | excitatory synapse | 62 | 21 | -0.510 | -2.068 | <0.001 | 0.009 |
| GO:0098978 | glutamatergic synapse | 390 | 136 | -0.359 | -1.952 | <0.001 | 0.012 |
| GO:0044298 | cell body membrane | 29 | 15 | -0.535 | -1.879 | 0.004 | 0.019 |
| GO:0097060 | synaptic membrane | 354 | 145 | -0.350 | -1.877 | <0.001 | 0.014 |
| GO:0030427 | site of polarized growth | 153 | 48 | -0.375 | -1.849 | <0.001 | 0.016 |
| GO:0043198 | dendritic shaft | 40 | 16 | -0.498 | -1.849 | <0.001 | 0.013 |
| GO:0044304 | main axon | 60 | 20 | -0.441 | -1.826 | <0.001 | 0.013 |
| GO:0098984 | neuron to neuron synapse | 339 | 104 | -0.333 | -1.788 | <0.001 | 0.019 |
| GO:0044309 | neuron spine | 165 | 55 | -0.365 | -1.786 | <0.001 | 0.018 |
| GO:0099572 | postsynaptic specialization | 320 | 135 | -0.334 | -1.778 | <0.001 | 0.018 |
| GO:0005775 | vacuolar lumen | 139 | 63 | 0.321 | 1.554 | 0.002 | 0.096 |
| GO:0101031 | protein folding chaperone complex | 40 | 21 | 0.431 | 1.607 | 0.019 | 0.077 |
| GO:0005796 | Golgi lumen | 65 | 34 | 0.408 | 1.718 | 0.002 | 0.031 |
| GO:0072562 | blood microparticle | 71 | 36 | 0.415 | 1.779 | <0.001 | 0.021 |
| GO:0005840 | ribosome | 230 | 116 | 0.386 | 1.982 | <0.001 | 0.001 |
| GO:0098636 | protein complex involved in cell adhesion | 39 | 19 | 0.531 | 2.019 | <0.001 | 0.001 |
| GO:0009897 | external side of plasma membrane | 237 | 98 | 0.393 | 2.042 | <0.001 | 0.001 |
| GO:0062023 | collagen-containing extracellular matrix | 298 | 129 | 0.392 | 2.073 | <0.001 | 0.001 |
| GO:0098576 | lumenal side of membrane | 34 | 17 | 0.656 | 2.379 | <0.001 | <0.001 |
| GO:0042611 | MHC protein complex | 21 | 17 | 0.818 | 2.608 | <0.001 | <0.001 |

ES = Enrichment score; NES = Normalized enrichment score.

## References

[1] Max A. Laansma, Joanna K. Bright, Sarah Al-Bachari, Tim J. Anderson, Tyler Ard, Francesca Assogna, Katherine A. Baquero, Henk W. Berendse, Jamie Blair, Fernando Cendes, John C. Dalrymple-Alford, Rob M.A. Bie, Ines Debove, Michiel F. Dirkx, Jason Druzgal, Hedley C.A. Emsley, Gäetan Garraux, Rachel P. Guimarães, Boris A. Gutman, Rick C. Helmich, Johannes C. Klein, Clare E. Mackay, Corey T. McMillan, Tracy R. Melzer, Laura M. Parkes, Fabrizio Piras, Toni L. Pitcher, Kathleen L. Poston, Mario Rango, Letícia F. Ribeiro, Cristiane S. Rocha, Christian Rummel, Lucas S.R. Santos, Reinhold Schmidt, Petra Schwingenschuh, Gianfranco Spalletta, Letizia Squarcina, Odile A. Heuvel, Chris Vriend, Jiun-Jie Wang, Daniel Weintraub, Roland Wiest, Clarissa L. Yasuda, Neda Jahanshad, Paul M. Thompson, Ysbrand D. Werf, and The ENIGMA-Parkinson's Study. International multicenter analysis of brain structure across clinical stages of parkinson’s disease. Movement Disorders, 36(11):2583–2594, 01 2021.

[2] Elijah Mak, Li Su, Guy B. Williams, Michael J. Firbank, Rachael A. Lawson, Alison J. Yarnall, Gordon W. Duncan, Adrian M. Owen, Tien K. Khoo, David J. Brooks, James B. Rowe, Roger A. Barker, David J. Burn, and John T. O’Brien. Baseline and longitudinal grey matter changes in newly diagnosed parkinson’s disease: ICICLE-PD study. Brain, 138(10):2974–2986, 2015.

[3] Yashar Zeighami, Miguel Ulla, Yasser Iturria-Medina, Mahsa Dadar, Yu Zhang, Kevin Michel-Herve Larcher, Vladimir Fonov, Alan C Evans, D Louis Collins, and Alain Dagher. Network structure of brain atrophy in de novo parkinson’s disease. eLife, 4:e08440, 09 2015.

[4] Heiko Braak, Kelly Del Tredici, Udo Rüb, Rob A.I de Vos, Ernst N.H Jansen Steur, and Eva Braak. Staging of brain pathology related to sporadic parkinson’s disease. Neurobiology of Aging, 24(2):197–211, 2003.

[5] Heiko Braak, Udo Rüb, and Kelly Del Tredici. Cognitive decline correlates with neuropathological stage in parkinson’s disease. Journal of the Neurological Sciences, 248(1-2):255–258, 10 2006.

[6] William W. Seeley, Richard K. Crawford, Juan Zhou, Bruce L. Miller, and Michael D. Greicius. Neurodegenerative diseases target large-scale human brain networks. Neuron, 62(1):42–52, 2009.

[7] Jason D. Warren, Jonathan D. Rohrer, Jonathan M. Schott, Nick C. Fox, John Hardy, and Martin N. Rossor. Molecular nexopathies: a new paradigm of neurodegenerative disease. Trends in Neurosciences, 36(10):561–569, 2013.

[8] Mathias Jucker and Lary C. Walker. Self-propagation of pathogenic protein aggregates in neurodegenerative diseases. Nature, 501(7465):45–51, 2013.

[9] Kelvin C. Luk, Victoria Kehm, Jenna Carroll, Bin Zhang, Patrick O’Brien, John Q. Trojanowski, and Virginia M.-Y. Lee. Pathological -synuclein transmission initiates parkinson-like neurodegeneration in nontransgenic mice. Science, 338(6109):949–953, 2012.

[10] Masami Masuda-Suzukake, Takashi Nonaka, Masato Hosokawa, Takayuki Oikawa, Tetsuaki Arai, Haruhiko Akiyama, David M. A. Mann, and Masato Hasegawa. Prion-like spreading of pathological -synuclein in brain. Brain, 136(4):1128–1138, 2013.

[11] Anne-Laure Mougenot, Simon Nicot, Anna Bencsik, Eric Morignat, Jérémy Verchère, Latefa Lakhdar, Stéphane Legastelois, and Thierry Baron. Prion-like acceleration of a synucleinopathy in a transgenic mouse model. Neurobiology of Aging, 33(9):2225–2228, 2012.

[12] Silvia Basaia, Federica Agosta, Ibai Diez, Elisenda Bueichekú, Federico d’Oleire Uquillas, Manuel Delgado-Alvarado, César Caballero-Gaudes, MariCruz Rodriguez-Oroz, Tanja Stojkovic, Vladimir S. Kostic, Massimo Filippi, and Jorge Sepulcre. Neurogenetic traits outline vulnerability to cortical disruption in parkinson’s disease. NeuroImage: Clinical, 33:102941, 2022.

[13] S. Pandya, Y. Zeighami, B. Freeze, M. Dadar, D.L. Collins, A. Dagher, and A. Raj. Predictive model of spread of parkinson’s pathology using network diffusion. NeuroImage, 192:178–194, 03 2019.

[14] Shady Rahayel, Christina Tremblay, Andrew Vo, Bratislav Misic, Stéphane Lehéricy, Isabelle Arnulf, Marie Vidailhet, Jean-Christophe Corvol, Marie Vidailhet, Jean-Christophe Corvol, Isabelle Arnulf, Stéphane Lehéricy, Marie Vidailhet, Graziella Mangone, Jean-Christophe Corvol, Isabelle Arnulf, Sara Sambin, Jonas Ihle, Caroline Weill, David Grabli, Florence Cormier-Dequaire, Louise Laure Mariani, Bertrand Degos, Richard Levy, Fanny Pineau, Julie Socha, Eve Benchetrit, Virginie Czernecki, Marie-Alexandrine Glachant, Sophie Rivaud-Pechoux, Elodie Hainque, Isabelle Arnulf, Smaranda Leu Semenescu, Pauline Dodet, Jean-Christophe Corvol, Graziella Mangone, Samir Bekadar, Alexis Brice, Suzanne Lesage, Fanny Mochel, Farid Ichou, Vincent Perlbarg, Benoit Colsch, Arthur Tenenhaus, Stéphane Lehéricy, Rahul Gaurav, Nadya Pyatigorskaya, Lydia Yahia-Cherif, Romain Valabrègue, Cécile Galléa, Marie-Odile Habert, Dijana Petrovska, Laetitia Jeancolas, Vanessa Brochard, Alizé Chalançon, Carole Dongmo-Kenfack, Christelle Laganot, Valentine Maheo, Jean-François Gagnon, Ronald B Postuma, Jacques Montplaisir, Simon Lewis, Elie Matar, Kaylena Ehgoetz Martens, Per Borghammer, Karoline Knudsen, Allan K Hansen, Oury Monchi, Ziv Gan-Or, Alain Dagher, and for the Alzheimer's Disease Neuroimaging Initiative. Mitochondrial function-associated genes underlie cortical atrophy in prodromal synucleinopathies. Brain, 146(8):3301–3318, 2023.

[15] Christina Tremblay, Shady Rahayel, Andrew Vo, Filip Morys, Golia Shafiei, Nooshin Abbasi, Ross D Markello, Ziv Gan-Or, Bratislav Misic, and Alain Dagher. Brain atrophy progression in parkinson’s disease is shaped by connectivity and local vulnerability. Brain Communications, 3(4):fcab269, 2021.

[16] Andrew Vo, Christina Tremblay, Shady Rahayel, Golia Shafiei, Justine Y. Hansen, Yvonne Yau, Bratislav Misic, and Alain Dagher. Network connectivity and local transcriptomic vulnerability underpin cortical atrophy progression in parkinson’s disease. NeuroImage: Clinical, 40:103523, 2023.

[17] Y. Yau, Y. Zeighami, T. E. Baker, K. Larcher, U. Vainik, M. Dadar, V. S. Fonov, P. Hagmann, A. Griffa, B. Mišić, D. L. Collins, and A. Dagher. Network connectivity determines cortical thinning in early parkinson’s disease progression. Nature Communications, 9(1):12, 01 2018.

[18] Per Borghammer and Nathalie Van Den Berge. Brain-first versus gut-first parkinson’s disease: A hypothesis. Journal of Parkinson’s Disease, 9(Suppl 2):S281–S295, 2019.

[19] Yongbin Wei, Lianne H. Scholtens, Elise Turk, and Martijn P. van den Heuvel. Multiscale examination of cytoarchitectonic similarity and human brain connectivity. Network Neuroscience, 3(1):124–137, 2018.

[20] Juan Zhou, Efstathios D. Gennatas, Joel H. Kramer, Bruce L. Miller, and William W. Seeley. Predicting regional neurodegeneration from the healthy brain functional connectome. Neuron, 73(6):1216–1227, 2012.

[21] Aurina Arnatkeviciute, Ben D. Fulcher, Mark A. Bellgrove, and Alex Fornito. Imaging transcriptomics of brain disorders. Biological Psychiatry Global Open Science, 2(4):319– 331, 2022.

[22] Shady Rahayel, Christina Tremblay, Andrew Vo, Ying Qiu Zheng, Stéphane Lehéricy, Isabelle Arnulf, Marie Vidailhet, Jean Christophe Corvol, Marie Vidailhet, Jean-Christophe Corvol, Isabelle Arnulf, Stéphane Lehéricy, Graziella Mangone, Sara Sambin, Jonas Ihle, Caroline Weill, David Grabli, Florence Cormier-Dequaire, Louise Laure Mariani, Bertrand Degos, Richard Levy, Fanny Pineau, Julie Socha, Eve Benchetrit, Virginie Czernecki, Marie-Alexandrine Glachant, Sophie Rivaud-Pechoux, Elodie Hainque, Smaranda Leu Semenescu, Pauline Dodet, Samir Bekadar, Alexis Brice, Suzanne Lesage, Fanny Mochel, Farid Ichou, Vincent Perlbarg, Benoit Colsch, Arthur Tenenhaus, Rahul Gaurav, Nadya Pyatigorskaya, Lydia Yahia-Cherif, Romain Valabrègue, Cécile Galléa, Marie-Odile Habert, Dijana Petrovska, Laetitia Jeancolas, Vanessa Brochard, Alizé Chalançon, Carole Dongmo-Kenfack, Christelle Laganot, Valentine Maheo, Jean François Gagnon, Ronald B Postuma, Jacques Montplaisir, Simon Lewis, Elie Matar, Kaylena Ehgoetz Martens, Per Borghammer, Karoline Knudsen, Allan Hansen, Oury Monchi, Bratislav Misic, and Alain Dagher. Brain atrophy in prodromal synucleinopathy is shaped by structural connectivity and gene expression. Brain, 145(9):3162–3178, 2022.

[23] George E C Thomas, Angeliki Zarkali, Mina Ryten, Karin Shmueli, Ana Luisa Gil Martinez, Louise-Ann Leyland, Peter McColgan, Julio Acosta-Cabronero, Andrew J Lees, and Rimona S Weil. Regional brain iron and gene expression provide insights into neurodegeneration in parkinson’s disease. Brain, 144(6):awab084–, 2021.

[24] Paul M Thompson, Jason L Stein, Sarah E Medland, Derrek P Hibar, Alejandro Arias Vasquez, Miguel E Renteria, Roberto Toro, Neda Jahanshad, Gunter Schumann, Barbara Franke, Margaret J Wright, Nicholas G Martin, Ingrid Agartz, Martin Alda, Saud Alhusaini, Laura Almasy, Jorge Almeida, Kathryn Alpert, Nancy C Andreasen, Ole A Andreassen, Liana G Apostolova, Katja Appel, Nicola J Armstrong, Benjamin Aribisala, Mark E Bastin, Michael Bauer, Carrie E Bearden, Orjan Bergmann, Elisabeth B Binder, John Blangero, Henry J Bockholt, Erlend Bøen, Catherine Bois, Dorret I Boomsma, Tom Booth, Ian J Bowman, Janita Bralten, Rachel M Brouwer, Han G Brunner, David G Brohawn, Randy L Buckner, Jan Buitelaar, Kazima Bulayeva, Juan R Bustillo, Vince D Calhoun, Dara M Cannon, Rita M Cantor, Melanie A Carless, Xavier Caseras, Gianpiero L Cavalleri, M Mallar Chakravarty, Kiki D Chang, Christopher R K Ching, Andrea Christoforou, Sven Cichon, Vincent P Clark, Patricia Conrod, Giovanni Coppola, Benedicto Crespo-Facorro, Joanne E Curran, Michael Czisch, Ian J Deary, Eco J C de Geus, Anouk den Braber, Giuseppe Delvecchio, Chantal Depondt, Lieuwe de Haan, Greig I de Zubicaray, Danai Dima, Rali Dimitrova, Srdjan Djurovic, Hongwei Dong, Gary Donohoe, Ravindranath Duggirala, Thomas D Dyer, Stefan Ehrlich, Carl Johan Ekman, Torbjørn Elvsåshagen, Louise Emsell, Susanne Erk, Thomas Espeseth, Jesen Fagerness, Scott Fears, Iryna Fedko, Guillén Fernández, Simon E Fisher, Tatiana Foroud, Peter T Fox, Clyde Francks, Sophia Frangou, Eva Maria Frey, Thomas Frodl, Vincent Frouin, Hugh Garavan, Sudheer Giddaluru, David C Glahn, Beata Godlewska, Rita Z Goldstein, Randy L Gollub, Hans J Grabe, Oliver Grimm, Oliver Gruber, Tulio Guadalupe, Raquel E Gur, Ruben C Gur, Harald HH Göring, Saskia Hagenaars, Tomas Hajek, Geoffrey B Hall, Jeremy Hall, John Hardy, Catharina A Hartman, Johanna Hass, Sean N Hatton, Unn K Haukvik, Katrin Hegenscheid, Andreas Heinz, Ian B Hickie, Beng-Choon Ho, David Hoehn, Pieter J Hoekstra, Marisa Hollinshead, Avram J Holmes, Georg Homuth, Martine Hoogman, L Elliot Hong, Norbert Hosten, Jouke-Jan Hottenga, Hilleke E Hulshoff Pol, Kristy S Hwang, Clifford R Jack, Mark Jenkinson, Caroline Johnston, Erik G Jönsson, René S Kahn, Dalia Kasperaviciute, Sinead Kelly, Sungeun Kim, Peter Kochunov, Laura Koenders, Bernd Krämer, John B J Kwok, Jim Lagopoulos, Gonzalo Laje, Mikael Landen, Bennett A Landman, John Lauriello, Stephen M Lawrie, Phil H Lee, Stephanie Le Hellard, Herve Lemaître, Cassandra D Leonardo, Chiang-Shan Li, Benny Liberg, David C Liewald, Xinmin Liu, Lorna M Lopez, Eva Loth, Anbarasu Lourdusamy, Michelle Luciano, Fabio Macciardi, Marise W J Machielsen, Glenda M Macqueen, Ulrik F Malt, René Mandl Dara S Manoach, Jean-Luc Martinot, Mar Matarin, Karen A Mather, Manuel Mattheisen, Morten Mattingsdal, Andreas Meyer-Lindenberg, Colm McDonald, Andrew M McIntosh, Francis J McMahon, Katie L McMahon, Eva Meisenzahl, Ingrid Melle, Yuri Milaneschi, Sebastian Mohnke, Grant W Montgomery, Derek W Morris, Eric K Moses, Bryon A Mueller, Susana Muñoz Maniega, Thomas W Mühleisen, Bertram Müller-Myhsok, Benson Mwangi, Matthias Nauck, Kwangsik Nho, Thomas E Nichols, Lars-Göran Nilsson, Allison C Nugent, Lars Nyberg, Rene L Olvera, Jaap Oosterlaan, Roel A Ophoff, Massimo Pandolfo, Melina Papalampropoulou-Tsiridou, Martina Papmeyer, Tomas Paus, Zdenka Pausova, Godfrey D Pearlson, Brenda W Penninx, Charles P Peterson, Andrea Pfennig, Mary Phillips, G Bruce Pike, Jean-Baptiste Poline, Steven G Potkin, Benno Pütz, Adaikalavan Ramasamy, Jerod Rasmussen, Marcella Rietschel, Mark Rijpkema, Shannon L Risacher, Joshua L Roffman, Roberto Roiz-Santiañez, Nina Romanczuk-Seiferth, Emma J Rose, Natalie A Royle, Dan Rujescu, Mina Ryten, Perminder S Sachdev, Alireza Salami, Theodore D Satterthwaite, Jonathan Savitz, Andrew J Saykin, Cathy Scanlon, Lianne Schmaal, Hugo G Schnack, Andrew J Schork, S Charles Schulz, Remmelt Schür, Larry Seidman, Li Shen, Jody M Shoemaker, Andrew Simmons, Sanjay M Sisodiya, Colin Smith, Jordan W Smoller, Jair C Soares, Scott R Sponheim, Emma Sprooten, John M Starr, Vidar M Steen, Stephen Strakowski, Lachlan Strike, Jessika Sussmann, Philipp G Sämann, Alexander Teumer, Arthur W Toga, Diana Tordesillas-Gutierrez, Daniah Trabzuni, Sarah Trost, Jessica Turner, Martijn Van den Heuvel, Nic J van der Wee, Kristel van Eijk, Theo G M van Erp, Neeltje E M van Haren, Dennis van ‘t Ent, Marie-Jose van Tol, Maria C Valdés Hernández, Dick J Veltman, Amelia Versace, Henry Völzke, Robert Walker, Henrik Walter, Lei Wang, Joanna M Wardlaw, Michael E Weale, Michael W Weiner, Wei Wen, Lars T Westlye, Heather C Whalley, Christopher D Whelan, Tonya White, Anderson M Winkler, Katharina Wittfeld, Girma Woldehawariat, Christiane Wolf, David Zilles, Marcel P Zwiers, Anbupalam Thalamuthu, Peter R Schofield, Nelson B Freimer, Natalia S Lawrence, Wayne Drevets, and EPIGEN Consortium IMAGEN Consortium Saguenay Youth Study (SYS) Group, Alzheimer’s Disease Neuroimaging Initiative. The ENIGMA consortium: large-scale collaborative analyses of neuroimaging and genetic data. Brain Imaging and Behavior, 8(2):153–182, 2014.

[25] Paul M. Thompson, Neda Jahanshad, Lianne Schmaal, Jessica A. Turner, Anderson M. Winkler, Sophia I. Thomopoulos, Gary F. Egan, and Peter Kochunov. The enhancing NeuroImaging genetics through meta-analysis consortium: 10 years of global collaborations in human brain mapping. Human Brain Mapping, 43(1):15–22, 2022.

[26] Margaret M. Hoehn and Melvin D. Yahr. Parkinsonism onset, progression, and mortality. Neurology, 17(5):427– 427, 1967.

[27] Rahul S. Desikan, Florent Ségonne, Bruce Fischl, Brian T. Quinn, Bradford C. Dickerson, Deborah Blacker, Randy L. Buckner, Anders M. Dale, R. Paul Maguire, Bradley T. Hyman, Marilyn S. Albert, and Ronald J. Killiany. An automated labeling system for subdividing the human cerebral cortex on MRI scans into gyral based regions of interest. NeuroImage, 31(3):968–980, 2006.

[28] Ziad S. Nasreddine, Natalie A. Phillips, Valérie Bédirian, Simon Charbonneau, Victor Whitehead, Isabelle Collin, Jeffrey L. Cummings, and Howard Chertkow. The montreal cognitive assessment, MoCA: A brief screening tool for mild cognitive impairment. Journal of the American Geriatrics Society, 53(4):695–699, 2005.

[29] Jennifer Stine Elam, Matthew F. Glasser, Michael P. Harms, Stamatios N. Sotiropoulos, Jesper L.R. Andersson, Gregory C. Burgess, Sandra W. Curtiss, Robert Oostenveld, Linda J. Larson-Prior, Jan-Mathijs Schoffelen, Michael R. Hodge, Eileen A. Cler, Daniel M. Marcus, Deanna M. Barch, Essa Yacoub, Stephen M. Smith, Kamil Ugurbil, and David C. Van Essen. The human connectome project: A retrospective. NeuroImage, 244:118543, 2021.

[30] B. T. Thomas Yeo, Fenna M. Krienen, Jorge Sepulcre, Mert R. Sabuncu, Danial Lashkari, Marisa Hollinshead, Joshua L. Roffman, Jordan W. Smoller, Lilla Zöllei, Jonathan R. Polimeni, Bruce Fischl, Hesheng Liu, and Randy L. Buckner. The organization of the human cerebral cortex estimated by intrinsic functional connectivity. Journal of Neurophysiology, 106(3):1125–1165, 2011.

[31] Constantin Freiherr von Economo and Georg N Koskinas. Die cytoarchitektonik der hirnrinde des erwachsenen menschen. J. Springer, 1925.

[32] Lianne H. Scholtens, Marcel A. de Reus, Siemon C. de Lange, Ruben Schmidt, and Martijn P. van den Heuvel. An MRI von economo – koskinas atlas. NeuroImage, 170:249–256, 2018.

[33] Justine Y. Hansen, Golia Shafiei, Ross D. Markello, Kelly Smart, Sylvia M. L. Cox, Martin Nørgaard, Vincent Beliveau, Yanjun Wu, Jean-Dominique Gallezot, Étienne Aumont, Stijn Servaes, Stephanie G. Scala, Jonathan M. DuBois, Gabriel Wainstein, Gleb Bezgin, Thomas Funck, Taylor W. Schmitz, R. Nathan Spreng, Marian Galovic, Matthias J. Koepp, John S. Duncan, Jonathan P. Coles, Tim D. Fryer, Franklin I. Aigbirhio, Colm J. McGinnity, Alexander Hammers, Jean-Paul Soucy, Sylvain Baillet, Synthia Guimond, Jarmo Hietala, Marc-André Bedard Marco Leyton, Eliane Kobayashi, Pedro Rosa-Neto, Melanie Ganz, Gitte M. Knudsen, Nicola Palomero-Gallagher, James M. Shine, Richard E. Carson, Lauri Tuominen, Alain Dagher, and Bratislav Misic. Mapping neurotransmitter systems to the structural and functional organization of the human neocortex. Nature Neuroscience, 25(11):1569–1581, 2022.

[34] Ross D. Markello, Justine Y. Hansen, Zhen-Qi Liu, Vincent Bazinet, Golia Shafiei, Laura E. Suárez, Nadia Blostein, Jakob Seidlitz, Sylvain Baillet, Theodore D. Satterthwaite, M. Mallar Chakravarty, Armin Raznahan, and Bratislav Misic. neuromaps: structural and functional interpretation of brain maps. Nature Methods, 19(11):1472–1479, 2022.

[35] Michael J. Hawrylycz, Ed S. Lein, Angela L. Guillozet-Bongaarts, Elaine H. Shen, Lydia Ng, Jeremy A. Miller, Louie N. van de Lagemaat, Kimberly A. Smith, Amanda Ebbert, Zackery L. Riley, Chris Abajian, Christian F. Beckmann, Amy Bernard, Darren Bertagnolli, Andrew F. Boe, Preston M. Cartagena, M. Mallar Chakravarty, Mike Chapin, Jimmy Chong, Rachel A. Dalley, Barry David Daly, Chinh Dang, Suvro Datta, Nick Dee, Tim A. Dolbeare, Vance Faber, David Feng, David R. Fowler, Jeff Goldy, Benjamin W. Gregor, Zeb Haradon, David R. Haynor, John G. Hohmann, Steve Horvath, Robert E. Howard, Andreas Jeromin, Jayson M. Jochim, Marty Kinnunen, Christopher Lau, Evan T. Lazarz, Changkyu Lee, Tracy A. Lemon, Ling Li, Yang Li, John A. Morris, Caroline C. Overly, Patrick D. Parker, Sheana E. Parry, Melissa Reding, Joshua J. Royall, Jay Schulkin, Pedro Adolfo Sequeira, Clifford R. Slaughterbeck, Simon C. Smith, Andy J. Sodt, Susan M. Sunkin, Beryl E. Swanson, Marquis P. Vawter, Derric Williams, Paul Wohnoutka, H. Ronald Zielke, Daniel H. Geschwind, Patrick R. Hof, Stephen M. Smith, Christof Koch, Seth G. N. Grant, and Allan R. Jones. An anatomically comprehensive atlas of the adult human brain transcriptome. Nature, 489(7416):391–399, 2012.

[36] Rosa De Micco, Antonio Russo, and Alessandro Tessitore. Structural MRI in idiopathic parkinson’s disease. International Review of Neurobiology, 141(Journal of the Neurological Sciences 128 1995):405–438, 2018.

[37] Heather Wilson, Flavia Niccolini, Clelia Pellicano, and Marios Politis. Cortical thinning across parkinson’s disease stages and clinical correlates. Journal of the Neurological Sciences, 398:31–38, 2019. doi:10.1016/j.jns.2019.01.020.

[38] Mojtaba Zarei, Naroa Ibarretxe-Bilbao, Yaroslau Compta, Morgan Hough, Carme Junque, Nuria Bargallo, Eduardo Tolosa, and Maria Jose Martí. Cortical thinning is associated with disease stages and dementia in parkinson’s disease. Journal of Neurology, Neurosurgery & Psychiatry, 84(8):875, 2013.

[39] Ashish Raj, Amy Kuceyeski, and Michael Weiner. A network diffusion model of disease progression in dementia. Neuron, 73(6):1204–1215, 2012.

[40] Sveva Fornari, Amelie Schäfer, Mathias Jucker, Alain Goriely, and Ellen Kuhl. Prion-like spreading of alzheimer’s disease within the brain’s connectome. Journal of the Royal Society Interface, 16(159):20190356, 2019.

[41] Jil M. Meier, Hannelore K. Burgh, Abram D. Nitert, Peter Bede, Siemon C. Lange, Orla Hardiman, Leonard H. Berg, and Martijn P. Heuvel. Connectome-based propagation model in amyotrophic lateral sclerosis. Annals of Neurology, 87(5):725–738, 02 2020.

[42] Magdalini Polymenidou and Don W. Cleveland. The seeds of neurodegeneration: Prion-like spreading in ALS. Cell, 147(3):498–508, 2011.

[43] Golia Shafiei, Ross D. Markello, Carolina Makowski, Alexandra Talpalaru, Matthias Kirschner, Gabriel A. Devenyi, Elisa Guma, Patric Hagmann, Neil R. Cashman, Martin Lepage, M. Mallar Chakravarty, Alain Dagher, and Bratislav Mišić. Spatial patterning of tissue volume loss in schizophrenia reflects brain network architecture. Biological Psychiatry, 87(8):727–735, 2020.

[44] Golia Shafiei, Vincent Bazinet, Mahsa Dadar, Ana L Manera, D Louis Collins, Alain Dagher, Barbara Borroni, Raquel Sanchez-Valle, Fermin Moreno, Robert Laforce, Caroline Graff, Matthis Synofzik, Daniela Galimberti, James B Rowe, Mario Masellis, Maria Carmela Tartaglia, Elizabeth Finger, Rik Vandenberghe, Alexandre de Mendonça, Fabrizio Tagliavini, Isabel Santana, Chris Butler, Alex Gerhard, Adrian Danek, Johannes Levin, Markus Otto, Sandro Sorbi, Lize C Jiskoot, Harro Seelaar, John C van Swieten, Jonathan D Rohrer, Bratislav Misic, Simon Ducharme, Frontotemporal Lobar Degeneration Neuroimaging Initiative (FTLDNI), Howard Rosen, Bradford C Dickerson, Kimoko Domoto-Reilly, David Knopman, Bradley F Boeve, Adam L Boxer, John Kornak, Bruce L Miller, William W Seeley, Maria-Luisa Gorno-Tempini, Scott McGinnis, Maria Luisa Mandelli, GENetic Frontotemporal dementia Initiative (GENFI), Aitana Sogorb Esteve, Annabel Nelson, Arabella Bouzigues, Carolin Heller, Caroline V Greaves, David Cash, David L Thomas, Emily Todd, Hanya Benotmane, Henrik Zetterberg, Imogen J Swift, Jennifer Nicholas, Kiran Samra, Lucy L Russell, Martina Bocchetta, Rachelle Shafei, Rhian S Convery, Carolyn Timberlake, Thomas Cope, Timothy Rittman, Alberto Benussi, Enrico Premi, Roberto Gasparotti, Silvana Archetti, Stefano Gazzina, Valentina Cantoni, Andrea Arighi, Chiara Fenoglio, Elio Scarpini, Giorgio Fumagalli, Vittoria Borracci, Giacomina Rossi, Giorgio Giaccone, Giuseppe Di Fede, Paola Caroppo, Pietro Tiraboschi, Sara Prioni, Veronica Redaelli, David Tang-Wai, Ekaterina Rogaeva, Miguel Castelo-Branco, Morris Freedman, Ron Keren, Sandra Black, Sara Mitchell, Christen Shoesmith, Robart Bartha, Rosa Rademakers, Emma van der Ende, Jackie Poos, Janne M Papma, Lucia Giannini, Rick van Minkelen, Yolande Pijnenburg, Benedetta Nacmias, Camilla Ferrari, Cristina Polito, Gemma Lombardi, Valentina Bessi, Michele Veldsman, Christin Andersson, Hakan Thonberg, Linn Öijerstedt, Vesna Jelic, Paul Thompson, Tobias Langheinrich, Albert Lladó, Anna Antonell, Jaume Olives, Mircea Balasa, Nuria Bargalló, Sergi Borrego-Ecija, Ana Verdelho, Carolina Maruta, Catarina B Ferreira, Gabriel Miltenberger, Frederico Simões do Couto, Alazne Gabilondo, Ana Gorostidi, Jorge Villanua, Marta Cañada, Mikel Tainta, Miren Zulaica, Myriam Barandiaran, Patricia Alves, Benjamin Bender, Carlo Wilke, Lisa Graf, Annick Vogels, Mathieu Vandenbulcke, Philip Van Damme, Rose Bruffaerts, Pedro Rosa-Neto, Serge Gauthier, Agnès Camuzat, Alexis Brice, Anne Bertrand, Aurélie Funkiewiez, Daisy Rinaldi, Dario Saracino, Olivier Colliot, Sabrina Sayah, Catharina Prix, Elisabeth Wlasich, Olivia Wagemann, Sandra Loosli, Sonja Schönecker, Tobias Hoegen, Jolina Lombardi, Sarah Anderl-Straub, Adeline Rollin, Gregory Kuchcinski, Maxime Bertoux, Thibaud Lebouvier, Vincent Deramecourt, Beatriz Santiago, Diana Duro, Maria João Leitão, Maria Rosario Almeida, Miguel Tábuas-Pereira, Sónia Afonso, Annerose Engel, and Maryna Polyakova. Network structure and transcriptomic vulnerability shape atrophy in frontotemporal dementia. Brain, 146(1):awac069, 2022.

[45] Elior Drori, Shai Berman, and Aviv A. Mezer. Mapping microstructural gradients of the human striatum in normal aging and parkinson’s disease. Science Advances, 8(28):eabm1971, 2022.

[46] Stephen J. Kish, Kathleen Shannak, and Oleh Hornykiewicz. Uneven pattern of dopamine loss in the striatum of patients with idiopathic parkinson’s disease. The New England Journal of Medicine, 318(14):876–880, 04 1988. Kish, SJ Shannak, K Hornykiewicz, O eng Research Support, Non-U.S. Gov’t 1988/04/07 N Engl J Med. 1988 Apr 7;318(14):876–80.

[47] Bogdan Draganski, Ferath Kherif, Stefan Klöppel, Philip A. Cook, Daniel C. Alexander, Geoff J.M. Parker, Ralf Deichmann, John Ashburner, and Richard S.J. Frackowiak. Evidence for segregated and integrative connectivity patterns in the human basal ganglia. The Journal of Neuroscience, 28(28):7143–7152, 2008.

[48] Peter Manza, Sheng Zhang, Chiang-Shan R. Li, and Hoi-Chung Leung. Resting-state functional connectivity of the striatum in early-stage parkinson’s disease: Cognitive decline and motor symptomatology. Human Brain Mapping, 37(2):648–662, 11 2016.

[49] Andrew J. Peters, Julie M. J. Fabre, Nicholas A. Steinmetz, Kenneth D. Harris, and Matteo Carandini. Striatal activity topographically reflects cortical activity. Nature, 591(7850):420–425, 2021.

[50] Ye Tian, Daniel S. Margulies, Michael Breakspear, and Andrew Zalesky. Topographic organization of the human subcortex unveiled with functional connectivity gradients. Nature Neuroscience, 23(11):1421–1432, 2020.

[51] K. Guadalupe Cruz, Yi Ning Leow, Nhat Minh Le, Elie Adam, Rafiq Huda, and Mriganka Sur. Cortical-subcortical interactions in goal-directed behavior. Physiological Reviews, 103(1):347–389, 2023.

[52] Danielle S Bassett and Olaf Sporns. Network neuroscience. Nature Neuroscience, 20(3):353–364, 2017.

[53] Ed Bullmore and Olaf Sporns. Complex brain networks: graph theoretical analysis of structural and functional systems. Nature Reviews Neuroscience, 10(3):186–198, 2009.

[54] Martijn P. van den Heuvel and Olaf Sporns. Rich-club organization of the human connectome. The Journal of Neuroscience, 31(44):15775–15786, 2011.

[55] Jacob Horsager and Per Borghammer. Brain-first vs. body-first parkinson’s disease: An update on recent evidence. Parkinsonism & Related Disorders, 122:106101, 2024.

[56] Joana B. Pereira, Per Svenningsson, Daniel Weintraub, Kolbjørn Brønnick, Alexander Lebedev, Eric Westman, and Dag Aarsland. Initial cognitive decline is associated with cortical thinning in early parkinson disease. Neurology, 82(22):2017–2025, 06 2014.

[57] Carme Uribe, Barbara Segura, Hugo Cesar Baggio, Alexandra Abos, Maria Jose Marti, Francesc Valldeoriola, Yaroslau Compta, Nuria Bargallo, and Carme Junque. Patterns of cortical thinning in nondemented parkinson’s disease patients. Movement Disorders, 31(5):699–708, 04 2016.

[58] Thilo van Eimeren, Oury Monchi, Benedicte Ballanger, and Antonio P. Strafella. Dysfunction of the default mode network in parkinson disease: A functional magnetic resonance imaging study. Archives of Neurology, 66(7):877– 883, 2009.

[59] Yanbing Hou, Jing Yang, Chunyan Luo, Wei Song, Ruwei Ou, Wanglin Liu, Qiyong Gong, and Huifang Shang. Dysfunction of the default mode network in drug-naïve parkinson’s disease with mild cognitive impairments: A resting-state fMRI study. Frontiers in Aging Neuroscience, 8:247, 2016.

[60] Alessandro Tessitore, Fabrizio Esposito, Carmine Vitale, Gabriella Santangelo, Marianna Amboni, Antonio Russo, Daniele Corbo, Giovanni Cirillo, Paolo Barone, and Gioacchino Tedeschi. Default-mode network connectivity in cognitively unimpaired patients with parkinson disease. Neurology, 79(23):2226–2232, 2012.

[61] Hai Lin, Xiaodong Cai, Doudou Zhang, Jiali Liu, Peng Na, and Weiping Li. Functional connectivity markers of depression in advanced parkinson’s disease. NeuroImage: Clinical, 25:102130, 2020.

[62] Olaia Lucas-Jiménez, Natalia Ojeda, Javier Peña, María Díez-Cirarda, Alberto Cabrera-Zubizarreta, Juan Carlos Gómez-Esteban, María Ángeles Gómez-Beldarrain, and Naroa Ibarretxe-Bilbao. Altered functional connectivity in the default mode network is associated with cognitive impairment and brain anatomical changes in parkinson’s disease. Parkinsonism & Related Disorders, 33:58– 64, 2016.

[63] Marina C. Ruppert, Andrea Greuel, Julia Freigang, Masoud Tahmasian, Franziska Maier, Jochen Hammes, Thilo van Eimeren, Lars Timmermann, Marc Tittgemeyer, Alexander Drzezga, and Carsten Eggers. The default mode network and cognition in parkinson’s disease: A multimodal resting-state network approach. Human Brain Mapping, 42(8):2623–2641, 2021.

[64] Luqing Wei, Xiao Hu, Yajing Zhu, Yonggui Yuan, Weiguo Liu, and Hong Chen. Aberrant intra-and internetwork functional connectivity in depressed parkinson’s disease. Scientific Reports, 7(1):2568, 2017.

[65] Alok Shiomurti Tripathi, Needa Fatima, Pankaj Tripathi, Rina Tripathi, Alka, Magdi E. A. Zaki, Lucy Mohapatra, Mohammad Yasir, and Rahul K Maurya. Beneficial effect of 5-HT1b/1d agonist on parkinson’s disease by modulating glutamate and reducing deposition of -synuclein. Journal of Biochemical and Molecular Toxicology, 38(1):e23627, 2024.

[66] Maryka Quik, Susan Wonnacott, and Burt M. Sharp. 62* and 42* nicotinic acetylcholine receptors as drug targets for parkinson’s disease. Pharmacological Reviews, 63(4):938–966, 2011.

[67] Michael X. Henderson, Eli J. Cornblath, Adam Darwich, Bin Zhang, Hannah Brown, Ronald J. Gathagan, Raizel M. Sandler, Danielle S. Bassett, John Q. Trojanowski, and Virginia M. Y. Lee. Spread of -synuclein pathology through the brain connectome is modulated by selective vulnerability and predicted by network analysis. Nature Neuroscience, 22(8):1248–1257, 2019.

[68] Walter J. Schulz-Schaeffer. The synaptic pathology of -synuclein aggregation in dementia with lewy bodies, parkinson’s disease and parkinson’s disease dementia. Acta Neuropathologica, 120(2):131–143, 2010.

[69] Laura A. Volpicelli-Daley, Kelvin C. Luk, Tapan P. Patel, Selcuk A. Tanik, Dawn M. Riddle, Anna Stieber, David F. Meaney, John Q. Trojanowski, and Virginia M.-Y. Lee. Exogenous -synuclein fibrils induce lewy body pathology leading to synaptic dysfunction and neuron death. Neuron, 72(1):57–71, 2011.

[70] Didac Vidal-Pineiro, Nadine Parker, Jean Shin, Leon French, Håkon Grydeland, Andrea P Jackowski, Athanasia M Mowinckel, Yash Patel, Zdenka Pausova, Giovanni Salum, Øystein Sørensen, Kristine B Walhovd, Tomas Paus, Anders M Fjell, and ageing, Alzheimer's Disease Neuroimaging Initiative and the Australian Imaging Biomarkers and Lifestyle flagship study of. Cellular correlates of cortical thinning throughout the lifespan. Scientific Reports, 10(1):21803, 2020.

[71] Alaa Abdelgawad, Shady Rahayel, Ying-Qiu Zheng, Christina Tremblay, Andrew Vo, Bratislav Misic, and Alain Dagher. Predicting longitudinal brain atrophy in parkinson’s disease using a susceptible-infected-removed agent-based model. Network Neuroscience, 7(3):906–925, 2023.

[72] Ying-Qiu Zheng, Yu Zhang, Yvonne Yau, Yashar Zeighami, Kevin Larcher, Bratislav Misic, and Alain Dagher. Local vulnerability and global connectivity jointly shape neurodegenerative disease propagation. PLoS Biology, 17(11):e3000495, 11 2019.

[73] Christopher G. Goetz, Stanley Fahn, Pablo Martinez-Martin, Werner Poewe, Cristina Sampaio, Glenn T. Stebbins, Matthew B. Stern, Barbara C. Tilley, Richard Dodel, Bruno Dubois, Robert Holloway, Joseph Jankovic, Jaime Kulisevsky, Anthony E. Lang, Andrew Lees, Sue Leurgans, Peter A. LeWitt, David Nyenhuis, C. Warren Olanow, Olivier Rascol, Anette Schrag, Jeanne A. Teresi, Jacobus J. Van Hilten, and Nancy LaPelle. Movement disorder society-sponsored revision of the unified parkinson’s disease rating scale (MDS-UPDRS): Process, format, and clinimetric testing plan. Movement Disorders, 22(1):41– 47, 2007.

[74] Bruce Fischl. FreeSurfer. NeuroImage, 62(2):774–781, 2012.

[75] Jean-Philippe Fortin, Nicholas Cullen, Yvette I. Sheline, Warren D. Taylor, Irem Aselcioglu, Philip A. Cook, Phil Adams, Crystal Cooper, Maurizio Fava, Patrick J. McGrath, Melvin McInnis, Mary L. Phillips, Madhukar H. Trivedi, Myrna M. Weissman, and Russell T. Shinohara. Harmonization of cortical thickness measurements across scanners and sites. NeuroImage, 167:104–120, 2018.

[76] Renaud La Joie, Audrey Perrotin, Louisa Barré, Caroline Hommet, Florence Mézenge, Méziane Ibazizene, Vincent Camus, Ahmed Abbas, Brigitte Landeau, Denis Guilloteau, Vincent de La Sayette, Francis Eustache, Béatrice Desgranges, and Gaël Chételat. Region-specific hierarchy between atrophy, hypometabolism, and -amyloid (a) load in alzheimer’s disease dementia. The Journal of Neuroscience, 32(46):16265–16273, 11 2012.

[77] Josephine Barnes, Gerard R. Ridgway, Jonathan Bartlett, Susie M.D. Henley, Manja Lehmann, Nicola Hobbs, Matthew J. Clarkson, David G. MacManus, Sebastien Ourselin, and Nick C. Fox. Head size, age and gender adjustment in MRI studies: a necessary nuisance? NeuroImage, 53(4):1244–1255, 2010.

[78] Daniel E Ho, Kosuke Imai, Gary King, and Elizabeth A Stuart. MatchIt : Nonparametric preprocessing for parametric causal inference. Journal of Statistical Software, 42(8), 2011.

[79] Yoav Benjamini and Yosef Hochberg. Controlling the false discovery rate: A practical and powerful approach to multiple testing. Journal of the Royal Statistical Society: Series B (Methodological), 57(1):289–300, 1995.

[80] Aaron F. Alexander-Bloch, Haochang Shou, Siyuan Liu, Theodore D. Satterthwaite, David C. Glahn, Russell T. Shinohara, Simon N. Vandekar, and Armin Raznahan. On testing for spatial correspondence between maps of human brain structure and function. NeuroImage, 178:540– 551, 2018.

[81] Ross D. Markello and Bratislav Misic. Comparing spatial null models for brain maps. NeuroImage, 236:118052, 2021.

[82] František Váša and Bratislav Mišić. Null models in network neuroscience. Nature Reviews Neuroscience, 23(8):493–504, 2022.

[83] Sara Larivière, Casey Paquola, Bo-yong Park, Jessica Royer, Yezhou Wang, Oualid Benkarim, Reinder Vos de Wael, Sofie L. Valk, Sophia I. Thomopoulos, Matthias Kirschner, Lindsay B. Lewis, Alan C. Evans, Sanjay M. Sisodiya, Carrie R. McDonald, Paul M. Thompson, and Boris C. Bernhardt. The ENIGMA toolbox: multiscale neural contextualization of multisite neuroimaging datasets. Nature Methods, 18(7):698–700, 2021.

[84] Matthew F. Glasser, Stamatios N. Sotiropoulos, J. Anthony Wilson, Timothy S. Coalson, Bruce Fischl, Jesper L. Andersson, Junqian Xu, Saad Jbabdi, Matthew Webster, Jonathan R. Polimeni, David C. Van Essen, Mark Jenkinson, and for the WU-Minn HCP Consortium. The minimal preprocessing pipelines for the human connectome project. NeuroImage, 80:105–124, 2013.

[85] Gholamreza Salimi-Khorshidi, Gwenaëlle Douaud, Christian F. Beckmann, Matthew F. Glasser, Ludovica Griffanti, and Stephen M. Smith. Automatic denoising of functional MRI data: Combining independent component analysis and hierarchical fusion of classifiers. NeuroImage, 90:449–468, 2014.

[86] J-Donald Tournier, Robert Smith, David Raffelt, Rami Tabbara, Thijs Dhollander, Maximilian Pietsch, Daan Christiaens, Ben Jeurissen, Chun-Hung Yeh, and Alan Connelly. MRtrix3: A fast, flexible and open software framework for medical image processing and visualisation. NeuroImage, 202:116137, 2019.

[87] Robert E. Smith, Jacques-Donald Tournier, Fernando Calamante, and Alan Connelly. Anatomically-constrained tractography: Improved diffusion MRI streamlines tractography through effective use of anatomical information. NeuroImage, 62(3):1924–1938, 2012.

[88] Ben Jeurissen, Jacques-Donald Tournier, Thijs Dhollander, Alan Connelly, and Jan Sijbers. Multi-tissue constrained spherical deconvolution for improved analysis of multi-shell diffusion MRI data. NeuroImage, 103:411– 426, 2014.

[89] J-Donald Tournier, Fernando Calamante, and Alan Connelly. Robust determination of the fibre orientation distribution in diffusion MRI: Non-negativity constrained super-resolved spherical deconvolution. NeuroImage, 35(4):1459–1472, 2007.

[90] Robert E. Smith, Jacques-Donald Tournier, Fernando Calamante, and Alan Connelly. SIFT2: Enabling dense quantitative assessment of brain white matter connectivity using streamlines tractography. NeuroImage, 119:338– 351, 2015.

[91] Richard F. Betzel, Alessandra Griffa, Patric Hagmann, and Bratislav Mišić. Distance-dependent consensus thresholds for generating group-representative structural brain networks. Network Neuroscience, 3(2):475–496, 2019.

[92] Ross D Markello, Aurina Arnatkeviciute, Jean-Baptiste Poline, Ben D Fulcher, Alex Fornito, and Bratislav Misic. Standardizing workflows in imaging transcriptomics with the abagen toolbox. eLife, 10, 2021.

[93] Aurina Arnatkeviciūtė, Ben D. Fulcher, and Alex Fornito. A practical guide to linking brain-wide gene expression and neuroimaging data. NeuroImage, 189:353–367, 01 2019.

[94] Michael Hawrylycz, Jeremy A Miller, Vilas Menon, David Feng, Tim Dolbeare, Angela L Guillozet-Bongaarts, Anil G Jegga, Bruce J Aronow, Chang-Kyu Lee, Amy Bernard, Matthew F Glasser, Donna L Dierker, Jörg Menche, Aaron Szafer, Forrest Collman, Pascal Grange, Kenneth A Berman, Stefan Mihalas, Zizhen Yao, Lance Stewart, Albert-László Barabási, Jay Schulkin, John Phillips, Lydia Ng, Chinh Dang, David R Haynor, Allan Jones, David C Van Essen, Christof Koch, and Ed Lein. Canonical genetic signatures of the adult human brain. Nature Neuroscience, 18(12):1832–1844, 2015.

[95] Anjali Krishnan, Lynne J. Williams, Anthony Randal McIntosh, and Hervé Abdi. Partial least squares (PLS) methods for neuroimaging: A tutorial and review. NeuroImage, 56(2):455–475, 2011.

[96] Anthony Randal McIntosh and Nancy J. Lobaugh. Partial least squares analysis of neuroimaging data: applications and advances. NeuroImage, 23:S250–S263, 2004.

[97] John M Elizarraras, Yuxing Liao, Zhiao Shi, Qian Zhu, Alexander R Pico, and Bing Zhang. WebGestalt 2024: faster gene set analysis and new support for metabolomics and multi-omics. Nucleic Acids Research, 52(W1):W415–W421, 2024.

